# Epistorm-Mix: Mapping Social Contact Patterns for Respiratory Pathogen Spread in the Post-Pandemic United States

**DOI:** 10.1101/2025.11.20.25340662

**Authors:** Maria Litvinova, Shelly Sinclair, Allisandra G. Kummer, Paulo C. Ventura, Trevor Foster, Kayoko Shioda, M. Elizabeth Halloran, Alessandro Vespignani, Marco Ajelli

## Abstract

Human contact patterns are a fundamental determinant of respiratory pathogen transmission, yet nationally representative post-pandemic data for the United States are limited. We present Epistorm-Mix, a 2024 probability-based online survey designed to be nationally representative by age, sex, race/ethnicity, household income, census region, and language. Respondents reported all person-to-person contacts from the preceding day, including the contact’s age and the setting (household, school, workplace, or community). We quantified contact numbers across demographic and social characteristics and used generalized additive models to test adjusted differences. We constructed age-stratified contact matrices and their setting-specific counterparts, benchmarking them against widely used synthetic matrices to simulate the spread of an epidemic of a respiratory pathogen. We found an average of 7.4 contacts per day, with significant heterogeneity across the population. Contact rates were highest among teenagers (15-19 years) and lowest among older adults (60+ years). In-person attendance at school or work was a major driver, resulting in 2-3 times more contacts than remote participation. We also identified key socioeconomic and demographic group heterogeneities: the number of contacts generally increased with household income, and Non-Hispanic (NH) Black and NH Asian individuals reported statistically significant fewer total contacts than NH White individuals. We found strong assortative mixing by age and demographic group with markedly distinct contact patterns across different social settings (households, schools, workplaces, and the community). While the study’s age-mixing patterns are broadly comparable to international findings, the identified demographic heterogeneities reflect social structures unique to the US, underscoring the need for country-specific data. Epistorm-Mix provides a nationally representative portrait of post-pandemic US contact patterns and serves as an open-access resource for modeling and public health planning.

## Introduction

Understanding how people interact is central to infectious disease epidemiology, particularly for respiratory pathogens that propagate through close contact^1^. Quantifying these contact patterns across age groups, settings, and time is essential for identifying transmission pathways, estimating key epidemiological parameters, and designing effective public health interventions^1–6^. Over the past two decades, empirical contact data have revolutionized the mathematical and computational modeling of infectious diseases, enabling a shift from oversimplified assumptions to more realistic, data-informed approaches^7,8^. This transformation has significantly improved the predictive power and policy relevance of epidemic models, increasing their application in public health planning and response^9–12^.

Before the COVID-19 pandemic, landmark studies like POLYMOD^5^ provided foundational insights into human contact patterns, revealing key features such as age-assortative mixing and the importance of setting-specific interactions (e.g., in households, schools, and workplaces). However, these studies were geographically limited, covering only a subset of countries worldwide and often lacking national representativeness (e.g., focusing on a specific urban/rural area or population group, such as college students or adults)^7^. The pandemic further disrupted social interactions globally, with remote work, school closures, and restrictions on gatherings that have redefined contact behaviors in ways not well captured by pre-pandemic data^13^. While some studies have quantified pandemic-era contact patterns, their data were typically collected during periods of sustained non-pharmaceutical interventions, which limits their relevance to the current social landscape^13^. Moreover, even before the pandemic, comprehensive, nationally representative contact data were scarce for the US population. Existing US studies focused on specific age groups, geographic locations, or relied on convenience sub-samples^13–19^. These data gaps have led to the widespread use of synthetic contact matrices derived from census and demographic data, often combined with assumptions about social mixing^20–22^. While these models provide valuable approximations in the absence of empirical data, they require careful validation against observed contact patterns, as they may overlook key behavioral, cultural, and contextual factors that shape real-world interaction.

Here, we present the Epistorm-Mix study, which addresses this critical gap by measuring contact patterns across the US population in 2024, providing detailed insights into the structure of social interactions relevant to the spread of respiratory pathogens and essential for epidemiological modeling and public health planning. The study relies on a probability-based online survey panel explicitly designed to be representative of the US population, ensuring coverage across key demographic variables including age, gender, race/ethnicity, household income, geographic census region, and language. This sampling strategy enables inclusion of individuals from traditionally hard-to-reach groups, allowing for a comprehensive demographic understanding of contact behavior. The resulting data provide an essential empirical foundation for improving epidemic modeling and informing targeted public health interventions in the post-pandemic landscape.

All the Epistorm-Mix data analyzed in this study, including individual-level data and age- and setting-specific contact matrices, are openly available to the research and public health communities. To support further modeling efforts, we provide access to the full dataset and accompanying documentation through a dedicated webpage (https://www.epistorm.org/data/epistorm-mix) and GitHub repository (https://github.com/epistorm/Epistorm-Mix), ensuring broad usability and impact across epidemiological and policy domains.

## Results

To characterize in-person contact patterns in the post-pandemic US, we conducted a two-wave cross-sectional survey in May and October 2024. These two periods, both with schools in session and no COVID-19-related measures in place, were selected to provide a robust assessment of typical contact patterns and to serve as a baseline for understanding contact dynamics. A contact was defined as a person with whom the participant had an in-person conversation of at least five words or physical contact^5,23,24^. Physical contact was defined as any interaction that involves touching (e.g., a high-five, a hug). Participants were asked to complete a contact diary listing all the people with whom they, or their child (for participants aged 0-8 years old), had contact during the previous day (“yesterday”). Participants were encouraged to recall their day, starting from the time they woke up until they went to sleep. For each contact, we gathered sociodemographic information (e.g., age, sex, race/ethnicity) and the context of the interaction (e.g., contact setting). Respondents were enrolled from a probability-based online panel designed to be representative of the US population. Weights were applied to each respondent to ensure the sample was representative by age, sex, race/ethnicity, household income, geographic census region, and language. A total sample of 1,930 respondents was included in this analysis, with approximately half of the sample in each wave. On the diary day, 73.9% of students were at school, and 80.8% of employed or self-employed respondents were at work. The ages of the students ranged from 0 to 54 years, with 7.6% aged 0-2 years, 9.9% aged 3-4 years, 70.9% aged 5-17 years, and 11.5% aged 18 years or older. Among students attending school on the diary day, 95.8% were present in person; among participants working on the diary day, 70.1% were present in person. On the diary day, 10% of respondents reported feeling sick (9.9% in May and 10.1% in October). Details on the study participants are reported in the Methods and Supplementary Material.

### Daily Number of Contacts

On average, respondents reported 7.4 (IQR: 2.0–9.0) total contacts, where IQR stands for the interquartile range, corresponding to the 25th and 75th percentiles of the distribution (Table 1). Respondents 5-19 years old reported generally higher total contacts on average (10.3 contacts; IQR: 4.0 – 13.0), whereas the average number of contacts was lower among older adults (60+ years old). Hispanic/Latino and Non-Hispanic (NH) White respondents reported the largest number of daily contacts relative to other racial/ethnic groups, with 7.8 (IQR: 3.0 – 10.0) and 7.6 (IQR: 2.0 – 9.0) contacts, respectively. NH Asian and NH Black reported fewer contacts, with an average of 6.3 contacts per day for both (IQR: 2.0 – 8.0 and 2.0 – 7.0, respectively). Female and male respondents reported a similar number of contacts (7.7, IQR: 2.0 – 10.0 vs. 7.0, IQR: 2.0 – 9.0).

**Table 1.** Number of reported contacts across socio-demographic groups.

| Characteristic | N<sup>1</sup> |  | Total contacts<sup>2</sup> |
| --- | --- | --- | --- |
| Total Population | 1,930 (100%) |  | 7.4 (2.0 - 9.0) |

| Characteristic | N<sup>1</sup> | Total contacts<sup>2</sup> | Characteristic | N<sup>1</sup> | Total contacts<sup>2</sup> |
| --- | --- | --- | --- | --- | --- |
| **Main Activity and Mode of Attendance** |  |  | **Race/Ethnicity** |  |  |
| Student | 349 (18%) | 9.4 (4.0 - 12.0) | White, NH | 1,117 (58%) | 7.6 (2.0 - 9.0) |
| At school in person | 240 (12%) | 11.4 (6.0 - 14.0) | Hispanic/Latino | 370 (19%) | 7.8 (3.0 - 10.0) |
| Not at school | 98 (5%) | 5.0 (3.0 - 6.0) | Black, NH | 226 (12%) | 6.3 (2.0 - 7.0) |
| Studied fully online | 11 (1%) | 4.7 (4.0 - 5.0) | Asian, NH | 111 (6%) | 6.3 (2.0 - 8.0) |
| Employed | 749 (39%) | 9.4 (3.0 - 11.0) | Other, NH | 107 (6%) | 7.0 (2.0 - 9.0) |
| At work in person | 438 (23%) | 13.0 (5.0 - 18.0) |  |  |  |
| Not at work | 141 (7%) | 4.8 (2.0 - 7.0) |  |  |  |
| Worked fully online | 170 (9%) | 4.0 (1.0 - 6.0) |  |  |  |
| Self-employed | 94 (5%) | 4.8 (2.0 - 7.0) | **Sex** |  |  |
| At work in person | 46 (2%) | 6.3 (2.0 - 9.0) | Female | 970 (50%) | 7.7 (2.0 - 10.0) |
| Not at work | 25 (1%) | 3.7 (1.0 - 5.0) | Male | 953 (49%) | 7.0 (2.0 - 9.0) |
| Worked fully online | 24 (1%) | 3.2 (1.0 - 4.0) |  |  |  |
| Student & (self-) employed | 87 (4%) | 10.7 (3.0 - 15.0) |  |  |  |
| At work/In school in person | 55 (3%) | 13.7 (4.0 - 21.0) |  |  |  |
| Not at work/in school | 24 (1%) | 6.0 (3.0 - 8.0) |  |  |  |
| Worked/studied fully online | 7 (0%) | 3.3 (2.0 - 4.0) |  |  |  |
| Other | 651 (34%) | 3.9 (1.0 - 5.0) |  |  |  |
| **Age Group (years)** |  |  | **Household Income** |  |  |
| 0-4 | 128 (7%) | 6.9 (3.0 - 9.0) | Less Than \$10,000 | 71 (4%) | 4.2 (1.0 - 6.0) |
| 5-9 | 112 (6%) | 10.1 (5.0 - 13.0) | \$10,000 to \$24,999 | 138 (7%) | 5.4 (1.0 - 7.0) |
| 10-14 | 107 (6%) | 8.7 (4.0 - 11.0) | \$25,000 to \$49,999 | 290 (15%) | 6.2 (2.0 - 7.0) |
| 15-19 | 129 (7%) | 10.3 (4.0 - 13.0) | \$50,000 to \$74,999 | 297 (15%) | 6.5 (2.0 - 9.0) |
| 20-24 | 113 (6%) | 7.7 (2.0 - 9.0) | \$75,000 to \$99,999 | 260 (13%) | 8.4 (3.0 - 10.0) |
| 25-29 | 120 (6%) | 6.0 (2.0 - 8.0) | \$100,000 to \$149,999 | 376 (19%) | 8.3 (3.0 - 10.0) |
| 30-34 | 120 (6%) | 8.2 (1.0 - 9.0) | \$150,000 or more | 498 (26%) | 8.3 (2.0 - 10.0) |
| 35-39 | 135 (7%) | 8.1 (3.0 - 9.0) |  |  |  |
| 40-44 | 148 (8%) | 7.7 (2.0 - 10.0) |  |  |  |
| 45-49 | 97 (5%) | 7.8 (2.0 - 10.0) |  |  |  |
| 50-54 | 113 (6%) | 7.4 (2.0 - 8.0) |  |  |  |
| 55-59 | 137 (7%) | 8.9 (2.0 - 11.0) |  |  |  |
| 60-64 | 139 (7%) | 5.2 (1.0 - 7.0) |  |  |  |
| 65-69 | 114 (6%) | 5.3 (2.0 - 6.0) |  |  |  |
| 70-74 | 100 (5%) | 4.5 (1.0 - 7.0) |  |  |  |
| 75-79 | 65 (3%) | 5.1 (1.0 - 6.0) |  |  |  |
| 80+ | 52 (3%) | 3.8 (1.0 - 5.0) |  |  |  |

Notes: <sup>1</sup> N represents the weighted number of observations, calculated as the sum of individual participant weights. Percentage in parentheses shows the group's share in the total sample size (i.e., the total sum of weights). <sup>2</sup> Mean number of contacts (IQR)

Students attending school in person reported more than double the mean number of contacts compared to students not at school or learning remotely (Table 1). Respondents who worked in person had an average of 3.3 and 2.7 times more total contacts than those working online or not at work, respectively. These differences are also observed for self-employed respondents, albeit to a smaller degree (Table 1). We found that the mean number of contacts increases with the household income before plateauing at an income of $75,000 or higher (for comparison, the US median household income in 2023 was $80,610^25^). Specifically, the mean number of contacts nearly doubled from 4.2 contacts per day (IQR: 1.0 – 6.0) for households with an income lower than $10,000 to 8.4 (IQR: 3.0 – 10.0) contacts for households with an income between $75,000 and $99,999 (Table 1).

Moreover, we found a decrease of 1.2 contacts per day during weekends compared to weekdays (Table S6). A similar decrease (1.2 contacts per day) was found for adults with underlying conditions. The number of contacts was comparable between May and October. Remarkable differences were observed between adult respondents employed in different industries and occupations, ranging from 5 (IQR: 2.0 – 7.0) contacts for people in arts, IT, mathematics, physical and other sciences to 17.4 (IQR: 5.0 – 33.0) for health diagnosing or treating practitioners and 19.2 (IQR: 6.0 – 29.0) for individuals in education and training. We observed relatively small differences between residents of different regions, ranging from 6.9 (IQR: 2.0 – 8.0) contacts in the Northeast to 8.0 (IQR: 3.0 – 10.0) contacts in the Midwest. Differences for additional participant characteristics are reported in the Supplementary Material.

We conducted a multivariable statistical analysis using generalized additive models (GAM) to test the statistical significance of differences between population groups by age, race/ethnicity, sex, sick status, household size, household income, main activity, and mode of attendance (Fig. 1). The differences between online and in-person school/work attendance were statistically significant. In-person employed workers were identified as the group with the highest number of contacts compared to in-person students (relative ratio, RR=1.44, 95% CI: 1.20–1.74). Differences between NH White and Hispanic/Latino were not statistically significant. At the same time, NH Black and NH Asian had statistically significant fewer contacts than NH White (RR=0.87, 95% CI: 0.76 – 0.98 and RR=0.75, 95% CI: 0.63–0.89, respectively). After adjustment, female participants had statistically significant higher number of contacts than males (RR=1.21, 95% CI: 1.12–1.31). The increasing trend in the number of contacts by household income found in the descriptive analysis was confirmed, and statistically significant after controlling for covariates (Fig. 1). After adjusting for other covariates, we observed statistically significant non-linear marginal effects of age (see Supplementary Material). We also found no statistically significant variations in the number of contacts by survey week or by survey wave (Table S10). Being sick resulted in reporting around 10% fewer contacts (statistically significant before controlling for mode of attendance), while having a sick household member did not have a statistically significant association with the number of contacts. Additional results and sensitivity analyses with alternative regression specifications for total and non-household contacts are presented in the Supplementary Materials.

**Figure 1.**
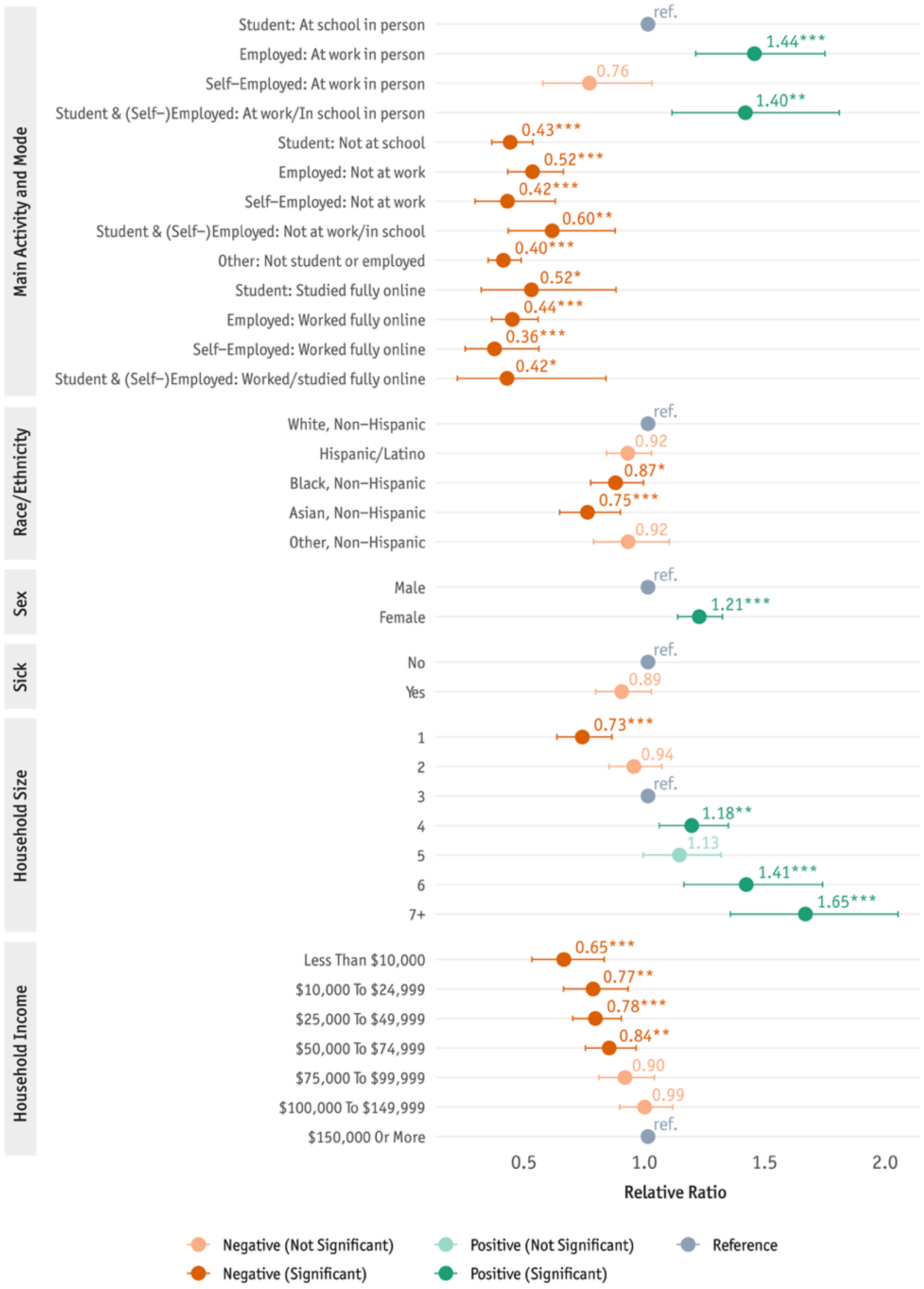
Relative ratios (RR) of the number of contacts by population group relative to NH White, male, non-sick, in-person students from a 3-person household with income of $150,000 or more. Error bars represent 95% CI. RRs are based on the GAM analysis (for more details see Table S10 in Supplementary Materials).

### Contact Patterns: Where and with Whom

Approximately 3.8 – 5.0 contacts were reported in school for school-age individuals (Fig. 2A). Working-age individuals reported 2.1-4.8 contacts at the workplace, with young workers (20-29 years old) reporting a lower number (∼3). The number of contacts in “other” settings (also referred to as community) ranged between 1.2 and 3.1, with a slight increase for older adults.

**Figure 2.**
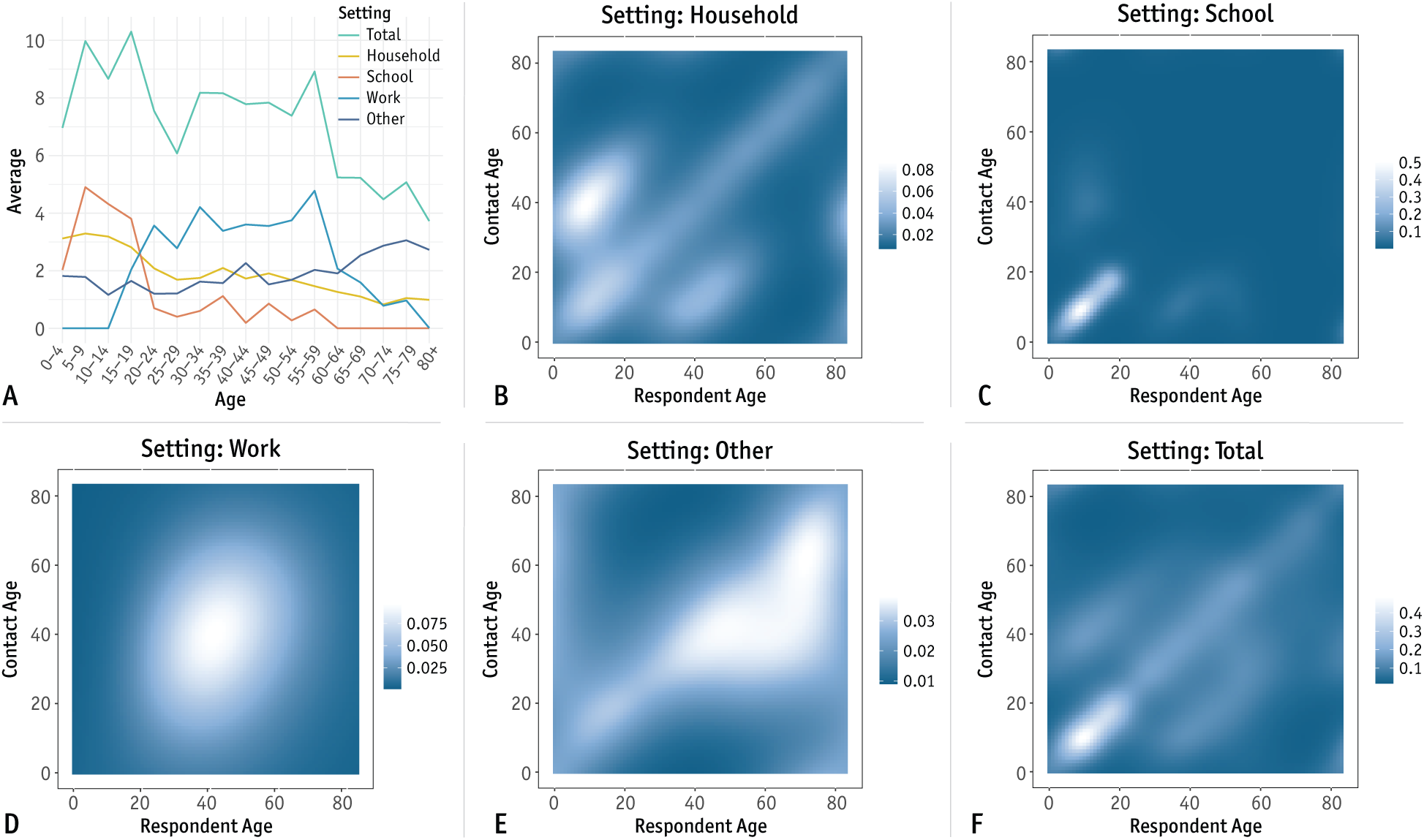
A. Average number of contacts per day by age group of the respondent in total and disaggregated by social setting. **B** Household contact matrix stratified by age. Each element of the matrix represents the estimated average number of contacts that a respondent of a given age has with individuals of other ages. The heatmap shows the results of a bivariate smoothing from a GAM model with tensor-product regression spline function of the respondent contact age (see Supplementary Material). **C** As B, but for contacts at school. **D** As B but for contacts at work. **E** As B but for contacts in other settings. **F** As B but for total contacts.

By analyzing the characteristics of each contact, we estimated not only where people have contacts, but also with whom they have contact (see Methods for details). As a coarse-grained representation, we estimated age-stratified contact matrices, where the entry *i*, *j* of the matrix represents the mean number of contacts that a respondent of age *i* has with individuals of age *j*. To characterize where people have contacts, these contact matrices were further disaggregated by social setting in which the contact took place (i.e., household, school, workplace, and community).

For household contacts, the contact matrix shows the classic three diagonals: the central diagonal representing contacts between spouses and siblings (main diagonal), the lower diagonal representing contacts between parents and children, and the upper diagonal representing contacts between children and parents (Fig. 2B). Due to the large number of contacts of students with same-age individuals in school setting, contact patterns in schools show a strong assortativity by age with the single highest number of contacts within the same age group taking place between 5-9 years old (5.0 contacts per day, Fig. S1B). The matrix also shows contacts taking place between students and school employees, but the average is relatively small as compared to the number of contacts between students (Fig. 2C). The workplace setting shows relatively homogenous contacts between work-age individuals (Fig. 2D) but still shows some level of assortativity by age (Table S4). Contacts in other social settings (e.g., community) exhibit strong assortativity by age (Fig. 2E). Moreover, individuals aged 40+ years have contacts skewed towards younger working age groups (Fig. 2E). When combining these setting-specific patterns, the total number of contacts shows assortativity by age, with the presence of the three diagonals found for the household setting, and the highest number of contacts being within school age groups (Fig. 2F). Additional age-stratified contact matrices are reported in the Supplementary Material, including frequency-based contact matrices, reciprocal contact matrices, contact matrices conditional on in-person presence of an individual in a specific setting, and contact matrices based on scenarios on in-person learning and work.

Similarly to age-stratified contact matrices, we estimated contact matrices stratified by race/ethnicity. We found high assortativity by race/ethnicity in all social settings, implying that individuals tend to interact more frequently with others in their own racial/ethnic group (Fig. S16A-E). The highest assortativity level is found in the household settings where the largest number of contacts occur within the same racial/ethnic group except for the highly heterogeneous (and small) NH Other group that have more contacts with the NH White group, corresponding with the largest population group with over 50% of the total population^26^ (Fig. S16B). We can observe similar patterns with only slightly lower levels of assortativity for other settings, including the community. Individuals who are more frequently reported as contacts relative to other off-diagonal groups are NH White.

Overall, NH White respondents reported the largest number of contacts in the workplace (mean: 2.7) and in the community (mean: 2.2), while Hispanic/Latino respondents had the largest number of contacts in the household (mean: 2.3, Fig. S16A). NH Other and Hispanic/Latino respondents reported the largest number of contacts in school (mean: 1.7 and 1.2, respectively), while NH Black and NH White respondents reported the lowest number of contacts at school (mean: 0.8) with small differences from NH Asian (mean: 0.9). It is important to note that these results represent the average number of contacts of the entire population of each racial/ethnic group, including those who do not work or study.

When analyzing contacts between males and females, we found high assortativity in all settings, except the household, where females have more contacts with males and vice versa (see Supplementary Material, Fig. S19-S21). We also observe notable differences in contact patterns by age between same-sex and opposite-sex interactions (Supplementary Material, Fig. S22 – S25). For example, males have more contacts at work after 70 years relative to females, even more so with other males, and higher contact assortativity by age is observed within sexes in school. All the presented results were consistent with the analyses performed using the unweighted sample and raw data (see Supplementary Material, Fig. S10-S15).

### Comparison with Pre-Pandemic Contact Matrices

Social contact patterns data in the US is limited, with no direct estimates of contacts in the absence of social distancing measures^7^. With no comparable pre-pandemic contact matrices for the US general population, we compared our results with pre-pandemic contact matrices inferred from synthetic populations or other indirect approaches^20,21^. We found a strong statistical association between our estimates and pre-pandemic estimates reported in Mistry et al.^20^ (Pearson’s *r* = 0.92, p-value < 0.001) and Prem et al.^21^ (Pearson’s *r* = 0.88, p-value < 0.001) (Fig. 3A). However, the absolute number of contacts is different: 7.4 contacts per day vs. approximately 12 contacts per day in both pre-pandemic studies^20,21^.

**Figure 3.**
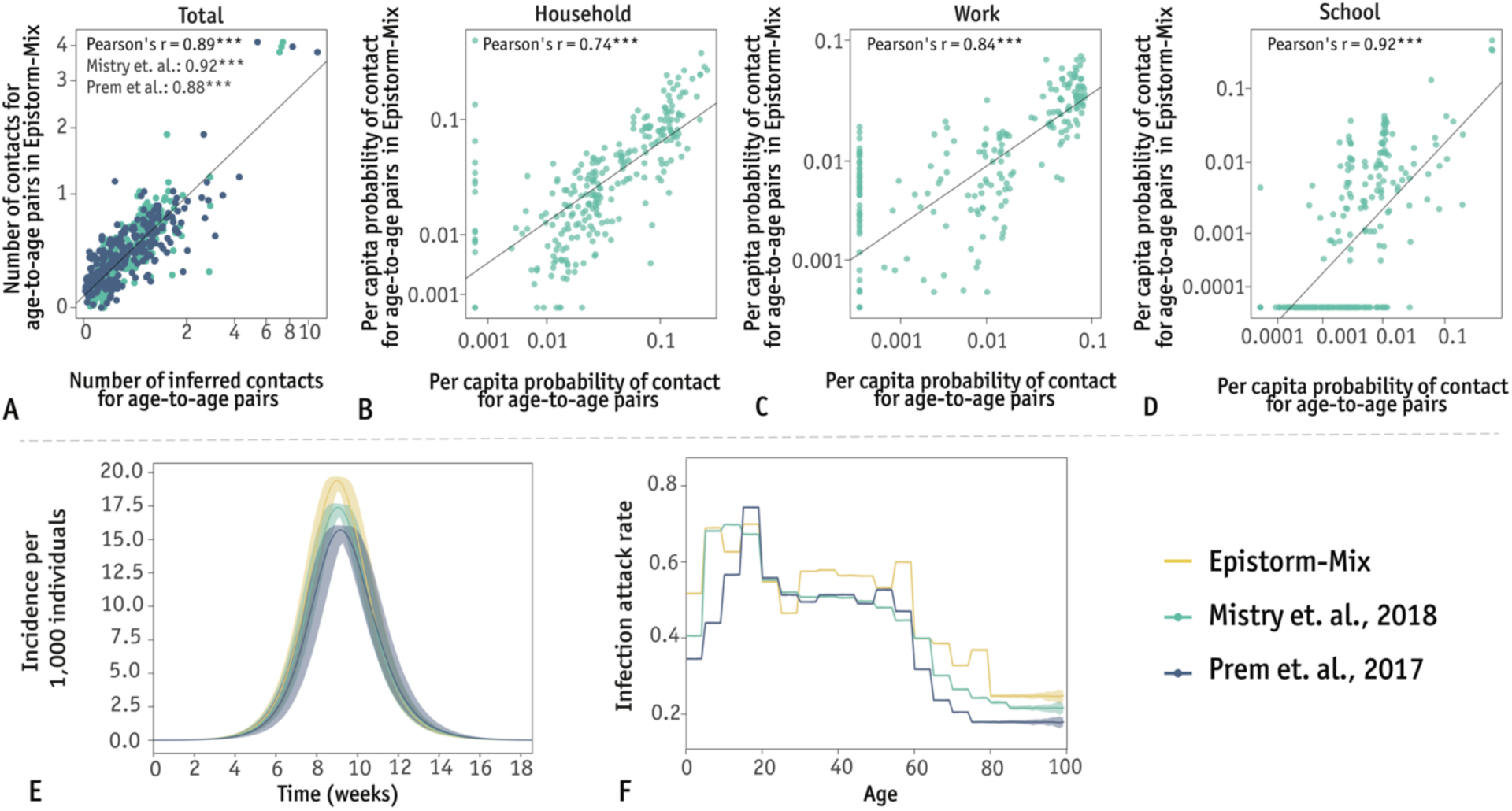
**A.** Correlation between elements of our age-stratified contact matrix, *M_ij_*, and the pre-pandemic contact matrices reported in Mistry et al.^20^ and Prem et al.^21^. Each point represents an element of the contact matrix. **B.** As A, but comparing the frequency-based household contact matrix estimated in this study with that reported in Mistry et al.^20^ (in log scale). **C.** As B, but for the school contact matrix. **D.** As B, but for the work contact matrix. **E.** Model-based estimates of the weekly incidence of new infections (mean and 95%CI) based different contact matrices for the US: Epistorm-Mix, Mistry et al.^20^ and Prem et al.^21^. In the simulations, we assumed a reproduction number of 1.5 and a generation time of 3 days. **F.** As E, but for model-based estimates of the final infection attack rate by age (mean and 95%CI).

For Mistry et al.^20^, we can further compare frequency-based contact matrices disaggregated by social setting (see Supplementary Material for the definition). Frequency-based, setting-specific contact matrices are relevant for mathematical modeling, as they can be combined using pathogen-appropriate weights for household, school, workplace, and community contacts to represent the mixing patterns most relevant to a given pathogen. Moreover, setting-specific weights can be determined to reflect interventions such as school and workplace closures; thus, the resulting aggregate matrix can be used to analyze the impact of interventions. We found a strong statistically significant correlation between Epistorm-Mix contact matrices and those reported in Mistry et al.^20^ for the household, school, and workplace settings (Fig. 3B-D). For the “other” (community) setting, we found a statistically significant, but negligible correlation (Pearson’s *r* = 0.04); this is related to the random mixing assumption in the community made in Mistry et al.^20^

By leveraging the estimated contact matrices, we also studied the effect of contact patterns on the epidemic dynamics of a respiratory pathogen. To this aim, we developed a stochastic age-structured susceptible-infectious-recovered (SIR) model where new infections in age group *i* depend on an age-dependent force of infection at time 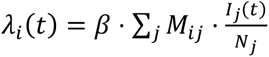, where *M_i,j_* represents the mean number of contacts of an individual of age *i* with individuals of age *j*, *β* represents the per-contact transmission risk, *I_j_*(*t*) represents the number of infectious individuals of age *j* at time *t*, and *N_j_* represents the number of individuals of age *j*. For a fair comparison across contact data sources, we assumed the same reproduction number (i.e., the average number of infections generated by a typical infector), generation time (i.e., average time between two consecutive generations of infections), and initial conditions (see Methods and Supplementary Material for details).

We found that the similarities between the pre-pandemic contact matrices and our post-pandemic one led to comparable predicted epidemic timing (Fig. 3E). However, we found smaller differences in the attack rate between school-age and working-age groups as compared to the results based on pre-pandemic contact matrices as well as higher attack rates among older adults (Fig. 3F).

## Discussion

The importance of understanding the key drivers of population exposure to airborne pathogens during the COVID-19 pandemic led to several studies estimating the number of contacts among the US adult population using comparable definitions of contact^13–18,27^. However, these estimates were obtained in 2020-2022, when physical distancing measures were still in place. Moreover, data for minors was not collected, thus providing only a fragmented picture of contact patterns during that period. These studies reported an average of 2 to 5 contacts per day^13–18,27^, although ∼10 daily contacts were reported for employees in long-term care facilities when working in-person^28^. In this work, we quantified the post-pandemic contact patterns for a nationally representative sample of the US population, estimating the heterogeneity in the number of contacts across different population groups and activity characteristics, and providing coarse-grained descriptions of contact patterns stratified by various population groups.

Our results for adults show that the number of contacts has increased relative to the pandemic period. Schools and workplaces are confirmed as major contact hubs potentially driving exposure patterns, making the dynamics of in-person vs. remote working and learning a public health relevant area of study. Our results show that remote/no attendance of school and work resulted in 2-3 times lower number of contacts than in-person students/workers. Remote working and learning also has an indirect effect on in-person students/workers by reducing their pool of potential contacts at school/work. This suggests a potential lasting redefinition of in-person social interactions since the pandemic. Moreover, despite showing multiple degrees of assortativity, we found that contacts were assortative by age and race/ethnicity in all settings, including the workplace and the community. Assortative mixing by sex was pervasive across settings, except for households. These findings suggest an underlying mechanism leading to in-group preference and less random mixing. This could impact pathogen transmission dynamics, potentially increasing the risk of infection within more assortative groups.

Echoing the rationale of this study, post-pandemic contact surveys with a similar contact definition were conducted in other countries, allowing us to make a cross-country comparison. Our estimate of the total number of contacts (7.4 contacts per day) aligns with estimates for European countries (6.0-9.9 contacts per day)^2,29^. Studies from Chinese provinces reported a larger number of daily contacts (11.5 and 12.0), primarily due to a significantly higher number of contacts in the school setting^30,31^ (see Supplementary Materials). Despite variations in absolute numbers, our age-stratified estimates of contact patterns in the US show strong and statistically significant correlations with those in all other countries. Moreover, these studies found a similar effect of in-person attendance on the number of reported contacts^29^. It is important to note that pre-pandemic estimates exist for those countries and are consistently higher than post-pandemic ones, similar to what we observed for the US (albeit based on indirect pre-pandemic estimates^20,21^). The differences and similarities with international estimates likely reflect a complex interplay of cultural differences, population structure, structural variations in educational and occupational environments, and varying societal adaptations to remote participation in learning and working.

While pre-pandemic synthetic matrices and international contact surveys provide valuable insights into age-based mixing, they often fail to capture other critical dimensions of contact heterogeneity, including demographic features and socioeconomic status. In this study, we found a positive correlation between the number of contacts and household income and heterogeneities by race/ethnicity, reflecting deeply rooted social, economic, and residential patterns that are specific to the United States. The Epistorm-Mix dataset is therefore a crucial resource for improving epidemiological modeling tailored to the US population.

The Epistorm-Mix study has several limitations, some of which are inherent to any survey-based research, including the potential for unmeasured error that cannot be entirely eliminated. However, multiple sources of bias were systematically addressed. Coverage error was mitigated through rigorous panel recruitment strategies; sampling error was minimized by employing probability-based recruitment and study-specific sample selection; and nonresponse error was reduced through targeted communication, incentive structures, and panel management protocols, including retention strategies. To address measurement error, survey instruments were evaluated and refined by Ipsos survey design specialists with attention to question flow, wording, and response formats. Finally, data processing and editing errors were minimized through comprehensive quality control procedures during data cleaning and processing. Additional methodological details are provided in the Supplementary Material. These strategies address different types of survey-related errors. Our findings may also be subject to recall bias. To minimize this bias, we instructed participants to record their contacts chronologically throughout the day and categorize them by the social setting in which they occurred. Additionally, in diary-based contact surveys, the participant burden always increases with the number of individually reported contacts. To alleviate this, multiple-choice options were prioritized over open-ended responses, and for professions with numerous work-related contacts (e.g., educators, healthcare professionals), these contacts were recorded as a group instead of individually. Our definition of a contact (in-person conversation of 5 or more words or physical contact) is widely adopted in the literature^5,23,24^ and relevant for many respiratory pathogens, but may not be adequate for pathogens that require a different type of contact to be transmitted. It is important to note that, while alternative data collection methods exist and may be less susceptible to some of these limitations, they also have their drawbacks. These drawbacks include construct validity issues related to analyzing indirect proxies of contacts (e.g., in mobility data and synthetic population studies), sampling biases such as no coverage for young children or hard-to-reach populations (e.g., in mobility data, sensor-based, and convenience sampling surveys), self-selection biases associated with opt-in requirements (e.g., in mobility data and convenience sampling surveys), limited or no recorded information about contacts (e.g., in mobility data and aggregated contact reporting studies).

Finally, our analysis focuses on the national level as the data sample is nationally representative. Our sample size does not allow for state-specific characterizations of contact patterns, but we provide statistics (and individual data) for the 9 US regions. Based on this data, future analysis can be performed to further characterize region-specific contact patterns. To incorporate socio-demographic heterogeneity and local context at the state- and location-specific levels, surveys with a sufficiently large sample size need to be conducted. Our survey covered two distinct periods of the year (Fall and Spring), and we did not find statistically significant differences in the number of contacts between the two periods. However, the survey does not provide information on contact patterns in the other two seasons, which may be more susceptible to fluctuations due to the school calendar and holidays.

In conclusion, Epistorm-Mix provides a nationally representative characterization of post-pandemic social contact patterns in the US. The comprehensive open-access dataset generated in this study, detailing contact numbers, sociodemographic characteristics of contacts, and interaction settings, provides the backbone to support further epidemiological modeling, research into transmission-related social and behavioral science, and public health planning. This data and analysis will serve as a data-driven integration of complex human interactions, population heterogeneity, and human social and economic behavior into the study of infectious disease dynamics, ultimately enhancing public health preparedness and response planning. To facilitate further analysis and application, all data products generated in Epistorm-Mix, including detailed contact matrices stratified by age, setting, and demographic subgroups, are publicly available. These resources will be instrumental for modeling and policy planning efforts aimed at improving infectious disease preparedness and response.

## Methods

### Survey administration and study population

The survey was administered to an online panel of respondents (KnowledgePanel^37^) established and maintained by Ipsos, which is widely used in health-related surveys, including those conducted by academics and the Centers for Disease Control and Prevention (CDC)^38–42^. The Panel relies on probability-based sampling methods for recruitment, providing a sampling frame that is representative of the US by age, sex, race/ethnicity, household income, geographic census region, and language for the Hispanic population^43^.

Academic calendars in the US vary between school districts, with the total number of school days per year typically ranging from 180 to 200. Therefore, most months include at least one period with school not in session. Our survey covered two periods of the year: the first survey wave was conducted in late Spring (May 15 - 31, 2024), which included the end of the academic year in some schools and Memorial Day federal holiday^44^; the second wave was conducted in Fall (October 8 - 23, 2024) when all schools were in session, except for Columbus Day and Yom Kippur^44,45^.

The study population consisted of non-institutionalized individuals aged 0 and above residing in the US. For 0-8-year-old participants, we employed guardian proxy reporting where parents/guardians completed the survey on behalf of their child. For 9-12-year-old participants, we used assisted self-reporting, where they were guided and aided by their parents/guardians. For participants aged 13-17 years, we used self-reporting, where they completed the survey independently after obtaining their parent/guardian’s consent. For all participants aged 18 and above, we used self-reporting. All self-reporting minors provided assent after their parent/guardian provided consent, while all adult survey-takers provided consent. The median completion time of the main survey was 10 minutes for Wave 1 and 11 minutes for Wave 2. Details on participant recruitment and survey administration are included in the Supplementary Material.

By using an established survey panel, we aim to increase the quality of data by limiting the issues typically associated with online surveys (e.g., issues of self-selection and broader sample unrepresentativeness in opt-in surveys, intentional misreporting, and low response quality)^43^ and by obtaining high cooperation rates across all population groups characteristic for KnowledgePanel^43^. In fact, the unweighted age distribution of the respondents in the sample was within 1 percentage point from population level statistics. Non-Hispanic White individuals represented more than 50% of the unweighted sample, with other racial groups falling within 3 percentage points of the population-level statistics.

The survey was approved by the Institutional Review Board at Indiana University (IRB #21846).

### Survey design

In the survey questionnaire, participants were first asked to contextualize the previous day (“yesterday”), including whether they were sick on that day, whether they worked or attended school, and their mode of attendance (in-person or remote). They were then asked to record the list of contacts they had during that day in an online contact diary. To facilitate respondents’ recall, the contact diary was divided into four sections based on the setting of the contact: home, school, work, or “other” (community). For each setting, they were asked to list all people with whom they had contact during the previous day. Then, for each contact, they were asked to provide the age of these contacts and choose the sociodemographic category that best described them (i.e., race/ethnicity, sex) and their relationship (e.g., person they live with, classmate/teacher, relative, friend). The questionnaire was offered in English and Spanish, and participants could choose to take the survey in their preferred language (Supplementary Materials, Appendix A and B).

Reporting large numbers of contacts places a significant burden on respondents, leading to decreased cooperation and underreporting of contacts. To mitigate this issue, we followed a common approach in the literature^5,47,48^ by encouraging employed individuals to report the number of numerous work-related contacts (30 contacts or more, further referred to as ‘numerous contacts’) rather than listing them individually. For individuals working in jobs with a high number (10 or more) of daily contacts with minors (e.g., educators, pediatricians), we also provided the opportunity to supply the total number of such contacts by age group instead of listing them individually.

In addition to the questionnaire, Ipsos provided a set of sociodemographic variables for the adult participants (see Supplementary Materials). As KnowledgePanel Core Profile Survey gathers information only for adults (18+), these variables align with the parent or guardian who was initially contacted to enroll participating children. Thus, additional questions on the age, race, and ethnicity of children were added to the survey questionnaires for children to ensure sample frame representativeness.

### Data Analysis

To account for sampling design, data for each wave were weighted using sample weights provided by Ipsos, which were calculated based on the 2022 American Community Survey estimates to ensure demographic representativeness by: age by sex, age by race/ethnicity, Census region, age and education level, household income, and language dominance within the Hispanic population. The two study waves were combined using a uniform weighting strategy, ensuring that each wave represented exactly half of the sample, thus mitigating potential bias from differential wave contributions. Details on data cleaning, data pre-processing, missing data imputations, and data analysis are reported in the Supplementary Material. Sensitivity analyses were performed using raw weighted data, weighted data after multiple imputations, and unweighted data after multiple imputations to assess the robustness of the results (Supplementary Materials). Analysis was performed in R, version 4.5.1 (2025-06-13)^49^. The list of used packages is provided in the Supplementary Materials and GitHub repository (see Data Availability).

To quantify heterogeneity in the number of contacts across respondent characteristics, we estimated the mean and 50% interquartile range (IQR) of the total number of contacts for each sociodemographic group. The mean is provided to be comparable to existing literature^2,5,6,16,18,19,23,24,27,29–31,47^ while IQR is provided to illustrate high level of individual variability in the reported number of contacts. The statistical significance of the differences in the number of contacts and rate ratios for individual characteristics was estimated using a set of multivariable negative binomial generalized additive models (GAMs)^50^. The negative binomial GAM approach was selected due to its flexibility to allow for potential non-linear effects of age and due to the overdispersion of contact count data. The best-fitting model was selected for the analysis based on the Akaike Information Criterion (AIC)^51^.

We provide coarse-grained representations of contact patterns in the form of contact matrices (*M_ij_*), where each cell represents the weighted mean number of contacts of a participant in group *i* (e.g., age, race/ethnicity) reported with individuals in group *j*. In particular,

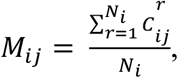

where *i* is the group of the participant, *j* is the group of the contact, *r* is the participant in group *i*, *C_ij_^r^* is the number of contacts reported by participant *r* in group *i* with a contact in group *j*, *N_i_* is the number of participants in age group *i*.

These matrices are stratified by age, by race/ethnicity, by sex, and by age and sex groups (Supplementary Materials). We also estimated frequency-based contact matrices (*F_ij_^r^*) that report the per capita probability of an individual of age *i* having a contact with individuals of age *j* in social setting *k*, following a procedure similar to Mistry et al.^20^ (see Supplementary Materials). Assortativity of contact matrices is estimated using the Q-index^22,33^.

We also developed a mathematical compartmental SIR model to estimate the effect of contact patterns on epidemic dynamics. In mathematical modeling, the contact matrix is a part of a multiplicative term that ultimately determines the force of infection that an individual of a specific population group is subject to during an epidemic. As such, the absolute number of contacts plays a less important role than the relative contact rates between population groups. Details on the compartmental model are reported in the Supplementary Materials.

## Data Availability

The cleaned (raw) data before imputations and the imputed data containing all variables used in this analysis are available on GitHub at https://github.com/epistorm/Epistorm-Mix together with the code to reproduce all our analyses.

## Acknowledgments

The authors would like to thank IPSOS, including Sergei Rodkin and Ying Wang, for their support with survey design and implementation.

