## Supplementary Material for "Epistorm-Mix: Mapping Social Contact Patterns for Respiratory Pathogen Spread in the Post-Pandemic United States"

Maria Litvinova[1,*], Shelly Sinclair[1], Allisandra G. Kummer[2], Paulo C. Ventura[2], Trevor Foster[1], Kayoko Shioda[3,4], M. Elizabeth Halloran[5,6], Alessandro Vespignani[7], Marco Ajelli[2,*]

[1] Department of Epidemiology and Biostatistics, Indiana University School of Public Health, Bloomington, IN, USA

[2] Laboratory for Computational Epidemiology and Public Health, Department of Epidemiology and Biostatistics, Indiana University School of Public Health, Bloomington, IN, USA

[3] Department of Global Health, Boston University School of Public Health, Boston, Massachusetts

[4] Center on Emerging Infectious Diseases, Boston University, Boston, Massachusetts

[5] Vaccine and Infectious Disease Division, Fred Hutchinson Cancer Center, Seattle, WA, USA.

[6] Department of Biostatistics, University of Washington, Seattle, WA, USA.

[7] Laboratory for the Modeling of Biological and Socio-technical Systems, Network Science Institute, Northeastern University, Boston, MA, USA

### Table of Contents


*1 Methods* ____ *4*

1.1 Survey Administration ____ 4

1.2 IRB Approval ____ 4

1.3 Ipsos KnowledgePanel ____ 5

1.4 Sample and Survey Weights ____ 6

1.5 Information on Survey Participants and Contacts ____ 8

1.5.1 Participant Data ____ 8

1.5.2 Contact Data ____ 13

1.6 Inclusion/Exclusion Criteria for Survey Participants ____ 15

1.7 Contact Data Organization and Cleaning ____ 16

1.8 Analyzed Variables, Data Codebook, and Code Availability ____ 16

1.9 Missing Contact Data Imputations ____ 19

1.10 Associations between Number of Contacts and Respondent Characteristics ____ 20

1.11 Frequency-Based Contact Matrices ____ 21

1.12 Conditional Contact Matrices ____ 22

1.13 Symmetrized Contact Matrices ____ 23

1.14 Epidemic Modeling ____ 23

1.15 Assortativity ____ 24

*2 Additional Results* ____ *24*

2.1 Descriptive Analysis of Total Contacts ____ 24

2.2 Association between Respondent Characteristics and Number of Contacts ____ 27

2.2.1 Univariable Analysis ____ 27

2.2.2 Interconnected Respondent Characteristics ____ 31

2.2.3 Multivariable Regression Analysis ____ 34

2.3 Descriptive Analysis of the Number of Non-Household Contacts ____ 39

2.4 Association Between Respondent Characteristics and Number of Non-Household Contacts ____ 43

2.5 Contact Matrices by Age ____ 49

2.5.1 Frequency-Based Contact Matrices ____ 50

2.5.2 Conditional Contact Matrices ____ 51

2.5.3 Symmetrized Contact Matrices ____ 52

2.5.4 Age-Stratified Contact Matrices for Alternative Scenarios: In-Person and Remote ____ 54

2.5.5 Sensitivity Analysis: Unweighted Sample Analysis ____ 55

2.5.6 Sensitivity Analysis: Raw Sample Analysis ____ 58

**2.6 Contact Matrices by Race/Ethnicity ____ 60**

2.6.1 Contacts by setting ____ 60

2.6.2 Conditional Contact Matrices ____ 61

**2.7 Contact Matrices by Sex ____ 62**

2.7.1 Contact Matrices by Age by Sex ____ 65

**2.8 Comparison With Pre-Pandemic Age-Stratified Contact Matrices ____ 69**

2.8.1 Pre- and Post-Pandemic ____ 69

2.8.2 Pre- and Post-Pandemic Frequency-Based Contact Matrices ____ 70

**2.9 Comparisons with Other Countries ____ 70**

***References ____ 73***

### 1 Methods

#### 1.1 Survey Administration

We conducted a two-wave cross-sectional survey that collected contact patterns data for a total of 1,930 individuals. The survey was administered and maintained by Ipsos, a widely used data supplier with experience in health-related surveys, having a history of being utilized by academics and the Centers for Disease Control and Prevention (CDC[1–5]. Survey participants were recruited from the Ipsos KnowledgePanel, consisting of more than 60,000 Panel members. Panel members were assigned to the survey based on the socio-demographic and geographic targets (see Section 1.4 “Sample and Survey Weights”) and KnowledgePanel weights (see Section 1.3 “Ipsos KnowledgePanel”). Ipsos invited one respondent from a representative sample of households to participate in this survey by sending a notification email letting them know that a new survey was available for completion. They were asked to complete it at their earliest convenience. The survey was administered to both English- and Spanish-speaking survey-takers

Upon completion, adult respondents received an entry into the Panel sweepstakes, and children 9-17 years old received a cash-equivalent incentive worth $5. After completing the survey once, respondents no longer had access to the study. Reminders were sent to non-responders on Day 3 of the field period. Additional reminders were sent to any remaining non-responders every three days. The field period was open until the socio-demographic and geographic targets (i.e., the number of participants in each population stratum) were met. After the survey was closed, the data were assigned study-specific weights and provided to Indiana University.

The survey questionnaire (also referred to as the survey instrument) was developed by the research team at Indiana University with input from Ipsos survey methodologists and survey specialists. Before each survey wave, we administered a pilot test of the survey to a small sample of participants ($N_1 = 25$, $N_2 = 24$). The first pilot was administered from March 26 to April 1, 2024, followed by Wave 1, which took place from May 15 to May 31, 2024. The second pilot was administered from October 1 to October 3, 2024, followed by Wave 2, which took place from October 8 to October 23, 2024. The pilot data were not included in the analysis but were used to identify potential difficulties for the participants or misunderstandings. The quality of the data and the feedback from the pilot participants were then utilized to evaluate and implement adjustments to the survey instrument, logic, and/or visualizations. Ipsos methodologists and survey specialists aided in all phases of the survey adjustments, after which Ipsos implemented necessary adjustments to the survey instrument to improve usability.

#### 1.2 IRB Approval

This study was approved by the Institutional Review Board (IRB) at Indiana University (IRB #21846). The initial approval of the study, including the review of informed consent, informed assent, recruitment materials, and the first draft of the survey instrument, occurred on March 7, 2024. After analyzing results from the first pilot test, the instrument was adjusted, and the amended protocol was approved on April 16, 2024. Before Wave 2, we improved the organization of the survey instrument and added two questions based on feedback from Wave 1. Additionally, we revised the informed assent and consent forms to include the CDC Certificate of Confidentiality after the Indiana University Human Research Protection Program Quality Assurance process. Approval from the IRB for the updated forms was obtained on September 30, 2024.

#### 1.3 Ipsos KnowledgePanel

Panel members were recruited into KnowledgePanel by Ipsos using a probability-based sampling procedure based on a national address-based sampling (ABS) methodology. ABS improves sampling by recruiting hard-to-reach individuals, such as non-Internet households and young adults[6]. Upon accepting the Panel invitation, Panel members complete a Core Profile Survey, which is used for Panel sampling and Panel weighting. The selection of survey study participants from Ipsos KnowledgePanel is based on the U.S. Census Bureau's American Community Survey (ACS) and Current Population Survey (CPS)[7,8]. KnowledgePanel includes the KnowledgePanel Latino members. KnowledgePanel Latino is developed to provide representative online samples for United States Hispanics, including English and Spanish-dominant Hispanics. It utilizes ABS as well as random digit dialing (RDD) sampling on census blocks with a Latino population density of 65% or higher.

Adults from sampled households are invited to join KnowledgePanel through a series of mailings, including an initial invitation letter, a reminder postcard, and a subsequent follow-up letter. Moreover, telephone refusal-conversion calls are made to nonresponding households for which a telephone number could be matched to a physical address. Invited households can join the panel by: (i) completing and mailing back a paper form in a postage-paid envelope; (ii) calling a toll-free hotline phone number maintained by Ipsos; (iii) going to a designated Ipsos website and completing the recruitment form online. During the initial recruitment of Panel members, all household members are enumerated. Following the enumeration, attempts are made to recruit every household member who is at least 13 years old to participate in KnowledgePanel surveys. For household members aged 13 to 17, consent is obtained from their parents or legal guardian during the initial recruitment interview. No direct communication with teenagers is attempted before obtaining parental consent. If a household does not have a computing device and/or internet access at home, they are given a tablet with a mobile data plan to complete panel surveys. Shipped devices are set up and supplied with instructions to simplify their use for responding to Panel surveys, with a specific technical support line available Monday through Friday. To assist Panel members with their survey taking, each individual has a personalized member portal listing all assigned surveys that have yet to be completed.

Once Panel members are recruited and profiled by completing the Core Profile Survey, they become eligible for selection for specific survey studies. Samples for such studies are based on a patented methodology[6] to ensure that the pool of Panel members is representative of the US population in terms of the following socio-demographic and geographic target dimensions:

- Sex: Male; Female
- Age group: 18-29; 30-44; 45-59; 60+
- Race/ethnicity: Non-Hispanic (NH) White; NH Black; NH Other or 2+ Races; Hispanic/Latino
- Education: Less than High School; High School Diploma or GED; Some College or Associates Degree; Bachelor's degree or higher
- Census Region: Northeast; Midwest; South; West
- Household income: $0-9,999; $10k-24,999; $25k-49,999; $50k-74,999; $75k-99,999; $100k-149,999; $150k+
- Home ownership status: Own; Rent/other
- Household size: 1; 2; 3; 4+
- Metropolitan Area: Yes; No

- Hispanic/Latino Origin: Mexican; Puerto Rican; Cuban; Other; NH
- Language Dominance: NH and English Dominant; Bilingual; Hispanic/Latino and Spanish Dominant

Based on the data from the Census ACS and CPS for the socio-demographic and geographic dimensions above, each Panel member is assigned the panel-specific design weight. Using these weights as measures of size, Ipsos typically utilizes a probability-proportional-to-size procedure to select study-specific samples. Still, study-specific procedures may differ (see Section S1.4 "Sample and Survey Weights").

#### 1.4 Sample and Survey Weights

This study aimed to determine the contact patterns of individuals of all ages (0 years and above). Therefore, non-institutionalized panelists aged 0 and older residing in the United States were invited to participate in this study. Two sample frames were used to ensure the sample was representative of the US population of all ages: the adult population (18+) and the parents/guardians of individuals 0-17 years old. Recruitment of panel members from the adult population frame proceeded according to the typical KnowledgePanel recruitment procedure. For parents/guardians, adults who reported having a child of 0-17 living with them during the Core Profile Survey were invited to the study and one of the children from the enumerated household was selected randomly and the parent/guardian was asked to either fill the survey on behalf of their child 0-8 years old, fill it in together with their child 9-12 years old, or send the invitation to their child 13-17 years old for independent completion.

The sample for this study was selected using the typical KnowledgePanel procedure, with the exception of minor oversampling of smaller strata by age and race/ethnicity (e.g., NH Asian). Table S 1 shows the targets for oversampling. After obtaining the data for each wave, the study-specific weights were computed by Ipsos via a raking procedure following three steps:

- Step 1: Design weights for the KnowledgePanel (KP) adult and parent samples were computed to reflect the probability of selecting participants' households. Design weights from the parent sample were multiplied by the number of children (0-17 years old) in the household to account for the probability of selecting one child. These weights served as design weights for children (0- 17 years old). The adult and child final design weights for participants were then combined and adjusted to reflect the correct proportion of individuals of participants aged 0-17 and 18+ within the total participants.
- Step 2: The combined design weights from Step 1 were adjusted to the following geodemographic distributions of the U.S. population (0+ years old) using an iterative proportional fitting (raking) procedure. The following benchmarks were obtained from the 2022 ACS[9] :
    - Age (0-14; 15-24; 25-44; 45-64; 65+) by sex (Male; Female)
    - Age (0-14; 15-24; 25-44; 45-64; 65+) by race/ethnicity (Hispanic/Latino; NH Asian; NH Black; NH Other; NH White)
    - Census Region (Northeast; Midwest; South; West)
    - Age and Education level (0-17 years; 18+ years with less than a high school education; 18+ years with a high school education; 18+ years with some college; 18+ years with a Bachelor degree or higher)

- Household Income ($0-$24,999; $25k-$49,999; $50k-$74,999; $75k-$99,999; $100k-$149,999; $150k+)
- Language Dominance within the Hispanic population for individuals ages 5+ years (English Dominant Hispanic/Latino, Bilingual Hispanic/Latino, Spanish Dominant Hispanic/Latino, All Others)

- Step 3: The computed weights were examined to identify and trim outliers at the extreme upper and lower tails of the weight distributions (1% - 99%) to avoid the creation of extreme weights. The final weights were scaled to add up to the total number of respondents and labeled as "*weight*".

The total design effect was estimated to be 1.08 for Wave 1 and 1.05 for Wave 2. The weights ranged from 0.329 to 1.945, with the mean weight being 1.

**Table S 1.** Achieved (unweighted) sample size by age and race/ethnicity.

| Age Group | Race/ Ethnicity | US Population | Wave 1 | | Wave 2 | |
|---|---|---|---|---|---|---|
| | | | N | % | N | % |
| 0 - 14 | Hispanic | 4.74% | 42 | 4.4% | 51 | 5.2% |
| | NH Asian | 0.88% | 16 | 1.7% | 16 | 1.6% |
| | NH Black | 2.33% | 24 | 2.5% | 32 | 3.3% |
| | NH Other | 1.70% | 15 | 1.6% | 23 | 2.3% |
| | NH White | 8.58% | 76 | 8.0% | 76 | 7.8% |
| | Total | 18.23% | 173 | 18.2% | 198 | 20.2% |
| 15 - 24 | Hispanic | 3.16% | 29 | 3.1% | 33 | 3.4% |
| | NH Asian | 0.60% | 12 | 1.3% | 10 | 1.0% |
| | NH Black | 1.70% | 24 | 2.5% | 17 | 1.7% |
| | NH Other | 0.88% | 10 | 1.1% | 12 | 1.2% |
| | NH White | 6.29% | 54 | 5.7% | 57 | 5.8% |
| | Total | 12.63% | 129 | 13.6% | 129 | 13.2% |
| 25 - 44 | Hispanic | 5.69% | 54 | 5.7% | 58 | 5.9% |
| | NH Asian | 1.88% | 20 | 2.1% | 21 | 2.1% |
| | NH Black | 3.34% | 34 | 3.6% | 41 | 4.2% |
| | NH Other | 1.54% | 18 | 1.9% | 12 | 1.2% |
| | NH White | 14.53% | 126 | 13.3% | 125 | 12.8% |
| | Total | 26.98% | 252 | 26.5% | 257 | 26.2% |
| 45 - 64 | Hispanic | 4.03% | 38 | 4.0% | 39 | 4.0% |
| | NH Asian | 1.50% | 21 | 2.2% | 23 | 2.3% |

| | | | | | | |
|---|---|---|---|---|---|---|
| | NH Black | 2.84% | 34 | 3.6% | 28 | 2.9% |
| | NH Other | 1.10% | 11 | 1.2% | 14 | 1.4% |
| | NH White | 15.55% | 129 | 13.6% | 130 | 13.3% |
| | Total | 25.02% | 233 | 24.5% | 234 | 23.9% |
| 65+ | Hispanic | 1.58% | 17 | 1.8% | 16 | 1.6% |
| | NH Asian | 0.83% | 13 | 1.4% | 11 | 1.1% |
| | NH Black | 1.55% | 19 | 2.0% | 18 | 1.8% |
| | NH Other | 0.52% | 13 | 1.4% | 11 | 1.1% |
| | NH White | 12.76% | 101 | 10.6% | 106 | 10.8% |
| | Total | 17.24% | 163 | 17.2% | 162 | 16.5% |
| **Total** | | 100% | **950** | 100.00% | **980** | 100.00% |

#### 1.5 Information on Survey Participants and Contacts

##### *1.5.1 Participant Data*

Obtained and analyzed information on survey participants came from two primary sources: questions from the current survey (CSQ) and information from the Ipsos Core Profile Survey (CPRS) filled in by adult Panel members. The CPRS information was available for adult participants (18 years and older) and the parents/guardians of the children (0-17 years old), also referred to as reference adults. As this would result in a lack of information on a range of socio-demographic characteristics for participating children, additional questions on age, race, ethnicity, language preferences, and proficiency were added to the survey questionnaires to ensure sample frame representativeness. Participants were also asked to provide information about their employment status, school enrollment and attendance, and whether they have functional limitations, were sick, or had a sick household member. Table S 2 presents the information on survey participants used for this analysis, derived from CPRS variables and collected through this survey (survey instrument shown in Appendices A and B).

**Table S 2.** Variables describing the survey participants.

| Variable | Values/Levels | 0-8 years | 9-17 years | 18+ years | Source |
|---|---|---|---|---|---|
| Age | Actual age in years | ✓ | ✓ | ✓ | CPRS/ CSQ |
| Sex | Male<br>Female | ✓ | ✓ | ✓ | CPRS/ CSQ |
| Sample Frame | Adults<br>Parents/guardians of respondents 0-17 years old | ✓ | ✓ | ✓ | CPRS |
| Spanish, Hispanic or Latino* | No, he is/she is/they are not<br>Yes, Mexican, Mexican - American, Chicano<br>Yes, Puerto Rican<br>Yes, Cuban, Cuban American | ✓ | ✓ | | CSQ |

| Variable | Values/Levels | 0-8 years | 9-17 years | 18+ years | Source |
|---|---|---|---|---|---|
| | Yes, other Spanish, Hispanic or Latino group | | | | |
| Detailed Race* | White<br>Black or African American<br>American Indian or Alaska Native<br>Asian<br>Native Hawaiian or other Pacific Islander<br>Some other race | ✓ | ✓ | ✓ | CPRS/ CSQ |
| Race/Ethnicity* | Hispanic/Latino<br>White, NH<br>Black, NH<br>2+ Races, NH<br>Other, NH | | | ✓ | CPRS |
| Hispanic/Latino Group Identification* | Mexican, Mexican - American, Chicano<br>Puerto Rican<br>Cuban, Cuban American<br>Other Spanish, Hispanic, or Latino group | ✓ | ✓ | | CSQ |
| Hispanic Census Subgroup* | NH<br>Mexican, Mexican American, Chicano<br>Puerto Rican<br>Cuban, Cuban - American<br>Other Spanish, Hispanic or Latino group | ✓ | ✓ | | CSQ |
| Asian Subgroup Identification* | Asian Indian<br>Chinese<br>Filipino<br>Japanese<br>Korean<br>Vietnamese<br>Other Asian | ✓ | ✓ | | CSQ |
| Native Hawaiian or Other Pacific Islander Group Identification* | Native Hawaiian<br>Guamanian or Chamorro<br>Samoan<br>Other Pacific Islander | ✓ | ✓ | | CSQ |
| English speaking* | Yes<br>No | ✓ | ✓ | | CSQ |
| English proficiency* | Very well<br>Well<br>Not well<br>Not at all | ✓ | ✓ | | CSQ |
| Other Language(s)* | Spanish<br>Chinese (any dialect)<br>Tagalog<br>Vietnamese<br>French<br>German<br>Another Language | ✓ | ✓ | | CSQ |
| Education, 5 categories | No high school diploma or GED | | | ✓ | CPRS |

| Variable | Values/Levels | 0-8 years | 9-17 years | 18+ years | Source |
|---|---|---|---|---|---|
| | High school graduate (high school diploma or the equivalent GED)<br>Some college or Associate degree<br>Bachelor’s degree<br>Master’s degree or higher | | | | |
| English proficiency* | Very well<br>Well<br>Not well<br>Not at all | ✓ | ✓ | | CSQ |
| Household Income | Less than $10,000<br>$10,000 to $24,999<br>$25,000 to $49,999<br>$50,000 to $74,999<br>$75,000 to $99,999<br>$100,000 to $149,999<br>$150,000 or more | ✓🖂🖂 | ✓🖂🖂 | ✓ | CPRS |
| Household Size | 1 – 10+ | ✓🖂🖂 | ✓🖂🖂 | ✓ | CPRS |
| ZIP code* | 01107 – 99623,<br>99999 = Unknown | ✓🖂🖂 | ✓🖂🖂 | ✓ | CPRS |
| State* | State of residence | ✓🖂🖂 | ✓🖂🖂 | ✓ | CPRS |
| Region, 4 categories | Midwest<br>Northeast<br>South<br>West | ✓🖂🖂 | ✓🖂🖂 | ✓ | CPRS |
| Region, 9 categories | South Atlantic<br>West South Central<br>West North Central<br>Middle Atlantic<br>East South Central<br>Pacific<br>East North Central<br>Mountain<br>New England | ✓🖂🖂 | ✓🖂🖂 | ✓ | CPRS |
| Sick Participant | Yes<br>No | ✓ | ✓ | ✓ | CSQ |
| Sick Household Member | Yes<br>No | ✓ | ✓ | ✓ | CSQ |
| School Enrollment (0-8) | Enrolled in school<br>Home schooled<br>Enrolled in day care<br>Other | ✓ | | | CSQ |
| School Attendance (0-8) | Yes, in person<br>Yes, online<br>No | ✓ | | | CSQ |
| Student | Yes<br>No | | ✓ | ✓ | CSQ |
| School Enrollment (9+) | Enrolled in school<br>Home schooled | | ✓ | ✓ | CSQ |

| Variable | Values/Levels | 0-8 years | 9-17 years | 18+ years | Source |
|---|---|---|---|---|---|
| | Other | | | | |
| Employed | Yes<br>No | | ✓ | ✓ | CSQ |
| Self-Employed | Yes<br>No | | ✓ | ✓ | CSQ |
| In-Person work/school attendance (9+) | Yes<br>No | | ✓ | ✓ | CSQ |
| Online work/school attendance (9+) | Yes<br>No | | ✓ | ✓ | CSQ |
| Industry* | Accommodation and Food Services<br>Administrative and Support Services (such As Call Centers, Security, Landscaping, and Janitorial)<br>Armed Forces<br>Arts, Entertainment, and Recreation<br>Child Day Care Services<br>Community/Non-Profit Organizations (including Religious and Political Organizations)<br>Construction and Specialty Contractors (such As Plumbing and Electrical)<br>Delivery Services, Warehousing, and Transportation (including Air, Rail, Water, Truck, and Passenger)<br>Education and Tutoring<br>Factory, Manufacturing, and Woodworking<br>Farming/Agriculture, Forestry, Fishing and Hunting, and Animal Production<br>Finance, Banking, and Insurance<br>Health Care (including Elder Care, Home Health Care)<br>Information (including Publishing, Media, Telecom, Internet Search, and Social Networking)<br>Management Of Companies and Enterprises<br>Mining, Quarrying, and Oil and Gas Extraction<br>Not Asked<br>Personal Services (including Beauty, Pet Care, and Household)<br>Professional, Scientific, Technical, and Business Services<br>Public Administration<br>Real Estate and Property Management<br>Refused<br>Repairs and Maintenance<br>Retail/Stores/Shopping (including Online Retail)<br>Utilities, Waste Management, and Remediation Services<br>Wholesale Trade | | | ✓ | CPRS |

| Variable | Values/Levels | 0-8 years | 9-17 years | 18+ years | Source |
|---|---|---|---|---|---|
| Occupation* | Architecture and Engineering<br>Armed Services<br>Arts, Design, Entertainment, Sports, and Media<br>Building and Grounds Cleaning and Maintenance<br>Business Operations (including Marketing)<br>Community and Social Services<br>Computer and Mathematical<br>Construction and Extraction<br>Education, Training, and Library<br>Farming, Forestry, and Fishing<br>Financial Operations or Financial Services (including Financial Advisor, Broker)<br>Food Preparation and Serving<br>Health Care Support (such As Nursing Aide, Orderly, Dental Assistant)<br>Health Diagnosing or Treating Practitioner (such As Physician, Nurse, Dentist, Veterinarian, Pharmacist)<br>Health Technologist or Technician (such As Paramedic, Lab Technician)<br>Installation, Maintenance, and Repair<br>Legal<br>Life, Physical, and Social Sciences<br>Management<br>Office and Administrative Support<br>Personal Care and Service<br>Precision Production (such As Machinist, Welder, Baker, Printer, Tailor)<br>Protective Service (such As Firefighter, Law Enforcement Worker)<br>Sales<br>Transportation and Material Moving<br>Other (Please Specify)<br>Refused | | | ✓ | CPRS |
| Underlying Conditions* | Asthma<br>Cancer or a malignancy of any kind<br>Diabetes or pre-diabetes<br>Heart attack, heart disease, or other heart condition<br>Hepatitis<br>HIV/AIDS<br>Inflammatory Bowel Disease (IBD) (such as Ulcerative colitis or Crohn's disease)<br>Weak or failing kidneys (Do not include kidney stones, bladder infections, or incontinence)<br>Lupus<br>Multiple sclerosis<br>Rheumatoid arthritis<br>Stroke | | | ✓ | CPRS |

| Variable | Values/Levels | 0-8 years | 9-17 years | 18+ years | Source |
|---|---|---|---|---|---|
| | No<br>Refused | | | | |
| Survey Feedback (Do you have any other comments about this survey?) | [Text] | ✓*** | ✓ | ✓ | CSQ |
| Date and Time of the Survey (Start and End) | [Date/Time]<br>Format: %Y-%m-%d %H:%M-%S | ✓ | ✓ | ✓ | CSQ |

* The information is excluded from the publicly shared dataset as only either it is not used or is aggregated (see Table S 5).
** Household variables reported by the reference adults from CPRS are assumed to characterize the household of the participants 0-17 years old.
*** The feedback is provided by the parent of the child 0-8 years old

Additionally, self-reported information on the reference adult occupation and industry was provided by Ipsos based on CPRF. For this analysis, we use the occupation and industry for adult (18+) participants who self-reported their contacts (see Table S 2). We also obtained information on the presence of 12 underlying conditions (diabetes, heart attack or other heart conditions, asthma, cancer, hepatitis C, HIV, inflammatory bowel disease, weak or failing kidneys, lupus, multiple sclerosis, rheumatoid arthritis, stroke) from Ipsos CPRF. These conditions were selected as they pose a higher risk of severe presentations of respiratory infections. Based on this information, a single variable was created to identify participants with underlying conditions. As other information obtained from CPRF, it is only available for adult participants or parents of participating children.

##### *1.5.2 Contact Data*

Once the participants provided their socio-demographics and contextualized their day, they were given the definition of a contact used in this study, a description of a contact diary, and age- and context-appropriate examples of how a contact diary would be filled for a hypothetical day (see Appendix A and B). They were also asked if they have a job requiring interactions with numerous people (30+) and children (10+). Individuals who reported having such jobs were instructed to include only the contacts of people they can characterize in the diary. They were informed that they will be able to provide the number of remaining contacts as a single value at the end of the contact diary.

Within the contact diary, participants were encouraged to recall their day, starting from the time they woke up until they went to sleep, when listing their contacts for each of the settings. In particular, participants were first asked if they had contacts (exchanged 5+ words or had physical contact) in a specific setting (with people who live with them - household, school, work, or anywhere else). Then, if they said they had contacts in that setting, they were asked to list people with whom they had contact. This design was created to improve participant focus on the specific setting, facilitate respondent recall, and minimize response burden. After the enumeration of contacts in all the settings, they were asked to provide information on each contact in a table prefilled with the previously provided contact names. The information included age, race/ethnicity, sex, and relationship with the contact (e.g., person they live with, classmate/teacher, relative, friend). All questions, except age, had predefined categories to choose from. After filling in this contact diary, individuals with jobs requiring numerous contacts with children (e.g., educators) were asked to provide the number of contacts for each of the age groups (0-2, 3-4, 5-9, 10-14, 15-17, 18+). In contrast, individuals with jobs requiring numerous contacts with other population groups were asked to provide the total number of remaining work-related contacts. The collected information is shown in Table S 3.

**Table S 3.** Collected information about contacts.

| Variable | Values/Levels |
|---|---|
| Name generator*<br>(Names, abbreviations, or other ways a participant refers to the contact) | [Text] |
| Social Setting | Household<br>School<br>Workplace<br>Other/Community |
| Relationship | Person [my child lives/I live] with<br>Classmate/teacher<br>Other schoolmate<br>Relative<br>Friend/acquaintance<br>Work colleague<br>Customer<br>Student<br>Other<br>Refused |
| Age (in years) | 0+ [Numeric] |
| Sex | Male<br>Female |
| Race/Ethnicity | White, NH<br>Hispanic/Latino<br>Black, NH<br>Asian, NH<br>Other, NH |
| Numerous Work-Related Contacts (More Than 30 People) | Yes<br>No |
| Number of Numerous Work-Related Contacts | 0+ [Numeric] |
| Numerous Work-Related Contacts (more than 10) with Children | Yes<br>No |
| Number of Numerous Work-Related Contacts | |
| With Students/Children Ages 0-2 | 0+ [Numeric] |
| With Students/Children Ages 3-4 | 0+ [Numeric] |
| With Students/Children Ages 5-9 | 0+ [Numeric] |
| With Students/Children Ages 10-14 | 0+ [Numeric] |
| With Students/Children Ages 15-17 | 0+ [Numeric] |
| With Students/Children Ages 18+ | 0+ [Numeric] |

* The information is excluded from the publicly shared dataset as only either it is not used or is aggregated (see Table S 5) .

#### 1.6 Inclusion/Exclusion Criteria for Survey Participants

We included responses from survey participants who:

(i) EITHER declared that they had no contacts during the reference day OR refused no more than 1 question on whether they had any contacts in a specific setting (i.e., household, school, work, other)

(ii) If they reported having contacts in at least one setting, they EITHER provided complete information on age, race/ethnicity, and sex for all listed contacts, if they listed fewer than 5 contacts, OR refused less than 1/3 of questions on each of the key contact characteristics (age, race/ethnicity, and sex) if they listed 5 or more contacts

(iii) If aged 0-8: did not refuse to self-identify as enrolled in school, enrolled in daycare, homeschooled, or other AND provided information on whether they attended school online, in-person, or were not at school, if they reported to be enrolled in school or daycare.

(iv) If aged 9+: did not refuse to self-identify as student, employed, self-employed, or other (i.e., not a student, employed or self-employed) AND provided information on whether they attended school/work online, in-person, or were not at school/work, if they identified as a student, employed or self-employed.

Ipsos performed the inclusion procedures with the inclusion logic above provided by the research team.

Additionally, we excluded respondents who: (i) selected the same (usually the first) category for all categorical contact variables, (ii) listed groups of contacts as a single contact (e.g., classmates, colleagues, people at the gym) without indicating how many contacts this group represented. We also excluded 2 participants who identified having contacts in a particular setting but reported only one contact with either "nobody "or "no one" listed in the name generator, due to the ambiguity of such a response. Finally, we excluded one participant who used only explicit language for their contact identifiers, and three participants with inconsistent responses (e.g., contacts with doctors, but all doctors were listed as 0 years old). We updated the respondents' weights after removing responses that met the exclusion criteria.

Survey response rate was about 51% for both waves (Table S 4). After applying the inclusion/exclusion criteria, survey data from 950 and 980 participants was analyzed for Wave 1 and Wave 2, respectively (Table S 4). The final qualification rate (number of qualified participants relative to the number of complete responses) was about 87% for both waves.

**Table S 4.** Included participant and response rate.

| | Total Fielded | Complete | Completion Rate | Included | Excluded | Qualified | Qualification Rate |
|---|---|---|---|---|---|---|---|
| Wave 1 | 2120 | 1087 | 51.3% | 963 | 13 | 950 | 87.4% |
| Wave 2 | 2186 | 1129 | 51.6% | 984 | 4 | 980 | 86.8% |

#### 1.7 Contact Data Organization and Cleaning

We separated respondent and contact variables from the respondent-level data provided by Ipsos to create easy-to-use datasets at both respondent and contact levels. To begin cleaning provided data, we verified that contacts belonged to the correct setting of the contact (as defined in the contact survey questionnaire). Some participants provided names and initials to identify each contact, while others provided descriptors (e.g., friend, classmate, mom). By classifying these descriptors into 12 groups (Table S 5), we created a contact identifier that can be further used in contact data quality controls and to allow sharing of this information while maintaining the privacy and de-identifiability of the data. Specifically, if the created identifier was not compatible with the setting definition, the contacts were moved out of the incorrectly selected setting. For instance, "Colleague" listed in the household setting was moved out and was either i) assigned to work or other setting based on a set of heuristic rules dependent on other respondent information; or ii) assigned to "Not in My Household" category and later imputed using multiple imputations (see Section S1.9).

We proceeded with data cleaning by removing duplicated contacts (e.g., child's parents reported in all settings rather than household only) based on the name generators and other provided information (e.g., same name and same or missing age, sex, race/ethnicity). A total of 250 duplicated contacts (2% of originally reported contacts) were eliminated from the data. Changes to the data were further manually reviewed by two independent researchers. Respondents who recorded many contacts at work or in the "other" setting tended to round the ages of their contacts, causing spikes in the contact age distribution. To mediate this, we used a smoothing spline regression curve to smooth these spikes in ages for contacts repeated within the same setting by the same participant. Lastly, we made manual adjustments based on respondents' comments (question QF1 in the instrument), as a few respondents indicated mistakes made while filling in the diary. All cleaning procedures resulted in a net removal of 173 contacts (1.4% of the originally reported contacts).

#### 1.8 Analyzed Variables, Data Codebook, and Code Availability

In addition to the provided information, we created aggregated groups of industries and occupations for adult respondents to balance data availability for a larger community with reducing potential privacy and identification concerns when the data is made publicly available. For the same reasons the name generator information was used to create a contact identifier that can be shared publicly. A categorical variable was also designed to characterize people at potentially higher risks of severe respiratory infections due to the presence of underlying conditions. Lastly, the categories of the respondents' race/ethnicity were aggregated to match the same categories as those of the contact. More details on the creation of these variables are available in Table S 5.

The final data is shared in the form of two datasets. The contact-level dataset includes information on the contact setting, relationship to the respondent, age, race/ethnicity, sex, participant ID, which can be used to match contact data with respondent data, and the respondent's weight. The participant-level dataset includes information on the participant ID, sample frame (adults or parents/guardians of respondents 0-17 years old), whether the participant was sick or had a sick household member, whether the participant had numerous contacts/educator contacts, sex, race/ethnicity, main activity (employed/student) combined with attendance mode (in-person/not in-person), geographic region, day of the week they completed the survey (both in US Central and participant local time zone), survey wave, and weight of the respondent. Respondents with zero contacts can be found in the respondent-level and contact-level datasets. For ease of identification, a binary variable was created to note if the participant had listed any contacts in the respondent-level dataset. For both respondent-level and contact-level data, we provide the raw dataset and the imputed dataset (see Section 1.9 "Missing Contact Data Imputations" below). Codebooks for respondent-level and contact-level data are provided with the data on GitHub at https://github.com/epistorm/Epistorm-Mix.

**Table S 5.** Description of created variables.

| Variable | Values/Levels | Procedure to create |
|---|---|---|
| Identifier | Names or Initials<br>Parent<br>Spouse<br>Kids<br>Friend<br>Relative<br>Colleague<br>Teacher<br>Student<br>Classmate<br>Other School<br>Customer<br>Service & Public Facing Locations<br>Multiple Contacts | To ensure the de-identification of the contacts, all name generators (Table S 3) were initially substituted with "Names or Initials" category. Word search was then used to classify contacts into other categories. For example, "husband", "wife", "esposa", "esposo", "significant other", "partner", "spouse", etc. were classified as "spouse".<br>All contacts added to the contact dataset based on "Number of Job-Related Numerous Contacts" variable (i.e. number of reported numerous contacts less than 30 – not satisfying the definition of numerous contacts provided to respondents) are assigned the identifier "Multiple contacts". |
| Race/Ethnicity (Respondents) | Hispanic/Latino<br>White, NH<br>Black, NH<br>Asian, NH<br>Other, NH | To harmonize the levels of race/ethnicity across respondents and contacts we utilized variables "Spanish, Hispanic or Latino", "Detailed Race", and "Race/Ethnicity" to create a single variable for respondents of all ages. "Asian, NH" is defined as respondents who identified as "Asian" but not as "Spanish, Hispanic or Latino". "American Indian or Alaska Native",<br>"Native Hawaiian or other Pacific Islander", and "2+ Races, NH" groups were included in "Other, NH". |
| Industry Group | Arts, Information, Finance, and Professional Services<br>Sales and Support Services<br>Education, Tutoring, and Child Care<br>Infrastructure, Logistics, Construction, and Maintenance<br>Health Care | Individual industries (Table S 2) were grouped to avoid categories with less than 20 observations for identifiability reasons. |

| | | |
|---|---|---|
| | Factory, Manufacturing, and Woodworking<br>Other<br>Not asked or refused<br>Minor | |
| Occupation Group | Arts, IT, Mathematical, and Physical, Life or Social Sciences<br>Building Cleaning, Maintenance and Repair<br>Business Operations<br>Construction, Architecture, Engineering and Precision Production<br>Education, Training, and Library<br>Financial Operations and Services<br>Food Preparation and Serving<br>Health Diagnosing or Treating Practitioner<br>Health Technicians and Care Support<br>Office and Administrative Support, Management and Legal<br>Sales and Personal Care Services<br>Transportation, Moving, Community and Social Services<br>Other<br>Not asked or refused<br>Minor | Individual occupations (Table S 2) were grouped to avoid categories with less than 20 observations for identifiability reasons. |
| Main Activity | Student<br>Employed<br>Self-Employed<br>Student & (Self-)Employed<br>Other | The following information from the survey (Table S 2) was used to create a single variable to categorize all the participants: School Enrollment (for participants 0-8 years old), Student, School Enrollment (for participants 9+ years old), Employed, Self-Employed. Participants 0-8 years old, who are enrolled in school or day care are categorized as "Student" to make the data more comparable with the data from the National Center for Education Statistics (NCES). All participants 9+ years old, are categorized based on the way they self-identify. For those who selected multiple categories: i) those self-identifying as students and either employed or self-employed were categorized as "Student & (Self-)Employed"; ii) those who identified as both employed and self-employed, are categorized as "Employed". Those who did |

| | | |
|---|---|---|
| | | not self-identified as either student, employed or self-employed are categorized as "Other". |
| Date and Time of the Survey (in the time zone of the respondent) | [Date/Time]<br>Format:<br>%Y-%m-%d %H:%M-%S | Participant zip code was used to assign appropriate time zone to the response. Based on the assigned time zone the local date and time was calculated based on the date and time provided by Ipsos (Chicago time zone for all respondents) |

For reproducibility all the code for data imputations and data analysis is provided on GitHub at https://github.com/epistorm/Epistorm-Mix. All data analysis was performed in R, version 4.5.1 (2025-06-13). We used the following packages in the analysis: "dplyr"[10], "survey"[11] , "tidyverse"[12], "ggplot2"[13], "flextable"[14], "gt"[15], "gtsummary"[16], "fitdistrplus"[17], "mgcv"[18–22], "sjPlot"[23], "patchwork"[24], "doParallel"[25], "extrafont"[26], "readxl"[27], "data.table"[28], "tidyr"[29], "psych"[30], "Hmisc"[31], "prettyGraphs"[32], and "missForest"[33].

##### 1.9 Missing Contact Data Imputations

All missing data were sorted into two categories: "Refused" (participants had an option to skip or refuse any questions they did not want to answer) and "Not in My Household" (contact setting and relationship lost due to misclassification of non-household contacts as household). To handle missing data characteristics due to non-response, we performed nonparametric multiple imputations for mixed-type data using random forest[33]. The random forest algorithm is used for iterative imputations, where newly imputed data is used to update the random forest prediction and perform imputation of the next variable. The iterations proceed until there are no more improvements to be made, namely, NRMSE stops decreasing, or maximum iterations are reached. The selected mixed-type data technique for random forest is specifically developed for the mixed-type high-dimensional data, like the contact data we analyze, and is robust to complex nonlinear interactions.

In addition to observed contact data, the following information on respondent characteristics were used for imputations: sample frame (adults or parents/guardians of respondents 0-17 years old), respondent age, race/ethnicity, sex, household income, size and geographical region of residence (at two levels of aggregation), respondent and household member health status (sick vs. not sick), numerous contacts job status (if the respondent is in a job with high number of contacts with individuals of any age or children), school enrollment, main activity (student, employed, self-employed, or other), industry and occupation groups, mode of school/work attendance (if any), and survey wave.

To best reflect the underlying correlations between contact characteristics, we imputed contact data in four steps. First, we imputed the contact's socio-demographics. This resulted in missing data ("Refused") on contact age, sex and race/ethnicity for 1.6% (187 out of 12,014), 0.9% (108 out of 12,014), and 1.1% (136 out of 12,014) of contacts, respectively, being imputed. Second, based on the complete contact socio-demographic characteristics, we imputed missing data on contact relationship and contact setting in the "Not in My Household" category (2.2% and 0.9% of contacts) that resulted from the cleaning procedures. Third, we followed with the imputation of the contact relationship in the "Refused" category (10.4% of contacts) to account for potential selfselection of respondents or correlation with contact characteristics when respondents refuse to provide the information on the relationship with the contact. This procedure enables us to distinguish among the three potential mechanisms of missingness: refusal due to respondent burden, data loss resulting from an incorrect choice of relationship/setting, and refusal to provide a relationship due to privacy concerns. At each step, the selected category being imputed was transformed into missing and imputed using an ad-hoc modification of the "missForest"[33] function from the "missForest"[33] R package. This allowed us to implement iterative fitting of a random forest model with a robust stop criterion (NRMSE) and account for a potential complex non-linear correlation with observed variables. All random forest inputs were set to default, including the maximum number of iterations (10) and tree size (100). The iterations were parallelized by forests using multiple computer cores for more time-efficient computation. The code for the utilized function and the imputation procedure is provided in the GitHub code directory.

After imputations, the contacts provided as numerous but not respecting the definition of numerous contacts (30 or more work-related contacts) were added to the data with the reported age group (if any). Among the 182 respondents reporting such contacts (reported as numerous but under 30 people, further referred to as multiple contacts), 72 self-identified as having a job requiring high contact rates with children (40 out of them were employed within the "Child Day Care, Education and Tutoring" industry group) and 110 self-identified as having a job requiring high contact rates with all/other age groups. In total, 2,026 contacts were added to the contact-level dataset as a result of this step, resulting in a 16.7% increase in the cumulative number of contacts.

The exact age, sex, and race/ethnicity of each contact were inferred using the following procedure. For "student" contacts reported by educators (i.e., respondents employed in the "Child Day Care, Education and Tutoring" industry group who self-identified as having a job with high contact rates with children), contact characteristics were sampled based on the distribution of population enrolled in school by age and race/ethnicity in the participant region of residence (9 categories – see Table S 2). Such distributions were extracted from pooled IPUMS ACS 2021-2023 1-year micro-level data[34]. The probability distribution for race/ethnicity conditional on the sampled age of the contact was applied with a scaling factor to incorporate assortativity by race/ethnicity of student-teacher contacts observed in the data. The scaling factor was calculated as the fraction of non-missing student contact information multiplied by the assortativity of the contacts between teachers of the participant's race/ethnicity and their students by race/ethnicity reported either by the students or by the teachers. Race/ethnicity distribution for missing data was sampled 10,000 times, and the sample best fitting the national race/ethnicity distribution of students was used.

For the respondents who are not educators, multiple contact age and race/ethnicity were inferred based on the US population distribution by age and race/ethnicity from IPUMS ACS 2021-2023 micro-level data, conditional on participant age group and participant-provided contact age group (if any). Sex was sampled using a balanced Bernoulli distribution.

#### 1.10 Associations between Number of Contacts and Respondent Characteristics

We performed a generalized additive model (GAM) analysis to evaluate the associations between total contacts and respondent characteristics. We first assessed the univariate associations between each covariate and the outcome, including an examination of different reference groups for categorical variables. As no smoothing was applied for categorical variables in this step, GAM automatically simplified into a generalized linear model (GLM). Subsequently, to identify the optimal multivariable model, we evaluated all possible combinations of covariates, including the numeric age for which spline smoothing was used. This process allowed us to estimate the associations for each covariate conditional on the others. For respondent age, the plate regression spline smoothing was used to maximize flexibility and computational efficiency. The same modeling procedure was then applied to analyze the relationship between total non-household contacts and the same set of covariates.

#### 1.11 Frequency-Based Contact Matrices

Based on the reported number of contacts by each individual, we calculated frequency-based contact matrices, following a procedure similar to the calculation of $F_{ij}^{k}$ in Mistry et al.[35]. To ensure a fair comparison, we first calculated the relative abundance of contacts between individuals of age *i* and individuals of age *j* reported by each participant *r* in setting *k*. This is the step that differs slightly, as $\Gamma_{ij}^{k(r)}$ in Mistry et al.[35] was calculated based on each configuration *s* of the setting *k* due to the synthetic nature of that procedure, while we based our calculation of $\Gamma^{k(r)}$ on each participant who reported having contacts in setting *k*. In particular:

$$\Gamma_{ij}^{r(k)} = \frac{C_{ij}^{r(k)}}{\sum_{r=1}^{N_{ik}} C_{ij}^{r(k)}},$$

where

- $i$ is the age group of the participant; $j$ is the age group of the contact.
- $r(k)$ is the participant who reported having the contact in the social setting.
- $C_{ij}^{r(k)}$ is the number of contacts reported by participant *r* of age group *I* with a contact of age group *j* in a social setting *k*.
- $N_{ik}$ is the number of participants in age group *I* who reported non-zero contacts in setting *k*.

Second, we calculate $F_{ij}^{k}$, i.e. the per capita probability of contact of an individual of age *i* with an individual of age *j* in setting *k, as*:

$$F_{ij}^{k} = \frac{\sum_r \Gamma_{ij}^{r(k)}}{N_i},$$

where $N_i$ is the total number of respondents of age group *i*.

By computing this matrix, we provide the modeling community with an easy and scalable way of incorporating setting-specific uniform changes in contact rates, for example in scenario analyses. In particular, in this study the average population-wide number of contacts:

$$\frac{1}{N}\sum_{i=1}^{A_i} N_i \sum_{j=1}^{A_j} M_{ij} = \frac{1}{N}\sum_{i=1}^{A_i} N_i \sum_{j=1}^{A_j}\sum_{k=1}^{A_k} M_{ij}^{k} =$$

$$\frac{1}{N}\sum_{i=1}^{A_i} N_i \sum_{j=1}^{A_j}\sum_{k=1}^{K} \omega_k F_{ij}^{k} = \frac{1}{N}\sum_{i=1}^{A_i} N_i \sum_{k=1}^{K} \omega_k \sum_{j=1}^{A_j} F_{ij}^{k} =$$

$$\frac{1}{N}\sum_{i=1}^{A_i} N_i \sum_{k=1}^{K} \omega_k \sum_{j=1}^{A_j} \frac{1}{N_i}\sum_{r=1}^{N_{ik}} \frac{C_{ij}^{r(k)}}{\sum_j C_{ij}^{r(k)}} =$$

$$\frac{1}{N}\sum_{k=1}^{K}\omega_k\sum_{i=1}^{A_i}N_{ik} = \frac{1}{N}\sum_{k=1}^{K}\omega_k N_k = \sum_{k=1}^{K}\omega_k\frac{N_k}{N}$$

- N is the total number of participants
- $A_i$ is the number of participant age groups *I* and $A_j$ is the number of contact age groups *j*.
- $K$ is the number of settings *k*
- $M_{ij}$ is the total contact matrix showing the average number of contacts reported between an individual in age group *I* and an individual in age group *j*.
- $M_{ij}^{k}$ is the setting-specific contact matrix showing the average number of contacts reported between an individual in age group *I* and an individual in age group *j* in a specific setting *k*.
- $N_{ik}$ is the number of participants in age group *I* who reported non-zero contacts in setting *k*.
- $N_k$ is the number of participants who reported non-zero contacts in setting *k* (referred to as $Z_k$ in Mistry et al.[35]).
- $\omega_k$ is a setting-specific scaling factor representing the average number of contacts in each setting *k* reported by participants who had at least one contact in setting k.

Based on the definition of $\Gamma^{r(k)}$ and the equation above, $\sum_{j=1}^{N} F_{ij}^{k}$ represents the per-capita probability of having non-zero contact in setting *k* given that a person belongs to age group *i*.

Additionally $\omega_k = \frac{1}{N_k}\sum_{i=1}^{A_i} N_{ik}\sum_{j=1}^{A_j}\Theta_{ij}^{k}$, where $\Theta_{ij}^{k}$ is the contact matrix representing the average number of contacts with contacts in age group *j* in setting *k* reported by participants in age group *I* with at least one contact in setting *k* (described below as "conditional contact matrices" and visualized in Figure S 6).

#### 1.12 Conditional Contact Matrices

We also computed the contact matrices representing the average number of reported contacts given that a respondent is present (has non-zero contacts) in a specific setting. In particular:

$$\Theta_{ij}^{k} = \frac{\sum_{r=1}^{N_{ik}} C_{ij}^{r(k)}}{N_{ik}},$$

where

- $C_{ij}^{r(k)}$ is the number of contacts reported by participant *r* of age group *I* with a contact of age group *j* in a social setting *k*.
- $N_{ik}$ is the number of participants in age group *I* who reported non-zero contacts in setting *k*.

These contact matrices represent the mixing patterns of individuals who have contacts in each setting, and, based on the definition of the average number of contacts above, the setting-specific scaling factor $\omega_k$ can be computed based on these matrices. In particular:

$$\sum_{k=1}^{K} \omega_k \frac{N_k}{N} = \frac{\sum_{k=1}^{K}\sum_{i=1}^{A_i}\sum_{j=1}^{A_j}\sum_{r=1}^{N_{ik}} C_{ij}^{r(k)}}{N} = \sum_{k=1}^{K} \frac{N_k}{N} \sum_{i=1}^{A_i}\sum_{j=1}^{A_j} \frac{N_{ik}}{N_k} \frac{\sum_{r=1}^{N_{ik}} C_{ij}^{r(k)}}{N_{ik}}, \text{ therefore}$$

$$\omega_k = \sum_{i=1}^{A_i} \frac{N_{ik}}{N_k} \sum_{j=1}^{A_j} \Theta_{ij}^{k}$$

#### 1.13 Symmetrized Contact Matrices

A symmetric contact matrix, in theory, implies that the number of contacts reported by group $I$ with group $j$ is equal to the number of contacts reported by group $j$ with group $i$. However, empirical contact matrices generated from social contact surveys typically provide asymmetric contact matrices, illustrating the number of contacts reported by each participant. These asymmetries reflect population-level socio-demographic mechanisms; for example, not all adults will have contact with children in their household. Still, most children will have contacts with adults in their households. For illustration purposes, we also show symmetrized contact matrices where each entry represents the number of reciprocal contacts between the two groups. In this study, the symmetrized matrices are computed as:

$$M_{ij}^{k} = \frac{M_{ij}^{k} + {M^{T}}_{ij}^{k}}{2}.$$

#### 1.14 Epidemic Modeling

We simulated the transmission of a generic respiratory pathogen using an age-structured stochastic Susceptible-Infectious-Removed (SIR) model. The following equations define the discrete-time evolution of the model:

$$S_i(t+\Delta t) = S_i(t) - \Delta I_i(t)$$
$$I_i(t+\Delta t) = I_i(t) + \Delta I_i(t) - \Delta R_i(t)$$
$$R_i(t+\Delta t) = R_i(t) + \Delta R_i(t)$$

where $S_i(t)$, $I_i(t)$ and $R_i(t)$ are the number of susceptible, infectious, and removed individuals of age $i$ at time $t$, respectively, and $\Delta t$ is the length of a time step. The quantities $\Delta I_i(t)$ and $\Delta R_i(t)$ are random variables defined as:

$$\Delta I_i(t) \sim \text{Binomial}(S_i(t),\ \lambda_i(t)\cdot\Delta t)$$
$$\Delta R_i(t) \sim \text{Binomial}(I_i(t),\ \gamma\cdot\Delta t)$$

where $\gamma$ is the inverse of the generation time and $\text{Binomial}(M, p)$ is a binomial random variable with $M$ trials and probability $p$ of success. The age-dependent function $\lambda_i(t)$ at time $t$ is given by:

$$\lambda_i(t) = \beta \cdot \sum_j M_{ij} \cdot \frac{I_j(t)}{N_j}$$

where $\beta$ is the per-contact pathogen transmission risk, $N_j$ is the total number of individuals of age $j$, and $M_{ij}$ is the contact matrix where each element represents the weighted mean number of contacts reported by an individual in a population subgroup $i$ with individuals in the subgroup $j$.

The basic reproduction number of this model is defined as $R_0 = \frac{\beta}{\gamma} \cdot \rho(M)$, where $\rho(M)$ is the spectral radius (i.e., largest eigenvalue in absolute value) of the contact matrix M[36].

Simulations are performed assuming $R_0 = 1.5$, and a generation time of 3 days as per influenza literature[37,38]. Simulations were initialized with $10^{-5} \cdot N$ initially infected individuals aged 45 years, while the rest of the population was set as susceptible to the infection.

All the model simulations and model output visualizations were performed in Python. The code is provided in the GitHub repository at https://github.com/epistorm/Epistorm-Mix.

##### 1.15 Assortativity

We quantified the degree of assortative mixing (i.e., the tendency for individuals to interact with others of similar characteristics, also known as homophily) by calculating the Q-index ($Q$), as defined by Iozzi et al.[39] and Fumanelli et al.[40]:

$$Q = (Tr(P) - 1)/(n - 1),$$

where:

- $n$ is the number of subgroups.
- $P$ is a normalized contact matrix, where each element $p_{ij}$ represents the proportion of contacts made by individuals in subgroup $i$ that are with individuals in subgroup $j$. This is calculated as: $p_{ij} = M_{ij}/\sum_{j=1}^{n} M_{ij}$. In simpler terms, we divide each cell in the original contact matrix by the total number of contacts reported by that group.
- $Tr(P)$ is the trace of the matrix $P$, representing the sum of the diagonal elements ($p_{ij}$i.e., the sum of the proportions of contacts that individuals within each group have with others in the same group.

A higher Q-index indicates greater assortativity, suggesting that contacts are more frequent within groups than between groups. A negative Q-index would indicate that the contact happens more often between groups than within groups, while a Q-index of 0 would indicate random mixing.

#### 2 Additional Results

##### 2.1 Descriptive Analysis of Total Contacts

As illustrated in Table S 6, the distribution of the unweighted sample size is very similar to the distribution of weighted sample size. The differences in the two distributions range between 0 and 4 percentage points for most characteristics, with the exception of race/ethnicity, where smaller population groups were intentionally oversampled, decreasing the sample for the NH White population by 7 percentage points.

**Table S 6.** Descriptive statistics.

| Characteristic | N obs[1] | Weighted N[2] | Number of total contacts[3] |
|---|---|---|---|
| **Total Population** | 1,930 (100%) | 1,930 (100%) | 7.4 (2.0 – 9.0) |
| **Main Activity & Attendance Mode** | | | |
| Student | 383 (20%) | 349 (18%) | 9.4 (4.0 – 12.0) |

| | | | |
|---|---|---|---|
| At school in person | 271 (14%) | 240 (12%) | 11.4 (6.0 – 14.0) |
| Not at school | 100 (5%) | 98 (5%) | 5.0 (3.0 – 6.0) |
| Studied fully online | 12 (1%) | 11 (1%) | 4.7 (4.0 – 5.0) |
| Employed | 741 (38%) | 749 (39%) | 9.4 (3.0 – 11.0) |
| At work in person | 428 (22%) | 438 (23%) | 13.0 (5.0 – 18.0) |
| Not at work | 136 (7%) | 141 (7%) | 4.8 (2.0 – 7.0) |
| Worked fully online | 177 (9%) | 170 (9%) | 4.0 (1.0 – 6.0) |
| Self-employed | 91 (5%) | 94 (5%) | 4.8 (2.0 – 7.0) |
| At work in person | 43 (2%) | 46 (2%) | 6.3 (2.0 – 9.0) |
| Not at work | 24 (1%) | 25 (1%) | 3.7 (1.0 – 5.0) |
| Worked fully online | 24 (1%) | 24 (1%) | 3.2 (1.0 – 4.0) |
| Student & (self-)employed | 92 (5%) | 87 (4%) | 10.7 (3.0 – 15.0) |
| At work/school in person | 58 (3%) | 55 (3%) | 13.7 (4.0 – 21.0) |
| Not at work/school | 26 (1%) | 24 (1%) | 6.0 (3.0 – 8.0) |
| Worked/studied fully online | 8 (0%) | 7 (0%) | 3.3 (2.0 – 4.0) |
| Other | 623 (32%) | 651 (34%) | 3.9 (1.0 – 5.0) |
| **Race/Ethnicity** | | | |
| White, NH | 980 (51%) | 1,117 (58%) | 7.6 (2.0 – 9.0) |
| Hispanic/Latino | 377 (20%) | 370 (19%) | 7.8 (3.0 – 10.0) |
| Black, NH | 271 (14%) | 226 (12%) | 6.3 (2.0 – 7.0) |
| Asian, NH | 163 (8%) | 111 (6%) | 6.3 (2.0 – 8.0) |
| Other, NH | 139 (7%) | 107 (6%) | 7.0 (2.0 – 9.0) |
| **Sex** | | | |
| Female | 909 (47%) | 970 (50%) | 7.7 (2.0 – 10.0) |
| Male | 1,014 (53%) | 953 (49%) | 7.0 (2.0 – 9.0) |
| **Household Income** | | | |
| Less Than $10,000 | 72 (4%) | 71 (4%) | 4.2 (1.0 – 6.0) |
| $10,000 to $24,999 | 136 (7%) | 138 (7%) | 5.4 (1.0 – 7.0) |
| $25,000 to $49,999 | 273 (14%) | 290 (15%) | 6.2 (2.0 – 7.0) |
| $50,000 to $74,999 | 275 (14%) | 297 (15%) | 6.5 (2.0 – 9.0) |
| $75,000 to $99,999 | 248 (13%) | 260 (13%) | 8.4 (3.0 – 10.0) |
| $100,000 to $149,999 | 382 (20%) | 376 (19%) | 8.3 (3.0 – 10.0) |
| $150,000 or more | 544 (28%) | 498 (26%) | 8.3 (2.0 – 10.0) |
| **5-year Age Group** | | | |
| 0-4 | 135 (7%) | 128 (7%) | 6.9 (3.0 – 9.0) |
| 5-9 | 111 (6%) | 112 (6%) | 10.1 (5.0 – 13.0) |
| 10-14 | 125 (6%) | 107 (6%) | 8.7 (4.0 – 11.0) |
| 15-19 | 145 (8%) | 129 (7%) | 10.3 (4.0 – 13.0) |
| 20-24 | 113 (6%) | 113 (6%) | 7.7 (2.0 – 9.0) |
| 25-29 | 117 (6%) | 120 (6%) | 6.0 (2.0 – 8.0) |
| 30-34 | 117 (6%) | 120 (6%) | 8.2 (1.0 – 9.0) |
| 35-39 | 131 (7%) | 135 (7%) | 8.1 (3.0 – 9.0) |
| 40-44 | 144 (7%) | 148 (8%) | 7.7 (2.0 – 10.0) |
| 45-49 | 98 (5%) | 97 (5%) | 7.8 (2.0 – 10.0) |
| 50-54 | 108 (6%) | 113 (6%) | 7.4 (2.0 – 8.0) |
| 55-59 | 128 (7%) | 137 (7%) | 8.9 (2.0 – 11.0) |
| 60-64 | 133 (7%) | 139 (7%) | 5.2 (1.0 – 7.0) |
| 65-69 | 116 (6%) | 114 (6%) | 5.3 (2.0 – 6.0) |
| 70-74 | 99 (5%) | 100 (5%) | 4.5 (1.0 – 7.0) |
| 75-79 | 60 (3%) | 65 (3%) | 5.1 (1.0 – 6.0) |
| 80+ | 50 (3%) | 52 (3%) | 3.8 (1.0 – 5.0) |

| | | | |
|---|---|---|---|
| **Ad-hoc Age Group** | | | |
| 0-4 | 135 (7%) | 128 (7%) | 6.9 (3.0 – 9.0) |
| 5-9 | 111 (6%) | 112 (6%) | 10.1 (5.0 – 13.0) |
| 10-17 | 218 (11%) | 183 (9%) | 9.6 (4.0 – 12.0) |
| 18-29 | 282 (15%) | 286 (15%) | 7.3 (2.0 – 9.0) |
| 30-39 | 248 (13%) | 256 (13%) | 8.2 (2.0 – 9.0) |
| 40-49 | 242 (13%) | 246 (13%) | 7.8 (2.0 – 10.0) |
| 50-59 | 236 (12%) | 250 (13%) | 8.3 (2.0 – 10.0) |
| 60+ | 458 (24%) | 471 (24%) | 4.9 (1.0 – 6.0) |
| **Household Size** | | | |
| 1 | 205 (11%) | 205 (11%) | 4.9 (0.0 – 6.0) |
| 2 | 603 (31%) | 610 (32%) | 6.1 (1.0 – 7.0) |
| 3 | 404 (21%) | 374 (19%) | 7.0 (2.0 – 9.0) |
| 4 | 378 (20%) | 385 (20%) | 9.2 (3.0 – 10.0) |
| 5 | 199 (10%) | 210 (11%) | 8.6 (4.0 – 10.0) |
| 6 | 76 (4%) | 77 (4%) | 9.9 (5.0 – 11.0) |
| 7 and more | 65 (3%) | 69 (4%) | 10.7 (6.0 – 12.0) |
| **Day** | | | |
| Weekday | 1,637 (85%) | 1,632 (85%) | 7.5 (2.0 – 9.0) |
| Weekend | 293 (15%) | 298 (15%) | 6.7 (2.0 – 8.0) |
| **Participant Sick** | | | |
| Yes | 193 (10%) | 194 (10%) | 6.7 (2.0 – 8.0) |
| No | 1,735 (90%) | 1,734 (90%) | 7.4 (2.0 – 9.0) |
| Refused | 2 (0%) | 2 (0%) | 25.7 (1.0 – 41.0) |
| **Household Member Sick** | | | |
| No, Not That I Know Of | 1,697 (88%) | 1,699 (88%) | 7.3 (2.0 – 9.0) |
| Yes | 231 (12%) | 229 (12%) | 8.0 (4.0 – 10.0) |
| Refused | 2 (0%) | 2 (0%) | 0.6 (0.0 – 1.0) |
| **Industry Group** | | | |
| Arts, Information, Finance, and Professional Services | 213 (11%) | 201 (10%) | 5.5 (2.0 – 7.0) |
| Education, Tutoring, and Child Care | 104 (5%) | 105 (5%) | 16.1 (4.0 – 24.0) |
| Factory, Manufacturing, and Woodworking | 71 (4%) | 74 (4%) | 8.3 (3.0 – 11.0) |
| Health Care | 106 (5%) | 108 (6%) | 10.7 (3.0 – 15.0) |
| Infrastructure, Logistics, Construction, and Maintenance | 127 (7%) | 136 (7%) | 6.9 (3.0 – 9.0) |
| Sales and Support Services | 182 (9%) | 191 (10%) | 9.4 (3.0 – 11.0) |
| Other | 112 (6%) | 114 (6%) | 7.1 (2.0 – 9.0) |
| Minor | 464 (24%) | 423 (22%) | 8.9 (4.0 – 11.0) |
| Not Asked or Refused | 551 (29%) | 579 (30%) | 4.0 (1.0 – 5.0) |
| **Occupation Group** | | | |
| Arts, IT, Mathematics, and Physical, Life or Social Sciences | 92 (5%) | 85 (4%) | 5.0 (2.0 – 7.0) |
| Building Cleaning, Maintenance Repair | 48 (2%) | 52 (3%) | 7.4 (3.0 – 9.0) |
| Business Operations | 53 (3%) | 52 (3%) | 6.5 (2.0 – 9.0) |
| Construction, Architecture, and Precision Production | 58 (3%) | 60 (3%) | 6.3 (3.0 – 9.0) |
| Education, Training, and Library | 68 (4%) | 66 (3%) | 19.2 (6.0 – 29.0) |
| Financial Operations and Services | 38 (2%) | 36 (2%) | 6.2 (3.0 – 8.0) |
| Food Preparation and Serving | 38 (2%) | 42 (2%) | 11.2 (3.0 – 12.0) |

| | | | |
|---|---|---|---|
| Health Diagnosing or Treating Practitioners | 34 (2%) | 33 (2%) | 17.4 (5.0 – 33.0) |
| Health Technicians and Care Support | 33 (2%) | 35 (2%) | 9.8 (4.0 – 19.0) |
| Not Asked or Refused | 554 (29%) | 582 (30%) | 4.0 (1.0 – 5.0) |
| Office and Administrative Support, Management and Legal | 170 (9%) | 176 (9%) | 8.0 (1.0 – 11.0) |
| Other | 146 (8%) | 148 (8%) | 7.1 (2.0 – 10.0) |
| Sales and Personal Care Services | 92 (5%) | 97 (5%) | 10.1 (3.0 – 13.0) |
| Transportation, Moving, Community and Social Services | 42 (2%) | 41 (2%) | 6.0 (1.0 – 9.0) |
| **Region** | | | |
| Midwest | 365 (19%) | 400 (21%) | 8.0 (3.0 – 10.0) |
| Northeast | 323 (17%) | 327 (17%) | 6.9 (2.0 – 8.0) |
| South | 751 (39%) | 745 (39%) | 7.4 (2.0 – 9.0) |
| West | 491 (25%) | 458 (24%) | 7.1 (2.0 – 9.0) |
| **Survey Week** | | | |
| May,15 – May,22 | 734 (38%) | 747 (39%) | 7.1 (2.0 – 9.0) |
| May,22 – May,29 | 200 (10%) | 208 (11%) | 6.9 (2.0 – 9.0) |
| May,29 – June,05 | 16 (1%) | 10 (1%) | 3.7 (1.0 – 6.0) |
| October,08 – October,15 | 844 (44%) | 848 (44%) | 7.8 (2.0 – 10.0) |
| October,15 – October,22 | 105 (5%) | 95 (5%) | 7.0 (3.0 – 9.0) |
| October,22 – October,29 | 31 (2%) | 22 (1%) | 7.7 (2.0 – 10.0) |
| **Wave** | | | |
| Wave 1 | 950 (49%) | 965 (50%) | 7.1 (2.0 – 9.0) |
| Wave 2 | 980 (51%) | 965 (50%) | 7.7 (2.0 – 10.0) |

[1] Total number of observations in the sample (% of the sample)

[2] Values represent the weighted number of observations, calculated as the sum of individual participant weights. Percentages in parentheses show the group's share of the total sum of weights, which is equal the total sample size.

[3] Mean number of contacts (50% IQR)

#### 2.2 Association between Respondent Characteristics and Number of Contacts

##### *2.2.1 Univariable Analysis*

When taken in isolation, we observe statistically significant differences in the total number of contacts of self-employed participants with employed, students, and students & (self-) employed, as well as between other activity group and the rest of the main activity groups (Table S 7). Focusing on the mode of attending school and work, we observe that participants attending work and/or school in person had 3.13*** (95% CI: 2.73 - 3.58) times more total contacts than not students nor employed, 3.12*** (95% CI: 2.85 - 3.41) times more contacts than individuals who worked/studied fully online, and 2.50*** (95% CI: 2.22 - 2.81) times more contacts than those who was not at school or work during the reference day. In fact, those who worked/studied fully online reported 20% (RR = 0.8**, 95% CI: 0.68 – 0.94) fewer total contacts than those not at work/school during the reference day. Those not at work/school reported 25% (RR = 1.25***, 95% CI: 1.1 - 1.41) more total contacts than neither students nor employed. All the above-mentioned differences were statistically significant when using a univariate GAM regression (analyzed in isolation). The differences in the number of contacts reported by sick participants and those with sick household members were not statistically significant. The differences in the number of contacts reported by NH White and Hispanic participants relative to NH Black (20% and 23% more contacts) were statistically significant at 5% level, while the differences relative to NH Asian (20% and 22% more contacts) were statistically significant only at 10% level. Other racial/ethnic differences range from 2% to 10%, with no statistical significance, highlighting the need to test the hypothesis using a larger sample. Moreover, these differences need to be reevaluated when other factors, such as household composition and income, school-specific contact patterns and labor market participation, are taken under consideration. In fact, we observe statistically significant differences between the number of contacts reported by participants in difference household income groups, with those having household income of less than $10,000 reporting 51% (RR = 0.51***, 95% CI: 0.4 - 0.66) of the number of contacts of those with household income $150,000 or more. We also observed the number of total contacts increasing with household size, with those living alone reporting 64% (RR = 0.46***, 95% CI: 0.35 - 0.6) fewer contacts than those in households of 7+ people. Negative binomial regression fitted the observed overdispersion in the data the best (theta values ranging between small values of 1.14 and 1.71). Population groups with less than 3 respondents were eliminated from this analysis.

**Table S 7.** Differences in the number of total contacts reported by socio-demographic groups.

| **Characteristic** | **Reference level** | **Compared level** | **RR (95% CI)** |
|---|---|---|---|
| Main Activity | employed | student | 1.00 (0.89 - 1.12) |
| | self-employed | employed | 1.94*** (1.58 - 2.37) |
| | self-employed | student | 1.93*** (1.55 - 2.39) |
| | student & (self-)employed | self-employed | 0.45*** (0.35 - 0.6) |
| | student & (self-)employed | employed | 0.88 (0.72 - 1.07) |
| | student & (self-)employed | student | 0.88 (0.71 - 1.08) |
| | other | student & (self-)employed | 2.74*** (2.24 - 3.38) |
| | other | self-employed | 1.24* (1.01 - 1.54) |
| | other | employed | 2.41*** (2.18 - 2.67) |
| | other | student | 2.4*** (2.13 - 2.71) |
| Attendance Mode | Worked/studied fully online | At work/school in person | 3.13*** (2.73 - 3.58) |
| | Not at work/school | Worked/studied fully online | 0.80** (0.68 - 0.94) |
| | Not at work/school | At work/school in person | 2.50*** (2.22 - 2.81) |
| | Not student nor employed | Not at work/school | 1.25*** (1.1 - 1.41) |
| | Not student nor employed | Worked/studied fully online | 1.00 (0.87 - 1.15) |
| | Not student nor employed | At work/school in person | 3.12*** (2.85 - 3.41) |
| Race/Ethnicity | Hispanic/Latino | White, NH | 0.98 (0.87 - 1.1) |
| | Black, NH | Hispanic/Latino | 1.23* (1.04 - 1.45) |
| | Black, NH | White, NH | 1.20* (1.04 - 1.38) |
| | Asian, NH | Black, NH | 1.00 (0.79 - 1.25) |
| | Asian, NH | Hispanic/Latino | 1.22† (0.98 - 1.51) |
| | Asian, NH | White, NH | 1.20† (0.98 - 1.45) |

| | | | |
|---|---|---|---|
| | Other, NH | Asian, NH | 0.90 (0.69 - 1.18) |
| | Other, NH | Black, NH | 0.90 (0.71 - 1.13) |
| | Other, NH | Hispanic/Latino | 1.10 (0.89 - 1.37) |
| | Other, NH | White, NH | 1.08 (0.88 - 1.31) |
| Sex | Male | Female | 1.09† (1 - 1.19) |
| Respondent Sick | Yes | No | 1.11 (0.96 - 1.29) |
| Household Member Sick | Yes | No, not that I know of | 0.91 (0.79 - 1.04) |
| Household Income | $10,000 to $24,999 | Less than $10,000 | 0.78† (0.58 - 1.05) |
| | $25,000 to $49,999 | $10,000 to $24,999 | 0.88 (0.72 - 1.08) |
| | $25,000 to $49,999 | Less than $10,000 | 0.68** (0.53 - 0.9) |
| | $50,000 to $74,999 | $25,000 to $49,999 | 0.96 (0.81 - 1.12) |
| | $50,000 to $74,999 | $10,000 to $24,999 | 0.84† (0.68 - 1.03) |
| | $50,000 to $74,999 | Less than $10,000 | 0.65** (0.5 - 0.86) |
| | $75,000 to $99,999 | $50,000 to $74,999 | 0.77** (0.66 - 0.91) |
| | $75,000 to $99,999 | $25,000 to $49,999 | 0.74*** (0.63 - 0.87) |
| | $75,000 to $99,999 | $10,000 to $24,999 | 0.65*** (0.53 - 0.8) |
| | $75,000 to $99,999 | Less than $10,000 | 0.51*** (0.39 - 0.66) |
| | $100,000 to $149,999 | $75,000 to $99,999 | 1.01 (0.87 - 1.18) |
| | $100,000 to $149,999 | $50,000 to $74,999 | 0.78** (0.67 - 0.91) |
| | $100,000 to $149,999 | $25,000 to $49,999 | 0.75*** (0.64 - 0.87) |
| | $100,000 to $149,999 | $10,000 to $24,999 | 0.65*** (0.54 - 0.8) |
| | $100,000 to $149,999 | Less than $10,000 | 0.51*** (0.39 - 0.67) |
| | $150,000 or more | $100,000 to $149,999 | 1.00 (0.88 - 1.14) |
| | $150,000 or more | $75,000 to $99,999 | 1.01 (0.87 - 1.17) |
| | $150,000 or more | $50,000 to $74,999 | 0.78*** (0.68 - 0.9) |
| | $150,000 or more | $25,000 to $49,999 | 0.75*** (0.65 - 0.86) |
| | $150,000 or more | $10,000 to $24,999 | 0.65*** (0.54 - 0.79) |
| | $150,000 or more | Less than $10,000 | 0.51*** (0.4 - 0.66) |
| Household Size | 2 | 1 | 0.81** (0.69 - 0.95) |
| | 3 | 2 | 0.88* (0.77 - 1) |
| | 3 | 1 | 0.71*** (0.6 - 0.84) |
| | 4 | 3 | 0.76*** (0.66 - 0.87) |
| | 4 | 2 | 0.66*** (0.59 - 0.75) |
| | 4 | 1 | 0.54*** (0.45 - 0.63) |

| | | | |
|---|---|---|---|
| | 5 | 4 | 1.07 (0.91 - 1.26) |
| | 5 | 3 | 0.81* (0.68 - 0.95) |
| | 5 | 2 | 0.71*** (0.61 - 0.83) |
| | 5 | 1 | 0.57*** (0.47 - 0.69) |
| | 6 | 5 | 0.87 (0.68 - 1.12) |
| | 6 | 4 | 0.93 (0.73 - 1.18) |
| | 6 | 3 | 0.71** (0.55 - 0.89) |
| | 6 | 2 | 0.62*** (0.49 - 0.78) |
| | 6 | 1 | 0.5*** (0.39 - 0.64) |
| | 7+ | 6 | 0.93 (0.68 - 1.26) |
| | 7+ | 5 | 0.81 (0.62 - 1.05) |
| | 7+ | 4 | 0.86 (0.67 - 1.1) |
| | 7+ | 3 | 0.65*** (0.51 - 0.83) |
| | 7+ | 2 | 0.57*** (0.45 - 0.73) |
| | 7+ | 1 | 0.46*** (0.35 - 0.6) |
| Ad-hoc Age Group | 5-9 | 0-4 | 0.69** (0.54 - 0.88) |
| | 10-17 | 5-9 | 1.05 (0.84 - 1.32) |
| | 10-17 | 0-4 | 0.72** (0.58 - 0.9) |
| | 18-29 | 10-17 | 1.31** (1.09 - 1.57) |
| | 18-29 | 5-9 | 1.37** (1.12 - 1.7) |
| | 18-29 | 0-4 | 0.94 (0.77 - 1.16) |
| | 30-39 | 18-29 | 0.9 (0.76 - 1.06) |
| | 30-39 | 10-17 | 1.17† (0.98 - 1.41) |
| | 30-39 | 5-9 | 1.23† (1 - 1.53) |
| | 30-39 | 0-4 | 0.85 (0.69 - 1.04) |
| | 40-49 | 30-39 | 1.06 (0.89 - 1.25) |
| | 40-49 | 18-29 | 0.95 (0.8 - 1.12) |
| | 40-49 | 10-17 | 1.24* (1.03 - 1.49) |
| | 40-49 | 5-9 | 1.30* (1.05 - 1.62) |
| | 40-49 | 0-4 | 0.89 (0.72 - 1.1) |
| | 50-59 | 40-49 | 0.94 (0.79 - 1.11) |
| | 50-59 | 30-39 | 0.99 (0.84 - 1.17) |
| | 50-59 | 18-29 | 0.89 (0.75 - 1.05) |
| | 50-59 | 10-17 | 1.16 (0.97 - 1.4) |
| | 50-59 | 5-9 | 1.22† (0.99 - 1.52) |

| | | | |
|---|---|---|---|
| | 50-59 | 0-4 | 0.84† (0.68 - 1.03) |
| | 60+ | 50-59 | 1.69*** (1.45 - 1.96) |
| | 60+ | 40-49 | 1.58*** (1.36 - 1.84) |
| | 60+ | 30-39 | 1.67*** (1.44 - 1.94) |
| | 60+ | 18-29 | 1.5*** (1.3 - 1.73) |
| | 60+ | 10-17 | 1.96*** (1.66 - 2.32) |
| | 60+ | 5-9 | 2.06*** (1.69 - 2.52) |
| | 60+ | 0-4 | 1.41*** (1.17 - 1.72) |
| Day | Weekend | Weekday | 1.13† (0.99 - 1.28) |
| Wave | Wave 2 | Wave 1 | 0.92† (0.84 – 1.00) |

RR is calculated as exponentiated coefficient of the univariate GAM regression with the reference group. †p < 0.1; *p < 0.05; **p < 0.01; ***p < 0.001

##### *2.2.2 Interconnected Respondent Characteristics*

The analysis of contact data presents a unique challenge due to the inherent complexity of connections. It is important to recognize that the various socio-demographic characteristics of the respondents inherently reflect socio-demographic processes and, therefore, are often interconnected (correlated). Table S 6 details differences in the number of contacts by respondent characteristics analyzed in isolation, providing a univariate view of the relationships between observed respondent characteristics and the number of reported contacts. However, a deeper understanding requires acknowledging the complex interconnected reality. In fact, Table S 7 and Table S 8 illustrate observed statistically significant differences between socio-demographic characteristics defining population groups. For example, Table S 8 showing age differences by population group wile accounting for survey design, illustrates that students are typically younger, while those in the "Other" main activity category (which includes retirees) are generally older. Similarly, larger households often imply the presence of children, which directly relates not only to the lower age of the participating children but also to the age of the parents within those households. The data in Table S 6 supports this: the higher contact numbers for "Students" (e.g., 9.4 total contacts, 6.3 non-household) align with the higher contacts seen in younger age groups (e.g., 10-14, 15-19). Conversely, the lower contact numbers for the other main activity group (3.9 total, 2.2 non-household) correspond with the smaller number of contacts observed in older age groups (e.g., 60+). Respondent age is consistently and statistically correlated with practically every other characteristic in Table S 7.

**Table S 8.** Difference in mean age between population groups using two-sample t-test.

| Characteristic | Reference level | Compared level | Age Difference (95% CI) |
|---|---|---|---|
| Main Activity | student | employed | 33.13*** (95% CI:32 – 34) |
| | student | self-employed | 37.05*** (95% CI:34 – 40) |
| | student | student & (self-)employed | 14.81*** (95% CI:12 – 17) |
| | student | other | 39.92*** (95% CI:38 – 42) |

| | | | |
|---|---|---|---|
| | employed | self-employed | 3.92** (95% CI:1 – 7) |
| | employed | student & (self-)employed | -18.32*** (95% CI: -21 – -16) |
| | employed | other | 6.79*** (95% CI:4 – 9) |
| | self-employed | student & (self-)employed | -22.24*** (95% CI: -26 – -18) |
| | student & (self-) employed | other | 25.11*** (95% CI:22 – 28) |
| Attendance Mode | At work/school in person | Worked/studied fully online | 10.78*** (95% CI:8 – 13) |
| | At work/school in person | Not student or employed | 18.44*** (95% CI:16 – 21) |
| | Worked/studied fully online | Not at work/school | -11.38*** (95% CI:-14 – -8) |
| | Not at work/school | Not student or employed | 19.04*** (95% CI:16 – 22) |
| Race/Ethnicity | White, NH | Hispanic/Latino | -9.72*** (95% CI:-12 – -7) |
| | White, NH | Black, NH | -6.27*** (95% CI:-9 – -3) |
| | White, NH | Other, NH | -11.59*** (95% CI:-16 – -8) |
| | Hispanic/Latino | Black, NH | 3.45* (95% CI:0 – 7) |
| | Hispanic/Latino | Asian, NH | 7.08*** (95% CI:3 – 11) |
| | Black, NH | Other, NH | -5.32** (95% CI:-10 – -1) |
| | Asian, NH | Other, NH | -8.95*** (95% CI:-14 – -4) |
| Sex | Female | Male | -1.91*(95% CI: -4– 0) |
| Participant Sick | No | Yes | -8.61*** (95% CI:-12 – -5) |
| Household Member Sick | No, not that I know of | Yes | -9.94*** (95% CI:-13 – -7) |
| Household Income | $150,000 or more | $25,000 to $49,999 | -4.59** (95% CI:-8 – -1) |
| | $150,000 or more | $50,000 to $74,999 | -4.2** (95% CI:-8 – -1) |
| | Less than $10,000 | $10,000 to $24,999 | 6.3* (95% CI:0 – 13) |
| | Less than $10,000 | $25,000 to $49,999 | 7.49** (95% CI:2 – 13) |
| | Less than $10,000 | $50,000 to $74,999 | 7.1** (95% CI:1 – 13) |
| | $10,000 to $24,999 | $75,000 to $99,999 | -5.68** (95% CI:-11 – -1) |
| | $25,000 to $49,999 | $75,000 to $99,999 | -6.87*** (95% CI:-11 – -3) |
| | $25,000 to $49,999 | $100,000 to $149,999 | -3.64* (95% CI:-7 – 0) |
| | $50,000 to $74,999 | $75,000 to $99,999 | -6.48*** (95% CI:-11 – -2) |
| | $50,000 to $74,999 | $100,000 to $149,999 | -3.25* (95% CI:-7 – 0) |
| Day | Weekday | Weekend | -4.65*** (95% CI:-8 – -2) |

*p < 0.05; **p < 0.01; ***p < 0.001

Additionally, Table S9 reports statistically significant correlations between the categorical respondents' characteristics. The magnitude of the connection, illustrated by the value of the Cramer's V, is mostly low, highlighting that no single characteristic can fully proxy another. However, the presence and consistent statistical significance of these correlations indicate that to better understand the observed differences in the number of contacts, one needs to control for the joint impacts of multiple socio-demographic characteristics. This is why in addition to the univariate analysis described in this section we also perform the multivariable analysis (Table S 10 and Table S 11, Figure S 1 and Figure S 2).

**Table S9.** Correlations between respondent socio-demographic characteristics.

| **Characteristics** | | **Cramer's V** | **Correlation Magnitude** |
|---|---|---|---|
| Main Activity | Race/Ethnicity | 0.022*** | Very Weak |
| | Sex | 0.062*** | Very Weak |
| | Household Member Sick | 0.028*** | Very Weak |
| | Household Size | 0.041*** | Very Weak |
| | Household Income | 0.028*** | Very Weak |
| | Weekend/Weekday | 0.049*** | Very Weak |
| Activity Mode | Race/Ethnicity | 0.026*** | Very Weak |
| | Sex | 0.074*** | Very Weak |
| | Household Size | 0.035*** | Very Weak |
| | Household Income | 0.039*** | Very Weak |
| | Weekend/Weekday | 0.176*** | Weak |
| | Wave | 0.047*** | Very Weak |
| Race/Ethnicity | Sex | 0.032* | Very Weak |
| | Participant Sick | 0.023** | Very Weak |
| | Household Member Sick | 0.024** | Very Weak |
| | Household Size | 0.027*** | Very Weak |
| | Household Income | 0.029*** | Very Weak |
| | Weekend/Weekday | 0.036** | Very Weak |
| Sex | Household Income | 0.049*** | Very Weak |
| Participant Sick | Household Member Sick | 0.077*** | Very Weak |
| | Household Size | 0.031*** | Very Weak |
| Household Member Sick | Household Size | 0.038*** | Very Weak |
| | Household Income | 0.025*** | Very Weak |
| Household Size | Household Income | 0.016*** | Very Weak |
| | Weekend/Weekday | 0.034** | Very Weak |
| Day: Weekend/Weekday | Wave | 0.21*** | Moderate |

Stars reflect the statistical significance of the Pearson's chi-squared test (Rao & Scott adjustment).

##### 2.2.3 *Multivariable Regression Analysis*

After adjusting for all covariates in a multivariable Generalized Additive Model (GAM), several factors emerged as significant predictors of the total number of contacts (Table S 10). The model providing the best fit, as determined by the Akaike Information Criterion (AIC), successfully accounts for approximately 26.2% of the total variance ($R^2$=0.262) in the number of total contacts reported by participants (Table S 10, Column [1]). This is the model presented in the main text of the paper. As sensitivity analysis, we also show results of the best fit models based on $R^2$ (Table S 10, Column [2]), which provides a marginal $R^2$ improvement (0.256) and uses ad-hoc age groups rather than numeric age, and the percentage of deviance explained (Table S 10, Column [3]), which includes 5-year age groups instead of numeric age. We also illustrate that removing the mode of participation (e.g., in-person vs. remote) from the analysis and accounting for the main activity only (e.g., student, employed) notably decreases the goodness-of-fit (e.g., $R^2$=0.140).

The combined variable of main activity and attendance mode proved to be a primary driver of contact patterns. Using students attending school in person as the reference, we found that employed individuals physically present at their workplace reported 44% more contacts (RR=1.44, 95% CI: 1.20 - 1.73, $p<0.001$). Conversely, all groups not physically present at a school or workplace reported significantly fewer contacts. For instance, individuals who worked or studied fully online had approximately 50-65% fewer contacts than students attending school in person (e.g., Employed working online: RR=0.44, 95% CI: 0.35 - 0.54, $p<0.001$). Similarly, those in the "Other" activity group, who are not students nor employed, had 60% fewer contacts (RR=0.40, 95% CI: 0.34 - 0.47, $p<0.001$) than the reference group.

An important distinction with respect to the trends in group means (Table S 6) is observed for the connection between age and the total number of contacts. Age-group differences are not statistically significant for most groups, except for ages 20-29, and 60-64. The negative correlation between the mean number of contacts and being 60+ (Table S 10) is not statistically significant after adjustment for main activity and mode of attendance. In fact, we observe the statistically significant non-linear effects of age as a continuous variable (Figure S 1). Based on model estimates for the marginal effects of age, the predicted number of contacts for the reference group (NH White, male, healthy in-person students from 3-person households with income of $150,000 and more) would be steadily decreasing in childhood until reaching its lowest point around age 25, almost plateauing after that with a small peak around the age of 40 and reaching the second lowest point at around the age of 60. After 60, the contacts would begin to rise in older age, reaching its highest peak around age 75. This highlights that the lower number of contacts observed for older adults is mostly associated with the decreased participation in the labor market and education during life transition periods.

Key sociodemographic characteristics also remained significant predictors after adjustment. Females reported 21% more contacts than males (RR=1.21, 95% CI: 1.12 - 1.31, $p<0.001$). A strong, positive dose-response relationship was observed with household size; compared to 3-person households, individuals in 1-person households reported 27% fewer contacts (RR=0.73, 95% CI: 0.62 - 0.85, $p<0.001$), an effect that grew to 65% more contacts (RR=1.65, 95% CI: 1.34 - 2.04, $p<0.001$) for those in households of seven or more. A clear gradient was also evident for household income. Using the highest income category (>$150,000) as the reference, all lower-income groups reported significantly fewer contacts. This effect was most pronounced for the lowest income group (<$10,000), which reported 35% fewer contacts (RR=0.65, 95% CI: 0.52 - 0.82, $p<0.001$). After controlling for these factors, statistically significant differences in race and ethnicity persisted. Compared to Non-Hispanic (NH) White participants, NH Black participants reported 13% fewer contacts (RR=0.87, 95% CI: 0.76 - 0.98, p=0.025) and NH Asian participants reported 25% fewer contacts (RR=0.75, 95% CI: 0.63 - 0.89, p<0.001).

Finally, participants who reported being sick showed a tendency towards fewer contacts, an effect that reached statistical significance in some models but disappeared when accounting for both, main activity and the mode of participation.

It is important to note that the results of multivariable analysis, which accounts for the simultaneous influence of multiple factors, may differ from the findings from the univariable analysis or crude group comparisons. For example, the link with race and ethnicity was slightly refined. In the univariable analysis, NH White participants had 20% more contacts than NH Black participants. After adjusting for confounding variables, this relationship held but was attenuated, with NH Whites having approximately 15% more contacts (RR = 1/0.87, for comparable reference groups). In contrast, the difference between NH White and NH Asian participants became more pronounced from 20% more contacts in the univariable model to a 33% more contacts in the adjusted model and became highly statistically significant. The effect of day of the week and wave differences disappeared once accounted for main activity and mode of attendance.

**Table S 10.** Results of multivariable GAM analysis for total contacts.

| | **Best AIC [1]** | | **Best $R^2$ [2]** | | **Best % Deviance [3]** | | **Main Activity only [4]** | |
|---|---|---|---|---|---|---|---|---|
| **Characteristic** | **RR (95% CI)[1]** | **p-value** | **RR (95% CI)[1]** | **p-value** | **RR (95% CI)[1]** | **p-value** | **RR (95% CI)[1]** | **p-value** |
| **Main Activity and Attendance Mode** | | | | | | | | |
| Student: At school in person | ref. | | ref. | | ref. | | | |
| Employed: At work in person | 1.44*** (1.20, 1.73) | **<0.001** | 1.46*** (1.20, 1.76) | **<0.001** | 1.54*** (1.27, 1.88) | **<0.001** | | |
| Self-Employed: At work in person | 0.76 (0.56, 1.02) | 0.064 | 0.75 (0.56, 1.02) | 0.064 | 0.84 (0.62, 1.14) | 0.3 | | |
| Student & (Self-) Employed: At work/school in person | 1.40** (1.10, 1.79) | **0.007** | 1.49** (1.16, 1.92) | **0.002** | 1.42** (1.10, 1.84) | **0.007** | | |
| Student: Not at school | 0.43*** (0.35, 0.52) | **<0.001** | 0.43*** (0.35, 0.52) | **<0.001** | 0.42*** (0.34, 0.52) | **<0.001** | | |
| Employed: Not at work | 0.52*** (0.42, 0.65) | **<0.001** | 0.53*** (0.42, 0.66) | **<0.001** | 0.55*** (0.43, 0.70) | **<0.001** | | |
| Self-Employed: Not at work | 0.42*** (0.28, 0.62) | **<0.001** | 0.43*** (0.29, 0.64) | **<0.001** | 0.43*** (0.29, 0.64) | **<0.001** | | |
| Student & (Self-) Employed: Not at work/school | 0.60** (0.42, 0.86) | **0.006** | 0.64* (0.44, 0.93) | **0.018** | 0.61* (0.42, 0.89) | **0.010** | | |
| Other: Not student or employed | 0.40*** (0.34, 0.47) | **<0.001** | 0.42*** (0.35, 0.50) | **<0.001** | 0.43*** (0.36, 0.51) | **<0.001** | | |
| Student: Studied fully online | 0.52* (0.31, 0.87) | **0.012** | 0.55* (0.33, 0.93) | **0.026** | 0.56* (0.33, 0.94) | **0.028** | | |
| Employed: Worked fully online | 0.44*** (0.35, 0.54) | **<0.001** | 0.44*** (0.35, 0.55) | **<0.001** | 0.46*** (0.37, 0.58) | **<0.001** | | |

| | | | | | | | | |
|---|---|---|---|---|---|---|---|---|
| Self-Employed: Worked fully online | 0.36*** (0.24, 0.55) | **<0.001** | 0.37*** (0.24, 0.56) | **<0.001** | 0.39*** (0.26, 0.60) | **<0.001** | | |
| Student & (Self-)Employed: Worked/studied fully online | 0.42* (0.21, 0.83) | **0.012** | 0.41* (0.21, 0.82) | **0.011** | 0.45* (0.22, 0.90) | **0.024** | | |
| **Main Activity** | | | | | | | | |
| student | | | | | | | ref. | |
| employed | | | | | | | 1.41*** (1.15, 1.71) | **<0.001** |
| self-employed | | | | | | | 0.77 (0.59, 1.01) | 0.055 |
| student & (self-)employed | | | | | | | 1.42** (1.11, 1.80) | **0.005** |
| other | | | | | | | 0.52*** (0.43, 0.63) | **<0.001** |
| **Race/Ethnicity** | | | | | | | | |
| White, NH | ref. | | ref. | | ref. | | ref. | |
| Hispanic/Latino | 0.92 (0.83, 1.01) | 0.092 | 0.91 (0.82, 1.01) | 0.063 | 0.94 (0.84, 1.04) | 0.2 | 0.97 (0.87, 1.09) | 0.7 |
| Black, NH | 0.87* (0.76, 0.98) | **0.025** | 0.86* (0.76, 0.98) | **0.019** | 0.88 (0.78, 1.01) | 0.064 | 0.86* (0.75, 0.99) | **0.034** |
| Asian, NH | 0.75*** (0.63, 0.89) | **<0.001** | 0.75*** (0.63, 0.89) | **<0.001** | 0.80* (0.67, 0.95) | **0.012** | 0.77** (0.64, 0.93) | **0.008** |
| Other, NH | 0.92 (0.77, 1.09) | 0.3 | 0.91 (0.76, 1.08) | 0.3 | 0.93 (0.78, 1.11) | 0.4 | 0.88 (0.73, 1.07) | 0.2 |
| **Sex** | | | | | | | | |
| Male | ref. | | ref. | | ref. | | ref. | |
| Female | 1.21*** (1.12, 1.31) | **<0.001** | 1.21*** (1.12, 1.31) | **<0.001** | 1.23*** (1.14, 1.33) | **<0.001** | 1.19*** (1.10, 1.30) | **<0.001** |
| **Sick** | | | | | | | | |
| No | ref. | | | | ref. | | ref. | |
| Yes | 0.89 (0.78, 1.01) | 0.067 | | | 0.89 (0.78, 1.02) | 0.090 | 0.85* (0.74, 0.98) | **0.026** |
| **Household Size** | | | | | | | | |
| 1 | 0.73*** (0.62, 0.85) | **<0.001** | 0.74*** (0.63, 0.87) | **<0.001** | 0.72*** (0.61, 0.84) | **<0.001** | 0.73*** (0.62, 0.87) | **<0.001** |
| 2 | 0.94 (0.84, 1.06) | 0.3 | 0.95 (0.84, 1.06) | 0.3 | 0.93 (0.83, 1.05) | 0.2 | 0.93 (0.82, 1.06) | 0.3 |
| 3 | ref. | | ref. | | ref. | | ref. | |
| 4 | 1.18** (1.05, 1.33) | **0.007** | 1.16* (1.03, 1.31) | **0.014** | 1.15* (1.02, 1.30) | **0.023** | 1.19* (1.04, 1.35) | **0.012** |
| 5 | 1.13 (0.98, 1.30) | 0.090 | 1.12 (0.97, 1.29) | 0.12 | 1.10 (0.96, 1.27) | 0.2 | 1.14 (0.97, 1.33) | 0.10 |
| 6 | 1.41*** (1.15, 1.73) | **<0.001** | 1.37** (1.12, 1.68) | **0.003** | 1.36** (1.11, 1.67) | **0.004** | 1.40*** (1.12, 1.76) | **0.003** |
| 7+ | 1.65*** (1.34, 2.04) | **<0.001** | 1.66*** (1.34, 2.04) | **<0.001** | 1.66*** (1.35, 2.05) | **<0.001** | 1.76*** (1.40, 2.21) | **<0.001** |
| **Household Income** | | | | | | | | |
| Less than $10,000 | 0.65*** (0.52, 0.82) | **<0.001** | 0.64*** (0.51, 0.81) | **<0.001** | 0.64*** (0.51, 0.81) | **<0.001** | 0.70** (0.55, 0.90) | **0.005** |
| $10,000 to $24,999 | 0.77** (0.65, 0.92) | **0.003** | 0.76*** (0.64, 0.90) | **0.002** | 0.76** (0.64, 0.91) | **0.003** | 0.86 (0.71, 1.04) | 0.11 |

| | | | | | | | | |
|---|---|---|---|---|---|---|---|---|
| $25,000 to $49,999 | 0.78*** (0.69, 0.89) | **<0.001** | 0.78*** (0.69, 0.89) | **<0.001** | 0.77*** (0.67, 0.87) | **<0.001** | 0.87* (0.76, 1.00) | **0.050** |
| $50,000 to $74,999 | 0.84** (0.74, 0.95) | **0.006** | 0.85* (0.75, 0.96) | **0.010** | 0.83** (0.73, 0.94) | **0.004** | 0.92 (0.80, 1.06) | 0.2 |
| $75,000 to $99,999 | 0.90 (0.79, 1.02) | 0.11 | 0.90 (0.80, 1.03) | 0.12 | 0.89 (0.79, 1.02) | 0.089 | 0.97 (0.84, 1.12) | 0.7 |
| $100,000 to $149,999 | 0.99 (0.88, 1.10) | 0.8 | 1.0 (0.89, 1.11) | >0.9 | 0.99 (0.88, 1.11) | 0.8 | 1.00 (0.89, 1.13) | >0.9 |
| $150,000 or more | ref. | | ref. | | ref. | | ref. | |
| **Age (spline)** | | **0.030** | | | | | | |
| **Ad-hoc Age Group** | | | | | | | | |
| 0-4 | | | 1.03 (0.85, 1.25) | 0.8 | | | | |
| 5-9 | | | 1.17 (0.97, 1.42) | 0.11 | | | | |
| 10-17 | | | ref. | | | | | |
| 18-29 | | | 0.79* (0.66, 0.96) | **0.017** | | | | |
| 30-39 | | | 0.90 (0.73, 1.11) | 0.3 | | | | |
| 40-49 | | | 0.91 (0.73, 1.12) | 0.4 | | | | |
| 50-59 | | | 0.90 (0.73, 1.12) | 0.4 | | | | |
| 60+ | | | 0.95 (0.77, 1.16) | 0.6 | | | | |
| **5-year Age Group** | | | | | | | | |
| 0-4 | | | | | 0.99 (0.80, 1.22) | >0.9 | 1.01 (0.81, 1.28) | >0.9 |
| 5-9 | | | | | 1.14 (0.92, 1.40) | 0.2 | 1.15 (0.91, 1.45) | 0.2 |
| 10-14 | | | | | 0.89 (0.72, 1.11) | 0.3 | 0.95 (0.75, 1.20) | 0.7 |
| 15-19 | | | | | ref. | | ref. | |
| 20-24 | | | | | 0.76 * (0.61, 0.96) | **0.022** | 0.73* (0.57, 0.94) | **0.015** |
| 25-29 | | | | | 0.68** (0.53, 0.86) | **0.002** | 0.65** (0.50, 0.85) | **0.001** |
| 30-34 | | | | | 0.83 (0.65, 1.05) | 0.12 | 0.86 (0.66, 1.11) | 0.2 |
| 35-39 | | | | | 0.84 (0.67, 1.07) | 0.2 | 0.82 (0.63, 1.06) | 0.12 |
| 40-44 | | | | | 0.87 (0.69, 1.09) | 0.2 | 0.81 (0.63, 1.04) | 0.10 |
| 45-49 | | | | | 0.79 (0.61, 1.02) | 0.067 | 0.79 (0.60, 1.04) | 0.087 |
| 50-54 | | | | | 0.82 (0.64, 1.05) | 0.11 | 0.78 (0.60, 1.01) | 0.062 |
| 55-59 | | | | | 0.85 (0.67, 1.09) | 0.2 | 0.87 (0.67, 1.13) | 0.3 |
| 60-64 | | | | | 0.70** (0.55, 0.89) | **0.004** | 0.70** (0.54, 0.91) | **0.007** |
| 65-69 | | | | | 0.98 (0.76, 1.26) | 0.9 | 0.95 (0.72, 1.25) | 0.7 |

| | | | | | | | | |
|---|---|---|---|---|---|---|---|---|
| 70-74 | | | | | 0.97 (0.74, 1.27) | 0.8 | 0.98 (0.74, 1.31) | >0.9 |
| 75-79 | | | | | 1.10 (0.82, 1.47) | 0.5 | 1.08 (0.78, 1.48) | 0.6 |
| 80+ | | | | | 0.93 (0.68, 1.29) | 0.7 | 0.90 (0.63, 1.27) | 0.5 |
| **Region** | | | | | | | | |
| Midwest | | | | | ref. | | ref. | |
| Northeast | | | | | 0.88* (0.77, 0.99) | **0.036** | 0.87 (0.76, 1.00) | 0.052 |
| South | | | | | 0.90* (0.81, 1.00) | **0.044** | 0.90 (0.80, 1.01) | 0.073 |
| West | | | | | 0.90 (0.80, 1.01) | 0.085 | 0.89 (0.78, 1.01) | 0.066 |
| **Day** | | | | | | | | |
| Weekday | | | | | ref. | | ref. | |
| Weekend | | | | | 1.09 (0.97, 1.22) | 0.15 | 0.85** (0.76, 0.96) | **0.008** |
| **Additional control variables** (not statistically significant) | | | | | **Household Member Sick, Survey Week** | | **Household Member Sick, Survey Week** | |
| **AIC** | **10,965** | | **10,972** | | **10,980** | | **11,330** | |
| **$R^2$** | **0.262** | | **0.262** | | **0.256** | | **0.140** | |

The goodness-of-fit measure or the variable distinguishing selected specification is indicated in the column name. [1] Relative Ratios (RR) are the exponentiated coefficients from the multivariable Generalized Additive Model (GAM). *p<0.05; **p<0.01; ***p<0.001. Abbreviation: CI = Confidence Interval

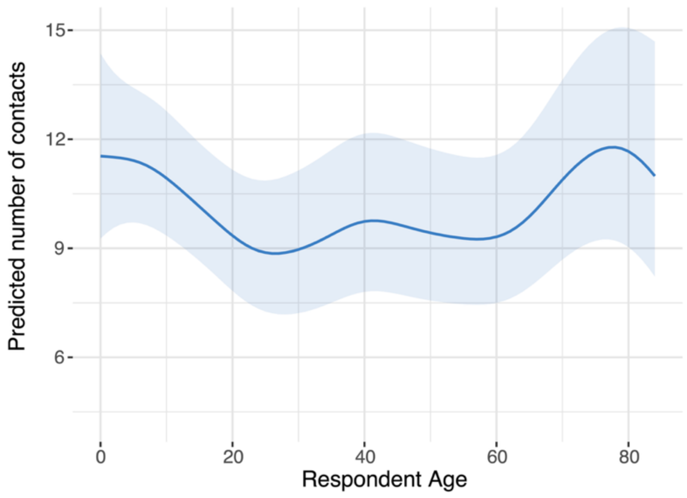


**Figure S 1.** Predicted number of contacts by age (marginal effects) for the reference group.

#### 2.3 Descriptive Analysis of the Number of Non-Household Contacts

We observed very similar trends in mean non-household contacts relative to the total contacts, with a small and not statistically significant difference in the ranking of race/ethnicity groups. A descriptive analysis of daily non-household contact patterns is presented in Table S 11, focusing on interactions outside the household. Across the entire population, participants reported a mean of 5.4 non-household contacts (IQR: 0.0 - 7.0). These interactions were a combination of three primary social settings: work (mean=2.3), school (mean=1.2), and other (mean=1.9). It is important to note that these figures represent population-level averages, meaning they are diluted by individuals who have zero contacts in a given setting (e.g., non-students have zero school contacts).

As for the total contacts, the combination of a person's main activity and their mode of attendance was the most significant factor influencing non-household contacts. Individuals physically present at work or school reported the highest number of non-household contacts. Employed individuals at their workplace averaged 11.2 non-household contacts, while students who were also employed and attended in person averaged 11.6. For these groups, the vast majority of their external contacts occurred within their primary social setting; for instance, work-related interactions (mean = 8.8) dominated the non-household contacts of in-person employees. Conversely, individuals participating remotely had dramatically fewer non-household contacts. Those studying fully online reported the lowest average of just 1.0 non-household contact, followed by those working fully online (1.5 contacts) with very small difference relative to fully online self-employed and students & (self-)employed. For these remote groups, contacts in the school and work settings were virtually zero, meaning their limited non-household interactions occurred exclusively in "Other" social settings.

Non-household contacts followed a distinct pattern across the lifespan, closely tied to participation in specific social settings. The number of non-household contacts peaked during school-age years, reaching a mean of 6.8 for children aged 5-9 and 7.5 for those aged 15-19. This peak was driven almost entirely by interactions within the school setting. Contacts dipped in young adulthood (mean = 4.3 for ages 25-29) before rising to a second peak before average age of retirement (mean = 7.5 for ages 55-59). This later peak was predominantly fueled by interactions in the work setting. After age 60, as participation in the work setting declined, the average number of non-household contacts fell steadily, reaching a low of 2.8 for those 80 and older. For these oldest age groups, school and work contacts were non-existent, and their external interactions took place entirely in "Other" settings.

The number of non-household contacts also varied by sociodemographic characteristics. A clear gradient of non-household contacts increasing with income is visible, driven by the contribution of all three social settings. The lowest-income group (<$10,000) reported a mean of 2.7 non-household contacts, which rose to 6.2 and over for all income brackets above $75,000. NH White participants reported the highest mean number of non-household contacts (5.8), while NH Asian participants reported the lowest (4.3). The highest contribution of "Other" setting to non-household contacts is observed among NH White and NH Asian. At the same time, Hispanics/Latino and NH Black have almost identical distribution of the setting-specific contributions with the highest impact of work contacts. Unlike total contacts, the relationship between household size and non-household contacts was not linear. Individuals in 1-person households reported a relatively high number of non-household contacts (4.6), which peaked for 4-person households (6.5) before declining slightly for the largest households. This trend is the result of very different patterns in individual settings. For example, the highest number of contacts in "Other" setting is reported by 2-person households. In contrast, school contacts are highest for individuals in households with 4-6 people, while work contacts are highest for 1-person and 4-person households. Females reported slightly more non-household contacts on average than males (5.7 vs. 5.2), mainly resulting from the differences in the "other" setting.

Population-level averages of the number of contacts are also 70% higher for work contacts during a weekday, relative to weekends, while relatively similar for other settings. Respondents who were sick also reported fewer non-household contacts in all settings, especially work and "other". Even higher decrease in work contacts is observed for those with a sick household member, while the average number of school contacts for those respondents increases.

**Table S 11.** Additional descriptive statistics.

| **Characteristic** | **Number of total contacts**[1] | **Number of non-household contacts**[1] | **Household**[2] | **School**[2] | **Work**[2] | **Other**[2] |
|---|---|---|---|---|---|---|
| **Total Population** | 7.4 (2.0 - 9.0) | 5.4 (0.0 - 7.0) | 1.9 (26.0%) | 1.2 (16.0%) | 2.3 (32.0%) | 1.9 (25.0%) |
| **Main Activity & Attendance Mode** | | | | | | |
| Student | 9.4 (4.0 - 12.0) | 6.3 (1.0 - 8.0) | 3.1 (33.0%) | 4.8 (51.0%) | 0.0 (0.0%) | 1.5 (16.0%) |
| At school in person | 11.4 (6.0 - 14.0) | 8.3 (4.0 - 10.0) | 3.0 (27.0%) | 6.9 (61.0%) | 0.0 (0.0%) | 1.4 (12.0%) |
| Not at school | 5.0 (3.0 - 6.0) | 1.9 (0.0 - 3.0) | 3.1 (61.0%) | 0.0 (1.0%) | 0.0 (0.0%) | 1.9 (38.0%) |
| Studied fully online | 4.7 (4.0 - 5.0) | 1.0 (0.0 - 1.0) | 3.7 (79.0%) | 0.0 (0.0%) | 0.0 (0.0%) | 1.0 (21.0%) |
| Employed | 9.4 (3.0 - 11.0) | 7.7 (1.0 - 10.0) | 1.7 (18.0%) | 0.6 (7.0%) | 5.3 (57.0%) | 1.8 (19.0%) |
| At work in person | 13.0 (5.0 - 18.0) | 11.2 (3.0 - 15.0) | 1.8 (14.0%) | 1.1 (8.0%) | 8.8 (68.0%) | 1.4 (10.0%) |
| Not at work | 4.8 (2.0 - 7.0) | 3.2 (0.0 - 5.0) | 1.6 (33.0%) | 0.0 (0.0%) | 0.4 (9.0%) | 2.7 (58.0%) |
| Worked fully online | 4.0 (1.0 - 6.0) | 2.5 (0.0 - 4.0) | 1.5 (37.0%) | 0.0 (0.0%) | 0.4 (11.0%) | 2.1 (52.0%) |
| Self-employed | 4.8 (2.0 - 7.0) | 3.2 (0.0 - 4.0) | 1.6 (34.0%) | 0.1 (2.0%) | 1.5 (30.0%) | 1.7 (34.0%) |
| At work in person | 6.3 (2.0 - 9.0) | 4.6 (0.0 - 7.0) | 1.7 (27.0%) | 0.2 (3.0%) | 2.9 (46.0%) | 1.5 (24.0%) |
| Not at work | 3.7 (1.0 - 5.0) | 2.0 (0.0 - 3.0) | 1.8 (47.0%) | 0.0 (0.0%) | 0.0 (0.0%) | 2.0 (53.0%) |
| Worked fully online | 3.2 (1.0 - 4.0) | 1.7 (0.0 - 2.0) | 1.5 (46.0%) | 0.0 (0.0%) | 0.2 (6.0%) | 1.5 (48.0%) |
| Student & (self-) employed | 10.7 (3.0 - 15.0) | 8.5 (1.0 - 15.0) | 2.2 (20.0%) | 2.3 (21.0%) | 4.6 (43.0%) | 1.7 (16.0%) |
| At work/school in person | 13.7 (4.0 - 21.0) | 11.6 (3.0 - 17.0) | 2.1 (16.0%) | 3.6 (26.0%) | 6.7 (49.0%) | 1.3 (9.0%) |
| Not at work/school | 6.0 (3.0 - 8.0) | 3.7 (0.0 - 5.0) | 2.3 (38.0%) | 0.0 (0.0%) | 1.1 (19.0%) | 2.6 (43.0%) |
| Worked/studied fully online | 3.3 (2.0 - 4.0) | 1.6 (1.0 - 3.0) | 1.7 (51.0%) | 0.0 (0.0%) | 0.0 (0.0%) | 1.6 (49.0%) |
| Other | 3.9 (1.0 - 5.0) | 2.2 (0.0 - 3.0) | 1.7 (43.0%) | 0.0 (0.0%) | 0.0 (0.0%) | 2.2 (57.0%) |
| **Race/Ethnicity** | | | | | | |
| White, NH | 7.6 (2.0 - 9.0) | 5.8 (0.0 - 7.0) | 1.8 (24.0%) | 1.2 (15.0%) | 2.5 (33.0%) | 2.1 (28.0%) |
| Hispanic/Latino | 7.8 (3.0 - 10.0) | 5.4 (0.0 - 7.0) | 2.3 (30.0%) | 1.3 (17.0%) | 2.5 (32.0%) | 1.6 (21.0%) |
| Black, NH | 6.3 (2.0 - 7.0) | 4.6 (0.0 - 5.0) | 1.7 (28.0%) | 1.1 (17.0%) | 2.1 (34.0%) | 1.4 (22.0%) |
| Asian, NH | 6.3 (2.0 - 8.0) | 4.3 (0.0 - 5.0) | 2.0 (32.0%) | 1.0 (15.0%) | 1.7 (27.0%) | 1.6 (25.0%) |
| Other, NH | 7.0 (2.0 - 9.0) | 4.9 (0.0 - 6.0) | 2.1 (30.0%) | 1.9 (27.0%) | 1.5 (21.0%) | 1.5 (22.0%) |

| | | | | | | |
|---|---|---|---|---|---|---|
| **Sex** | | | | | | |
| Female | 7.7 (2.0 - 10.0) | 5.7 (0.0 - 7.0) | 2.0 (26.0%) | 1.2 (16.0%) | 2.4 (31.0%) | 2.1 (27.0%) |
| Male | 7.0 (2.0 - 9.0) | 5.2 (0.0 - 6.0) | 1.9 (26.0%) | 1.2 (17.0%) | 2.3 (33.0%) | 1.7 (24.0%) |
| **Household Income** | | | | | | |
| Less Than $10,000 | 4.2 (1.0 - 6.0) | 2.7 (0.0 - 4.0) | 1.5 (36.0%) | 0.3 (7.0%) | 1.3 (30.0%) | 1.1 (26.0%) |
| $10,000 to $24,999 | 5.4 (1.0 - 7.0) | 3.8 (0.0 - 5.0) | 1.7 (31.0%) | 0.8 (14.0%) | 1.7 (31.0%) | 1.3 (24.0%) |
| $25,000 to $49,999 | 6.2 (2.0 - 7.0) | 4.5 (0.0 - 5.0) | 1.7 (27.0%) | 0.6 (10.0%) | 2.3 (37.0%) | 1.6 (26.0%) |
| $50,000 to $74,999 | 6.5 (2.0 - 9.0) | 4.6 (0.0 - 6.0) | 1.9 (29.0%) | 1.1 (17.0%) | 1.7 (26.0%) | 1.8 (28.0%) |
| $75,000 to $99,999 | 8.4 (3.0 - 10.0) | 6.3 (0.0 - 7.0) | 2.1 (25.0%) | 2.0 (24.0%) | 2.4 (28.0%) | 1.9 (23.0%) |
| $100,000 to $149,999 | 8.3 (3.0 - 10.0) | 6.2 (0.0 - 8.0) | 2.1 (25.0%) | 1.3 (15.0%) | 3.0 (36.0%) | 2.0 (24.0%) |
| $150,000 or more | 8.3 (2.0 - 10.0) | 6.3 (0.0 - 8.0) | 2.1 (25.0%) | 1.4 (17.0%) | 2.6 (31.0%) | 2.2 (27.0%) |
| **5-year Age Group** | | | | | | |
| 0-4 | 6.9 (3.0 - 9.0) | 3.8 (0.0 - 6.0) | 3.1 (45.0%) | 2.0 (29.0%) | 0.0 (0.0%) | 1.8 (26.0%) |
| 5-9 | 10.1 (5.0 - 13.0) | 6.8 (2.0 - 9.0) | 3.3 (33.0%) | 5.0 (49.0%) | 0.0 (0.0%) | 1.8 (18.0%) |
| 10-14 | 8.7 (4.0 - 11.0) | 5.5 (1.0 - 8.0) | 3.2 (37.0%) | 4.3 (50.0%) | 0.0 (0.0%) | 1.2 (13.0%) |
| 15-19 | 10.3 (4.0 - 13.0) | 7.5 (1.0 - 10.0) | 2.8 (27.0%) | 3.8 (37.0%) | 2.0 (20.0%) | 1.6 (16.0%) |
| 20-24 | 7.7 (2.0 - 9.0) | 5.6 (0.0 - 7.0) | 2.1 (27.0%) | 0.7 (9.0%) | 3.6 (46.0%) | 1.4 (18.0%) |
| 25-29 | 6.0 (2.0 - 8.0) | 4.3 (0.0 - 5.0) | 1.7 (28.0%) | 0.4 (7.0%) | 2.7 (46.0%) | 1.2 (20.0%) |
| 30-34 | 8.2 (1.0 - 9.0) | 6.5 (0.0 - 7.0) | 1.7 (21.0%) | 0.6 (7.0%) | 4.2 (51.0%) | 1.6 (20.0%) |
| 35-39 | 8.1 (3.0 - 9.0) | 6.1 (0.0 - 6.0) | 2.1 (26.0%) | 1.1 (14.0%) | 3.4 (42.0%) | 1.6 (19.0%) |
| 40-44 | 7.7 (2.0 - 10.0) | 6.0 (1.0 - 8.0) | 1.7 (22.0%) | 0.2 (2.0%) | 3.6 (46.0%) | 2.2 (29.0%) |
| 45-49 | 7.8 (2.0 - 10.0) | 5.9 (0.0 - 8.0) | 1.9 (24.0%) | 0.8 (11.0%) | 3.5 (45.0%) | 1.5 (19.0%) |
| 50-54 | 7.4 (2.0 - 8.0) | 5.8 (0.0 - 6.0) | 1.7 (23.0%) | 0.3 (4.0%) | 3.8 (50.0%) | 1.7 (23.0%) |
| 55-59 | 8.9 (2.0 - 11.0) | 7.5 (0.0 - 9.0) | 1.5 (16.0%) | 0.7 (7.0%) | 4.8 (54.0%) | 2.0 (23.0%) |
| 60-64 | 5.2 (1.0 - 7.0) | 4.0 (0.0 - 6.0) | 1.3 (24.0%) | 0.0 (0.0%) | 2.1 (40.0%) | 1.9 (36.0%) |
| 65-69 | 5.3 (2.0 - 6.0) | 4.2 (0.0 - 5.0) | 1.1 (21.0%) | 0.0 (0.0%) | 1.6 (31.0%) | 2.5 (48.0%) |
| 70-74 | 4.5 (1.0 - 7.0) | 3.7 (0.0 - 6.0) | 0.8 (18.0%) | 0.0 (0.0%) | 0.8 (17.0%) | 2.9 (64.0%) |
| 75-79 | 5.1 (1.0 - 6.0) | 4.0 (0.0 - 5.0) | 1.0 (21.0%) | 0.0 (0.0%) | 1.0 (19.0%) | 3.1 (60.0%) |
| 80+ | 3.8 (1.0 - 5.0) | 2.8 (0.0 - 4.0) | 1.0 (26.0%) | 0.0 (0.0%) | 0.0 (0.0%) | 2.8 (73.0%) |
| **Ad-hoc Age Group** | | | | | | |
| 0-4 | 6.9 (3.0 - 9.0) | 3.8 (0.0 - 6.0) | 3.1 (45.0%) | 2.0 (29.0%) | 0.0 (0.0%) | 1.8 (26.0%) |
| 5-9 | 10.1 (5.0 - 13.0) | 6.8 (2.0 - 9.0) | 3.3 (33.0%) | 5.0 (49.0%) | 0.0 (0.0%) | 1.8 (18.0%) |
| 10-17 | 9.6 (4.0 - 12.0) | 6.5 (1.0 - 8.0) | 3.1 (32.0%) | 4.4 (45.0%) | 0.7 (8.0%) | 1.4 (15.0%) |
| 18-29 | 7.3 (2.0 - 9.0) | 5.3 (0.0 - 7.0) | 2.0 (28.0%) | 1.0 (13.0%) | 3.0 (41.0%) | 1.3 (18.0%) |
| 30-39 | 8.2 (2.0 - 9.0) | 6.3 (0.0 - 6.0) | 1.9 (24.0%) | 0.9 (11.0%) | 3.8 (46.0%) | 1.6 (20.0%) |
| 40-49 | 7.8 (2.0 - 10.0) | 6.0 (0.0 - 8.0) | 1.8 (23.0%) | 0.4 (6.0%) | 3.6 (46.0%) | 2.0 (25.0%) |
| 50-59 | 8.3 (2.0 - 10.0) | 6.7 (0.0 - 7.0) | 1.6 (19.0%) | 0.5 (6.0%) | 4.3 (52.0%) | 1.9 (23.0%) |
| 60+ | 4.9 (1.0 - 6.0) | 3.8 (0.0 - 5.0) | 1.1 (22.0%) | 0.0 (0.0%) | 1.3 (27.0%) | 2.5 (51.0%) |
| **Household Size** | | | | | | |
| 1 | 4.9 (0.0 - 6.0) | 4.6 (0.0 - 6.0) | 0.3 (6.0%) | 0.2 (4.0%) | 2.7 (55.0%) | 1.7 (35.0%) |
| 2 | 6.1 (1.0 - 7.0) | 5.1 (0.0 - 6.0) | 1.0 (17.0%) | 0.6 (9.0%) | 2.3 (38.0%) | 2.2 (37.0%) |
| 3 | 7.0 (2.0 - 9.0) | 5.1 (0.0 - 7.0) | 1.8 (26.0%) | 0.9 (13.0%) | 2.3 (33.0%) | 1.9 (28.0%) |
| 4 | 9.2 (3.0 - 10.0) | 6.5 (0.0 - 8.0) | 2.7 (29.0%) | 2.2 (24.0%) | 2.7 (29.0%) | 1.7 (18.0%) |
| 5 | 8.6 (4.0 - 10.0) | 5.3 (0.0 - 7.0) | 3.3 (38.0%) | 2.3 (27.0%) | 1.6 (18.0%) | 1.4 (16.0%) |

| | | | | | | |
|---|---|---|---|---|---|---|
| 6 | 9.9 (5.0 - 11.0) | 6.0 (0.0 - 6.0) | 3.9 (40.0%) | 2.4 (24.0%) | 1.7 (18.0%) | 1.8 (19.0%) |
| 7+ | 10.7 (6.0 - 12.0) | 5.8 (0.0 - 6.0) | 4.9 (45.0%) | 1.6 (15.0%) | 2.9 (28.0%) | 1.3 (12.0%) |
| **Day** | | | | | | |
| Weekday | 7.5 (2.0 - 9.0) | 5.6 (0.0 - 7.0) | 1.9 (26.0%) | 1.3 (17.0%) | 2.5 (33.0%) | 1.8 (24.0%) |
| Weekend | 6.7 (2.0 - 8.0) | 4.6 (0.0 - 6.0) | 2.1 (31.0%) | 1.0 (15.0%) | 1.5 (22.0%) | 2.1 (32.0%) |
| **Participant Sick** | | | | | | |
| No | 7.4 (2.0 - 9.0) | 5.5 (0.0 - 7.0) | 1.9 (25.0%) | 1.2 (17.0%) | 2.4 (32.0%) | 1.9 (26.0%) |
| Yes | 6.7 (2.0 - 8.0) | 4.3 (0.0 - 5.0) | 2.4 (36.0%) | 1.0 (15.0%) | 2.0 (30.0%) | 1.3 (19.0%) |
| Refused | 25.7 (1.0 - 41.0) | 24.7 (0.0 - 40.0) | 1.0 (4.0%) | 12.4 (48.0%) | 6.8 (26.0%) | 5.6 (22.0%) |
| **Household Member Sick** | | | | | | |
| No, Not that I know of | 7.3 (2.0 - 9.0) | 5.5 (0.0 - 7.0) | 1.8 (25.0%) | 1.2 (16.0%) | 2.4 (33.0%) | 1.9 (26.0%) |
| Yes | 8.0 (4.0 - 10.0) | 5.4 (1.0 - 6.0) | 2.7 (33.0%) | 1.6 (20.0%) | 1.8 (22.0%) | 2.0 (25.0%) |
| Refused | 0.6 (0.0 - 1.0) | 0.0 (0.0 - 0.0) | 0.6 (100.0%) | 0.0 (0.0%) | 0.0 (0.0%) | 0.0 (0.0%) |
| **Region** | | | | | | |
| Midwest | 8.0 (3.0 - 10.0) | 6.2 (1.0 - 8.0) | 1.9 (23.0%) | 1.4 (18.0%) | 2.4 (30.0%) | 2.3 (29.0%) |
| Northeast | 6.9 (2.0 - 8.0) | 5.0 (0.0 - 6.0) | 1.9 (27.0%) | 1.2 (17.0%) | 2.2 (32.0%) | 1.6 (24.0%) |
| South | 7.4 (2.0 - 9.0) | 5.5 (0.0 - 7.0) | 1.9 (26.0%) | 1.2 (17.0%) | 2.5 (34.0%) | 1.7 (23.0%) |
| West | 7.1 (2.0 - 9.0) | 5.0 (0.0 - 6.0) | 2.0 (29.0%) | 1.0 (14.0%) | 2.1 (29.0%) | 1.9 (28.0%) |
| **Survey Week** | | | | | | |
| May,15 - May,22 | 7.1 (2.0 - 9.0) | 5.3 (0.0 - 7.0) | 1.9 (26.0%) | 1.2 (17.0%) | 2.2 (31.0%) | 1.8 (25.0%) |
| May,22 - May,29 | 6.9 (2.0 - 9.0) | 4.8 (0.0 - 6.0) | 2.1 (30.0%) | 0.6 (9.0%) | 2.5 (36.0%) | 1.7 (25.0%) |
| May,29 - June,05 | 3.7 (1.0 - 6.0) | 1.9 (0.0 - 4.0) | 1.8 (49.0%) | 0.8 (23.0%) | 0.0 (0.0%) | 1.0 (28.0%) |
| October,08-October,15 | 7.8 (2.0 - 10.0) | 5.8 (0.0 - 7.0) | 1.9 (25.0%) | 1.3 (17.0%) | 2.6 (33.0%) | 1.9 (25.0%) |
| October,15-October,22 | 7.0 (3.0 - 9.0) | 5.0 (1.0 - 6.0) | 2.0 (28.0%) | 1.0 (15.0%) | 1.7 (24.0%) | 2.2 (32.0%) |
| October,22-October,29 | 7.7 (2.0 - 10.0) | 5.7 (0.0 - 6.0) | 2.0 (26.0%) | 3.7 (48.0%) | 0.4 (5.0%) | 1.7 (22.0%) |
| **Wave** | | | | | | |
| Wave 1 | 7.1 (2.0 - 9.0) | 5.1 (0.0 - 6.0) | 1.9 (27.0%) | 1.1 (16.0%) | 2.2 (32.0%) | 1.8 (25.0%) |
| Wave 2 | 7.7 (2.0 - 10.0) | 5.7 (0.0 - 7.0) | 1.9 (25.0%) | 1.3 (17.0%) | 2.5 (32.0%) | 2.0 (25.0%) |
| **Industry Group** | | | | | | |
| Arts, Information, Finance, and Professional Services | 5.5 (2.0 - 7.0) | 4.0 (0.0 - 6.0) | 1.5 (28.0%) | 0.1 (3.0%) | 1.9 (35.0%) | 1.9 (35.0%) |
| Education, Tutoring, and Child Care | 16.1 (4.0 - 24.0) | 14.2 (1.0 - 22.0) | 1.9 (12.0%) | 5.2 (32.0%) | 6.3 (39.0%) | 2.8 (17.0%) |
| Factory, Manufacturing, and Woodworking | 8.3 (3.0 - 11.0) | 6.6 (1.0 - 10.0) | 1.6 (20.0%) | 0.0 (0.0%) | 5.7 (68.0%) | 1.0 (12.0%) |
| Health Care | 10.7 (3.0 - 15.0) | 9.1 (0.0 - 13.0) | 1.7 (16.0%) | 0.1 (1.0%) | 7.2 (67.0%) | 1.7 (16.0%) |
| Infrastructure, Logistics, Construction, and Maintenance | 6.9 (3.0 - 9.0) | 5.1 (1.0 - 6.0) | 1.8 (26.0%) | 0.0 (0.0%) | 3.3 (48.0%) | 1.8 (26.0%) |

| | | | | | | |
|---|---|---|---|---|---|---|
| Sales and Support Services | 9.4 (3.0 - 11.0) | 7.7 (0.0 - 9.0) | 1.8 (19.0%) | 0.3 (3.0%) | 5.6 (59.0%) | 1.8 (19.0%) |
| Minor | 8.9 (4.0 - 11.0) | 5.8 (1.0 - 8.0) | 3.1 (35.0%) | 3.8 (43.0%) | 0.3 (4.0%) | 1.6 (18.0%) |
| Not Asked or Refused | 4.0 (1.0 - 5.0) | 2.6 (0.0 - 3.0) | 1.5 (37.0%) | 0.2 (4.0%) | 0.4 (9.0%) | 2.0 (50.0%) |
| Other | 7.1 (2.0 - 9.0) | 5.7 (0.0 - 7.0) | 1.5 (20.0%) | 0.0 (0.0%) | 3.7 (51.0%) | 2.0 (28.0%) |
| **Occupation Group** | | | | | | |
| Arts, IT, Mathematical, and Physical, Life or Social Sciences | 5.0 (2.0 - 7.0) | 3.3 (0.0 - 5.0) | 1.6 (33.0%) | 0.3 (6.0%) | 1.5 (31.0%) | 1.5 (31.0%) |
| Building Cleaning, Maintainance and Repair | 7.4 (3.0 - 9.0) | 5.3 (0.0 - 7.0) | 2.1 (28.0%) | 0.0 (0.0%) | 4.4 (59.0%) | 0.9 (13.0%) |
| Business Operations | 6.5 (2.0 - 9.0) | 5.2 (0.0 - 7.0) | 1.3 (20.0%) | 0.0 (0.0%) | 3.5 (53.0%) | 1.8 (27.0%) |
| Construction, Architechture, Engineering and Precision Production | 6.3 (3.0 - 9.0) | 4.6 (2.0 - 6.0) | 1.7 (27.0%) | 0.0 (0.0%) | 2.8 (45.0%) | 1.7 (28.0%) |
| Education, Training, and Library | 19.2 (6.0 - 29.0) | 17.1 (5.0 - 27.0) | 2.1 (11.0%) | 5.7 (30.0%) | 7.8 (41.0%) | 3.6 (19.0%) |
| Financial Operations and Services | 6.2 (3.0 - 8.0) | 4.2 (0.0 - 5.0) | 2.0 (32.0%) | 0.0 (1.0%) | 2.3 (37.0%) | 1.9 (31.0%) |
| Food Preparation and Serving | 11.2 (3.0 - 12.0) | 8.6 (1.0 - 12.0) | 2.5 (23.0%) | 2.0 (18.0%) | 4.5 (40.0%) | 2.1 (19.0%) |
| Health Diagnosing or Treating Practitioner | 17.4 (5.0 - 33.0) | 15.7 (2.0 - 29.0) | 1.7 (10.0%) | 0.8 (4.0%) | 13.1 (75.0%) | 1.9 (11.0%) |
| Health Technicians and Care Support | 9.8 (4.0 - 19.0) | 7.9 (2.0 - 15.0) | 2.0 (20.0%) | 0.2 (2.0%) | 5.7 (58.0%) | 2.0 (20.0%) |
| Not Asked or Refused | 4.0 (1.0 - 5.0) | 2.6 (0.0 - 4.0) | 1.5 (37.0%) | 0.2 (4.0%) | 0.4 (9.0%) | 2.0 (50.0%) |
| Office and Administrative Support, Management and Legal | 8.0 (1.0 - 11.0) | 6.6 (0.0 - 9.0) | 1.4 (18.0%) | 0.1 (1.0%) | 4.5 (57.0%) | 1.9 (24.0%) |
| Other | 7.1 (2.0 - 10.0) | 5.6 (0.0 - 7.0) | 1.5 (21.0%) | 0.4 (5.0%) | 3.4 (47.0%) | 1.9 (26.0%) |
| Sales and Personal Care Services | 10.1 (3.0 - 13.0) | 8.4 (0.0 - 12.0) | 1.6 (16.0%) | 0.4 (4.0%) | 6.5 (64.0%) | 1.6 (16.0%) |
| Transportation, Moving, Community and Social Services | 6.0 (1.0 - 9.0) | 4.6 (1.0 - 6.0) | 1.5 (24.0%) | 0.3 (5.0%) | 2.7 (44.0%) | 1.6 (26.0%) |

[1] Mean number of contacts (50% IQR)
[2] Mean number of contacts (percentage of the number of total contacts)

#### 2.4 Association Between Respondent Characteristics and Number of Non-Household Contacts

When focusing on the results of the multivariable analysis of non-household contacts (Table S 12), we identify several significant predictors, with the best-fitting model (according to AIC) accounting for 24.3% of the variance ($R^2 = 0.243$).

Main activity and attendance mode remain the most powerful predictors of non-household contacts. Compared to students at school, those working or studying fully online had dramatically fewer non-household contacts—reductions ranged from 72% for employed individuals (RR=0.28, p < 0.001) to 86% for students (RR = 0.14, p < 0.001). Those physically present at a school or workplace generally had the most non-household contacts, except for in-person self-employed individuals, who have 42% fewer reported contacts (RR = 0.58, p = 0.017) than students.

The non-linear relationship with age is also significant for non-household contacts, following a similar wavelike pattern as seen for total contacts (Figure S 1). The difference of non-household contact dynamics lies in the lower magnitude of the contacts in childhood and young adulthood with respect to those after 60 years. The only statistically significant difference observed between age groups is for those aged 15–19 years and 25–29 years, with the latter reporting 37% fewer non-household contacts (RR = 0.63, p = 0.017).

Among other statistically significant correlations, females continue to have more non-household contacts than males (RR = 1.29, p < 0.001). The gradient with household income also persisted, with lower-income groups reporting significantly fewer non-household contacts than the highest-income group. For example, the lowest income group (< $10,000) had 40% fewer non-household contacts (RR=0.60, p=0.004) than the highest-income group. Finally, different from total contacts, being sick was associated with a statistically significant 19% reduction in non-household contacts (RR=0.80, p=0.027).

Isolating non-household contacts reveals several crucial differences from the analysis of total contacts, primarily by removing the strong, mechanical effect of living with other people. The most dramatic and expected difference is the household size, which is not a statistically significant predictor of non-household contacts. This confirms that its role in the total contacts model was almost entirely driven by counting interactions between people who live together.

Other links are illustrated to be either weaker or stronger for non-household contacts. For example, the effect of being female is stronger for non-household contacts. While being female was associated with 21% more total contacts, it was associated with 29% more non-household contacts. This suggests that the gender gap in contact patterns is primarily driven by interactions outside the home. The effect of being sick is clearer and more significant for non-household contacts.

**Table S 12.** Results of multivariable GAM analysis for non-household contacts.

| | **Best AIC [1]** | | **Best $R^2$ [2]** | | **Best % Deviance [3]** | | **Main Activity only [4]** | | **Ad-hoc Age Group [5]** | |
|---|---|---|---|---|---|---|---|---|---|---|
| **Characteristic** | **RR (95% CI)[1]** | **p-value** | **RR (95% CI)[1]** | **p-value** | **RR (95% CI)[1]** | **p-value** | **RR (95% CI)[1]** | **p-value** | **RR (95% CI)[1]** | **p-value** |
| **Main Activity and Attendance Mode** | | | | | | | | | | |
| Student: At school in person | ref. | | ref. | | ref. | | | | ref. | |
| Employed: At work in person | 1.27 (0.95, 1.69) | 0.10 | 1.20 (0.99, 1.47) | 0.070 | 1.38* (1.01, 1.89) | **0.040** | | | 1.29 (0.95, 1.73) | 0.10 |
| Self-Employed: At work in person | 0.58* (0.37, 0.91) | **0.017** | 0.55** (0.37, 0.82) | **0.004** | 0.64 (0.40, 1.02) | 0.058 | | | 0.56* (0.36, 0.89) | **0.014** |

| | | | | | | | | | | |
|---|---|---|---|---|---|---|---|---|---|---|
| Student & (Self-)Employed: At work/school in person | 1.46 (1.00, 2.13) | 0.050 | 1.31 (0.92, 1.86) | 0.14 | 1.47 (0.99, 2.19) | 0.059 | | | 1.55* (1.04, 2.29) | **0.030** |
| Student: Not at school | 0.22*** (0.16, 0.31) | **<0.001** | 0.22*** (0.16, 0.31) | **<0.001** | 0.22*** (0.16, 0.31) | **<0.001** | | | 0.23*** (0.16, 0.31) | **<0.001** |
| Employed: Not at work | 0.35*** (0.25, 0.49) | **<0.001** | 0.33*** (0.25, 0.43) | **<0.001** | 0.37*** (0.25, 0.53) | **<0.001** | | | 0.34*** (0.24, 0.49) | **<0.001** |
| Self-Employed: Not at work | 0.20*** (0.11, 0.36) | **<0.001** | 0.21*** (0.12, 0.38) | **<0.001** | 0.22*** (0.12, 0.41) | **<0.001** | | | 0.21*** (0.12, 0.39) | **<0.001** |
| Student & (Self-)Employed: Not at work/school | 0.46** (0.26, 0.79) | **0.005** | 0.41*** (0.24, 0.69) | **<0.001** | 0.45** (0.25, 0.80) | **0.007** | | | 0.48* (0.27, 0.85) | **0.011** |
| Other: Not student or employed | 0.21*** (0.16, 0.28) | **<0.001** | 0.24*** (0.19, 0.29) | **<0.001** | 0.23*** (0.17, 0.30) | **<0.001** | | | 0.23*** (0.17, 0.30) | **<0.001** |
| Student: Studied fully online | 0.14*** (0.06, 0.36) | **<0.001** | 0.13*** (0.05, 0.32) | **<0.001** | 0.16*** (0.06, 0.41) | **<0.001** | | | 0.15*** (0.06, 0.38) | **<0.001** |
| Employed: Worked fully online | 0.28*** (0.20, 0.39) | **<0.001** | 0.26*** (0.20, 0.34) | **<0.001** | 0.29*** (0.20, 0.41) | **<0.001** | | | 0.27*** (0.19, 0.38) | **<0.001** |
| Self-Employed: Worked fully online | 0.20*** (0.11, 0.38) | **<0.001** | 0.19*** (0.11, 0.35) | **<0.001** | 0.23*** (0.12, 0.43) | **<0.001** | | | 0.20*** (0.11, 0.38) | **<0.001** |
| Student & (Self-)Employed: Worked/studied fully online | 0.21** (0.07, 0.61) | **0.004** | 0.19** (0.07, 0.54) | **0.002** | 0.24** (0.08, 0.70) | **0.009** | | | 0.21** (0.07, 0.62) | **0.004** |
| **Main Activity** | | | | | | | | | | |
| student | | | | | | | ref. | | | |
| employed | | | | | | | 1.34 (0.98, 1.82) | 0.064 | | |
| self-employed | | | | | | | 0.58** (0.38, 0.87) | **0.009** | | |
| student & (self-)employed | | | | | | | 1.54* (1.06, 2.24) | **0.022** | | |
| other | | | | | | | 0.31*** (0.23, 0.42) | **<0.001** | | |
| **Sex** | | | | | | | | | | |
| Male | ref. | | ref. | | ref. | | ref. | | ref. | |
| Female | 1.29*** (1.15, 1.45) | **<0.001** | 1.28*** (1.14, 1.44) | **<0.001** | 1.31*** (1.16, 1.48) | **<0.001** | 1.25*** (1.10, 1.43) | **<0.001** | 1.28*** (1.14, 1.44) | **<0.001** |
| **Sick** | | | | | | | | | | |
| No | ref. | | ref. | | ref. | | ref. | | ref. | |
| Yes | 0.80* (0.66, 0.97) | **0.032** | 0.79* (0.64, 0.96) | **0.018** | 0.81* (0.67, 1.00) | **0.045** | 0.76* (0.62, 0.95) | **0.014** | 0.82 (0.67, 1.01) | 0.059 |
| **Household Income** | | | | | | | | | | |
| $150,000 or more | ref. | | ref. | | ref. | | ref. | | ref. | |
| Less than $10,000 | 0.60** (0.43, 0.85) | **0.004** | 0.56** (0.40, 0.79) | **0.001** | 0.58** (0.41, 0.83) | **0.003** | 0.65* (0.44, 0.94) | **0.023** | 0.60** (0.42, 0.85) | **0.004** |
| $10,000 to $24,999 | 0.68** (0.53, 0.89) | **0.004** | 0.66** (0.51, 0.86) | **0.002** | 0.67** (0.51, 0.87) | **0.003** | 0.79 (0.59, 1.05) | 0.10 | 0.67** (0.51, 0.87) | **0.003** |
| $25,000 to $49,999 | 0.71*** (0.58, 0.86) | **<0.001** | 0.72** (0.59, 0.88) | **0.001** | 0.69*** (0.57, 0.85) | **<0.001** | 0.80* (0.65, 1.00) | **0.049** | 0.72*** (0.59, 0.87) | **<0.001** |
| $50,000 to $74,999 | 0.80* (0.66, 0.97) | **0.021** | 0.80* (0.66, 0.96) | **0.020** | 0.78* (0.65, 0.95) | **0.014** | 0.90 (0.73, 1.11) | 0.3 | 0.81* (0.67, 0.98) | **0.031** |
| $75,000 to $99,999 | 0.89 (0.73, 1.08) | 0.2 | 0.88 (0.72, 1.07) | 0.2 | 0.87 (0.72, 1.06) | 0.2 | 0.95 (0.77, 1.17) | 0.6 | 0.88 (0.72, 1.08) | 0.2 |

| | | | | | | | | | | |
|---|---|---|---|---|---|---|---|---|---|---|
| $100,000 to $149,999 | 0.97 (0.82, 1.15) | 0.7 | 0.99 (0.83, 1.17) | 0.9 | 0.98 (0.82, 1.16) | 0.8 | 0.99 (0.82, 1.19) | >0.9 | 0.98 (0.83, 1.17) | 0.9 |
| **Age (spline)** | | **<0.001** | | | | | | | | |
| **Race/Ethnicity** | | | | | | | | | | |
| White, NH | | | ref. | | ref. | | ref. | | ref. | |
| Hispanic/Latino | 0.92 (0.79, 1.07) | 0.3 | 0.91 (0.78, 1.06) | 0.2 | 0.96 (0.81, 1.13) | 0.6 | 0.99 (0.83, 1.18) | >0.9 | 0.95 (0.81, 1.12) | 0.5 |
| Black, NH | 0.85 (0.70, 1.02) | 0.086 | 0.83 (0.69, 1.01) | 0.062 | 0.88 (0.72, 1.07) | 0.2 | 0.83 (0.67, 1.03) | 0.10 | 0.85 (0.70, 1.04) | 0.11 |
| Asian, NH | 0.70** (0.55, 0.91) | **0.007** | 0.69** (0.53, 0.89) | **0.004** | 0.77 (0.59, 1.00) | 0.053 | 0.73* (0.55, 0.98) | **0.038** | 0.74* (0.56, 0.97) | **0.027** |
| Other, NH | 0.90 (0.70, 1.18) | 0.5 | 0.89 (0.68, 1.15) | 0.4 | 0.94 (0.72, 1.23) | 0.6 | 0.85 (0.63, 1.13) | 0.3 | 0.92 (0.70, 1.20) | 0.5 |
| **5-year Age Group** | | | | | | | | | | |
| 0-4 | | | | | 0.89 (0.64, 1.25) | 0.5 | 0.95 (0.66, 1.36) | 0.8 | | |
| 5-9 | | | | | 1.07 (0.77, 1.50) | 0.7 | 1.13 (0.79, 1.62) | 0.5 | | |
| 10-14 | | | | | 0.81 (0.58, 1.14) | 0.2 | 0.87 (0.61, 1.26) | 0.5 | | |
| 15-19 | | | | | ref. | | ref. | | | |
| 20-24 | | | | | 0.74 (0.52, 1.05) | 0.10 | 0.70 (0.48, 1.03) | 0.068 | | |
| 25-29 | | | | | 0.63* (0.44, 0.92) | **0.017** | 0.62* (0.42, 0.92) | **0.018** | | |
| 30-34 | | | | | 0.81 (0.56, 1.18) | 0.3 | 0.87 (0.58, 1.30) | 0.5 | | |
| 35-39 | | | | | 0.86 (0.60, 1.25) | 0.4 | 0.86 (0.58, 1.27) | 0.4 | | |
| 40-44 | | | | | 1.01 (0.70, 1.45) | >0.9 | 0.92 (0.62, 1.35) | 0.7 | | |
| 45-49 | | | | | 0.77 (0.52, 1.13) | 0.2 | 0.81 (0.53, 1.24) | 0.3 | | |
| 50-54 | | | | | 0.88 (0.60, 1.28) | 0.5 | 0.85 (0.56, 1.27) | 0.4 | | |
| 55-59 | | | | | 0.91 (0.63, 1.32) | 0.6 | 0.96 (0.64, 1.43) | 0.8 | | |
| 60-64 | | | | | 0.75 (0.52, 1.10) | 0.14 | 0.77 (0.51, 1.16) | 0.2 | | |
| 65-69 | | | | | 1.19 (0.80, 1.76) | 0.4 | 1.15 (0.75, 1.76) | 0.5 | | |
| 70-74 | | | | | 1.28 (0.85, 1.93) | 0.2 | 1.33 (0.86, 2.07) | 0.2 | | |
| 75-79 | | | | | 1.49 (0.95, 2.32) | 0.079 | 1.47 (0.91, 2.38) | 0.12 | | |
| 80+ | | | | | 1.20 (0.74, 1.94) | 0.5 | 1.16 (0.69, 1.96) | 0.6 | | |
| **Ad-hoc Age Group** | | | | | | | | | | |
| 0-4 | | | | | | | | | 0.97 (0.71, 1.32) | 0.8 |
| 5-9 | | | | | | | | | 1.15 (0.85, 1.56) | 0.3 |
| 10-17 | | | | | | | | | ref. | |
| 18-29 | | | | | | | | | 0.80 | 0.2 |

| | | | | | | | | | | |
|---|---|---|---|---|---|---|---|---|---|---|
| | | | | | | | | | (0.60, 1.08) | |
| 30-39 | | | | | | | | | 0.95 (0.69, 1.33) | 0.8 |
| 40-49 | | | | | | | | | 1.04 (0.75, 1.45) | 0.8 |
| 50-59 | | | | | | | | | 1.03 (0.74, 1.44) | 0.9 |
| 60+ | | | | | | | | | 1.21 (0.88, 1.67) | 0.2 |
| **Day** | | | | | | | | | | |
| Weekday | | | | | ref. | | ref. | | ref. | |
| Weekend | | | | | 1.11 (0.93, 1.32) | 0.2 | 0.78** (0.65, 0.94) | **0.009** | 1.08 (0.91, 1.29) | 0.4 |
| **Household Size** | | | | | | | | | | |
| 1 | | | 1.07 (0.85, 1.35) | 0.5 | 0.94 (0.74, 1.19) | 0.6 | 0.97 (0.75, 1.25) | 0.8 | 0.98 (0.77, 1.23) | 0.8 |
| 2 | | | 1.20* (1.01, 1.42) | **0.035** | 1.10 (0.92, 1.31) | 0.3 | 1.10 (0.90, 1.33) | 0.4 | 1.12 (0.94, 1.33) | 0.2 |
| 3 | | | ref. | | ref. | | ref. | | ref. | |
| 4 | | | 1.07 (0.88, 1.28) | 0.5 | 1.01 (0.84, 1.22) | >0.9 | 1.08 (0.88, 1.32) | 0.5 | 1.04 (0.86, 1.26) | 0.7 |
| 5 | | | 0.88 (0.71, 1.10) | 0.3 | 0.87 (0.70, 1.09) | 0.2 | 0.93 (0.73, 1.19) | 0.6 | 0.88 (0.71, 1.11) | 0.3 |
| 6 | | | 0.97 (0.70, 1.34) | 0.9 | 0.97 (0.70, 1.34) | 0.9 | 1.07 (0.75, 1.52) | 0.7 | 0.99 (0.71, 1.36) | >0.9 |
| 7+ | | | 1.11 (0.80, 1.55) | 0.5 | 1.11 (0.80, 1.56) | 0.5 | 1.25 (0.87, 1.80) | 0.2 | 1.10 (0.79, 1.55) | 0.6 |
| **Additional control variables** (not statistically significant) | Household Member Sick | | | | Household Member Sick, Region, Survey Week | | Household Member Sick, Region, Survey Week | | Household Member Sick, Region, Survey Week | |
| **AIC** | **9,739** | | **9,758** | | **9,766** | | **10,043** | | **9,769** | |
| **$R^2$** | **0.243** | | **0.247** | | **0.232** | | **0.108** | | **0.234** | |

The goodness-of-fit measure or the variable distinguishing selected specification is indicated in the column name. [1]*p<0.05; **p<0.01; ***p<0.001. Abbreviations: CI = Confidence Interval

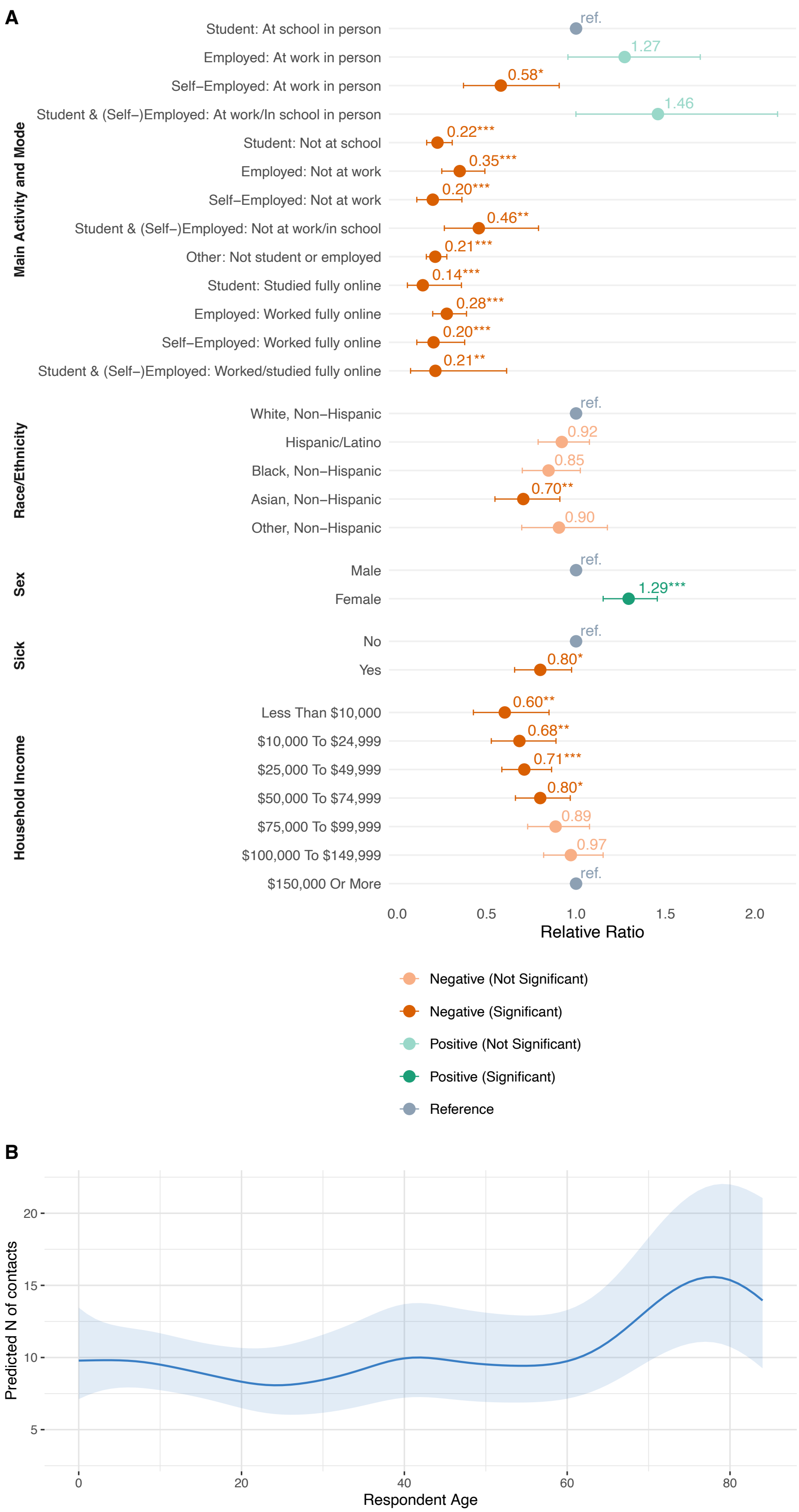


**Figure S 2.** **A** Relative ratios of the number of non-household contacts by population group relative to NH White, male, healthy in-person students from 3-person households with income of $150,000 and more. **B** Predicted number of contacts by age (marginal effects) for the reference group.

#### 2.5 Contact Matrices by Age

Our estimated age-stratified contact matrices for the average number of total contacts are shown in Figure S 3A-B.

For total contacts there is strong assortativity, especially for younger age groups due to school contacts (Figure S 3A). For household-specific contacts, there are three classical diagonals

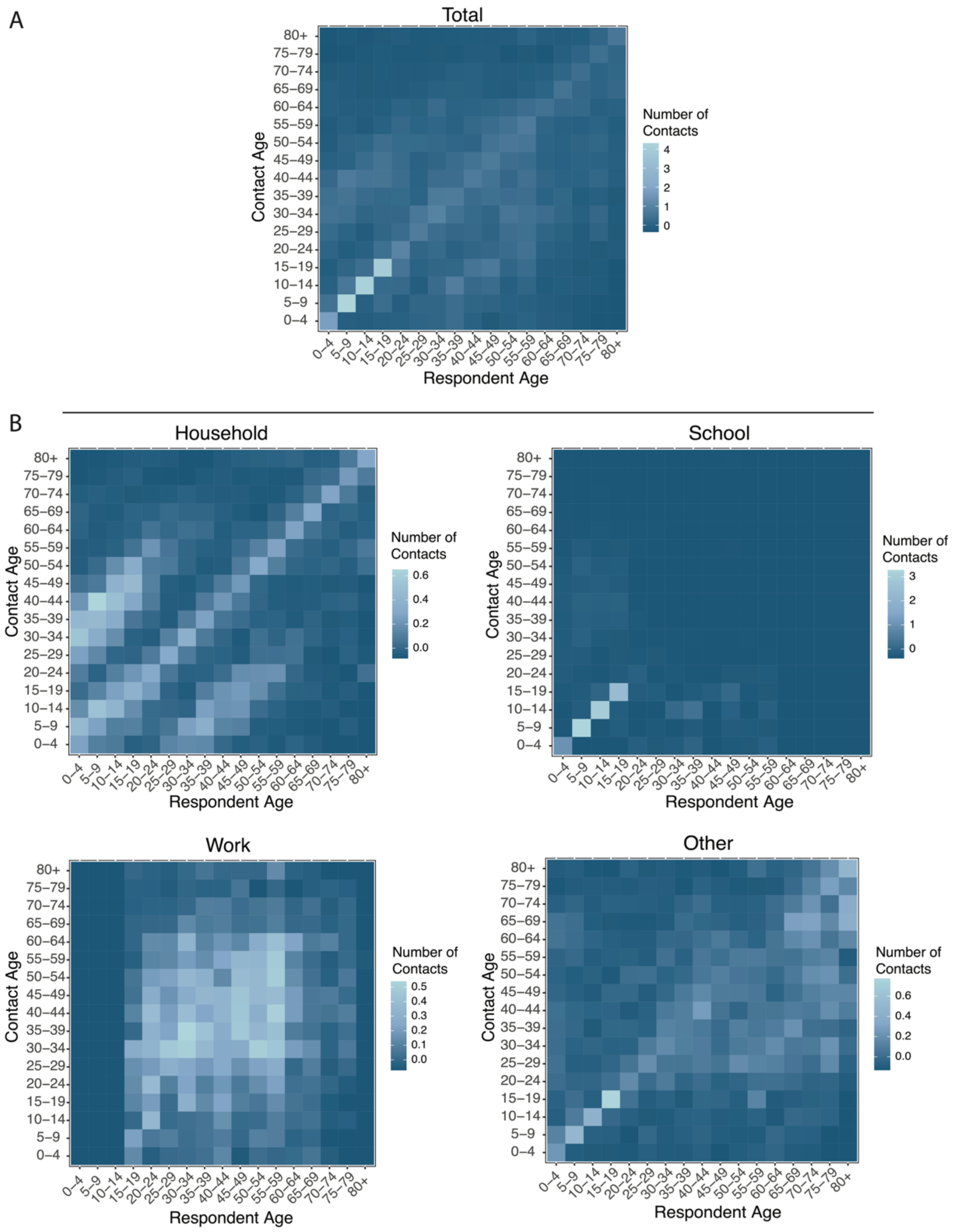


**Figure S 3. A.** Age-stratified contact matrix for total contacts. **B.** As A, but setting-specific.

representing contacts between spouses and siblings (main diagonal), between children with their parents (upper diagonal), and between parents with their children (lower diagonal) (Figure S 3B). For school-specific contacts, there is high assortativity among the school-age population, specifically among individuals aged 5-19 (Figure S 3B). For work-specific contacts, there is more homogeneous mixing between work age groups (Figure S 3B). For other contacts, there is strong assortativity, especially for younger age groups (Figure S 3B).

Average assortativity of contacts by age group for different contact settings is illustrated in Figure S 4. To calculate assortativity for work and school contacts by age, only a subset of the total matrix was used. For workplace, ages were limited to age groups including exclusively respondents and contacts between 18 and 65 years of age. Similarly, for school, age groups were limited to those comprised of respondents and contacts aged 5 – 18 years.

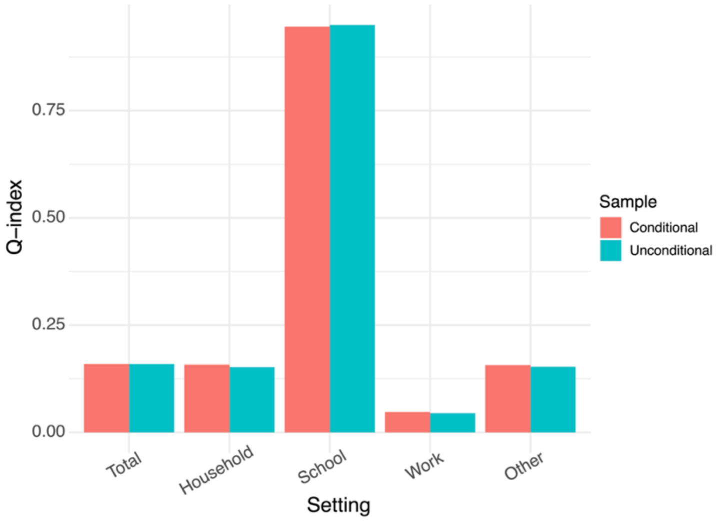


**Figure S 4.** Assortativity of total contacts by age in each setting.

##### *2.5.1 Frequency-Based Contact Matrices*

Frequency-based contact matrices are shown in Figure S 5. For household-specific contacts, there are three diagonal lines representing contacts between spouses and siblings (main diagonal), between children with their parents (upper diagonal), and between parents with their children (lower diagonal) (Figure S 5). For school-specific and work-specific contacts, age groups with extremely small number of respondents were excluded from this computation. Among included school and work ages, we observe high assortativity as well as for other contacts, especially among younger age groups.

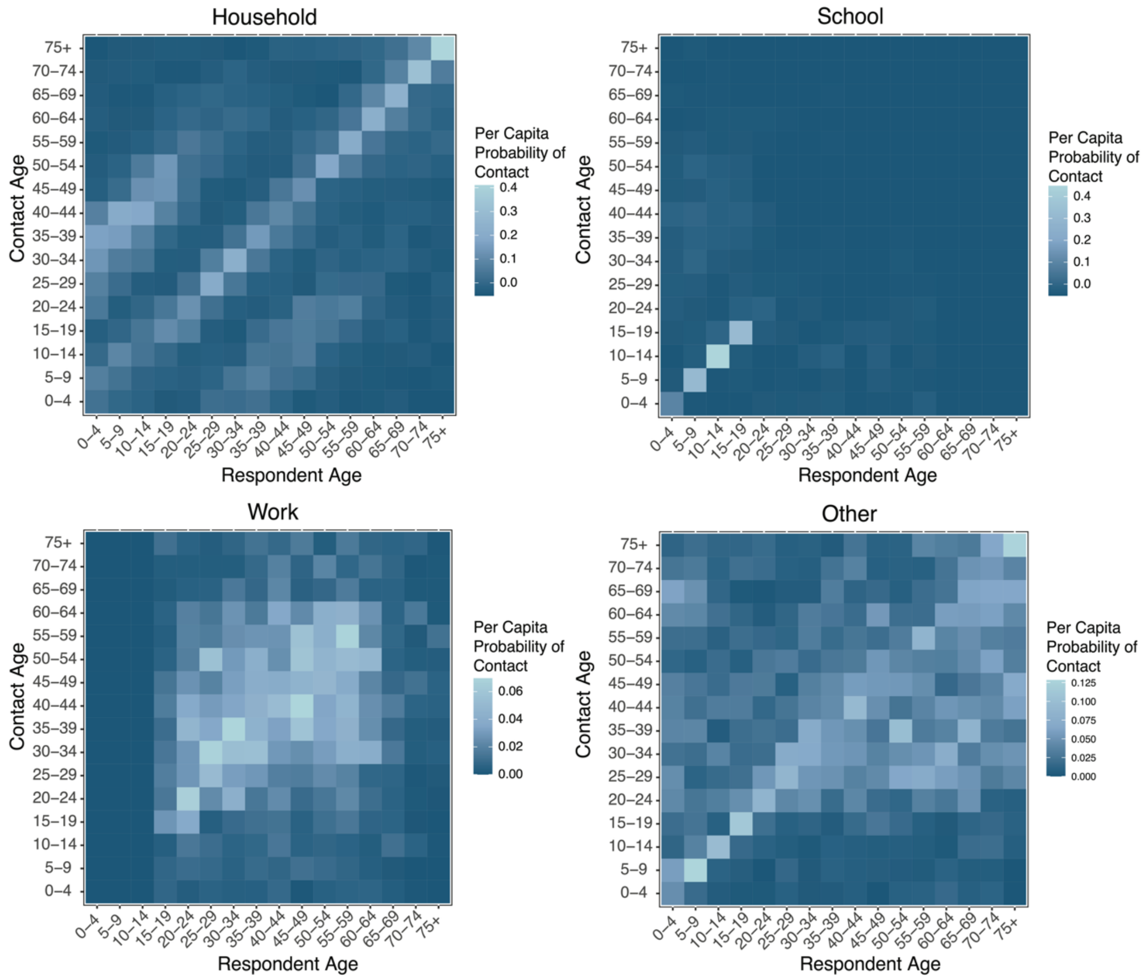


**Figure S 5.** US post-pandemic frequency-based contact matrices.

##### *2.5.2 Conditional Contact Matrices*

Our estimated conditional age-stratified contact matrices for the average number of total contacts are illustrated in Figure S 6. We observe high assortativity for school-specific conditional matrix (number of contacts in a setting conditional on the respondent having at least one contact in that setting, see Section 1.12 for methods), as well as high number of contacts for educator respondents. The rest of the contact patterns are very similar to the main analysis.

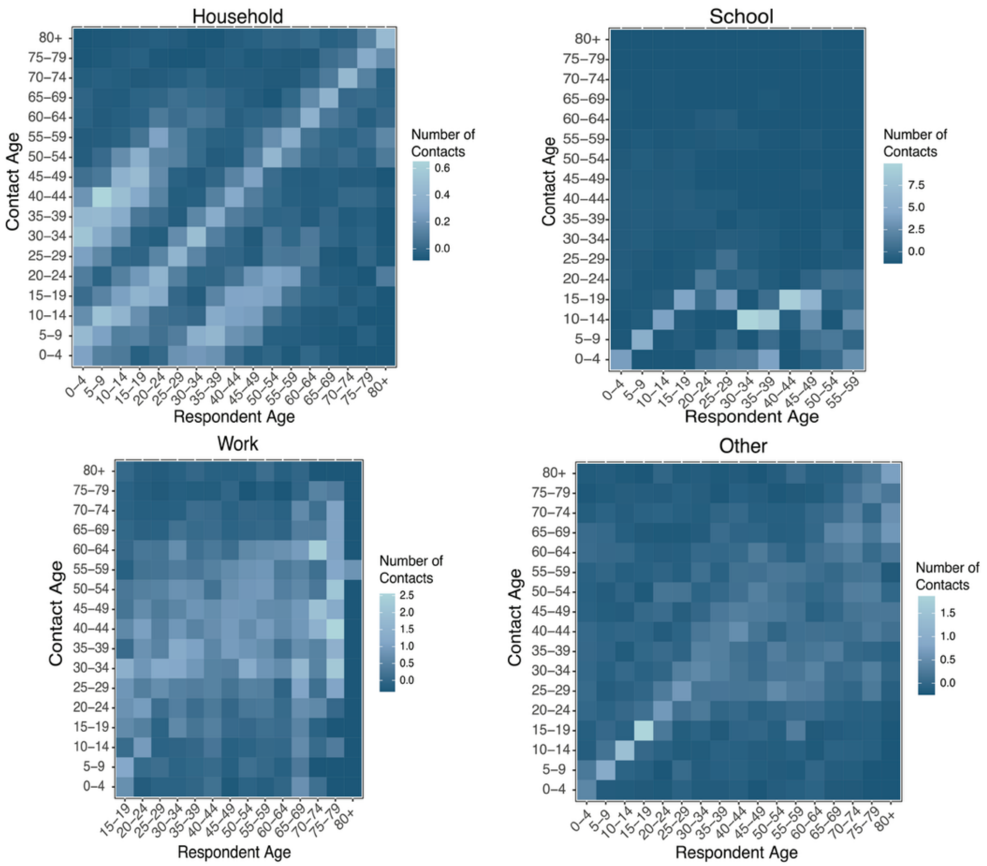


**Figure S 6.** Setting-specific age-stratified contact matrix of the number of contacts ($\Theta_{ij}^{k}$) conditional on the respondent having at least one contact in that setting.

##### *2.5.3 Symmetrized Contact Matrices*

The symmetrized matrices were created by combining a smoothed age-stratified contact matrix (M) with its transpose ($M^T$). While the diagonal remains the same, the off-diagonal estimates change. Estimated symmetrized age-stratified contact matrices are presented in Figure S **7**A-B, illustrating contacts between spouses and siblings (main diagonal) as well as between parents and children (lower and upper diagonals). Among school-specific contacts, there is high assortativity among students, while work contacts are more homogenous. For other contacts, there we observe high assortativity for all ages.

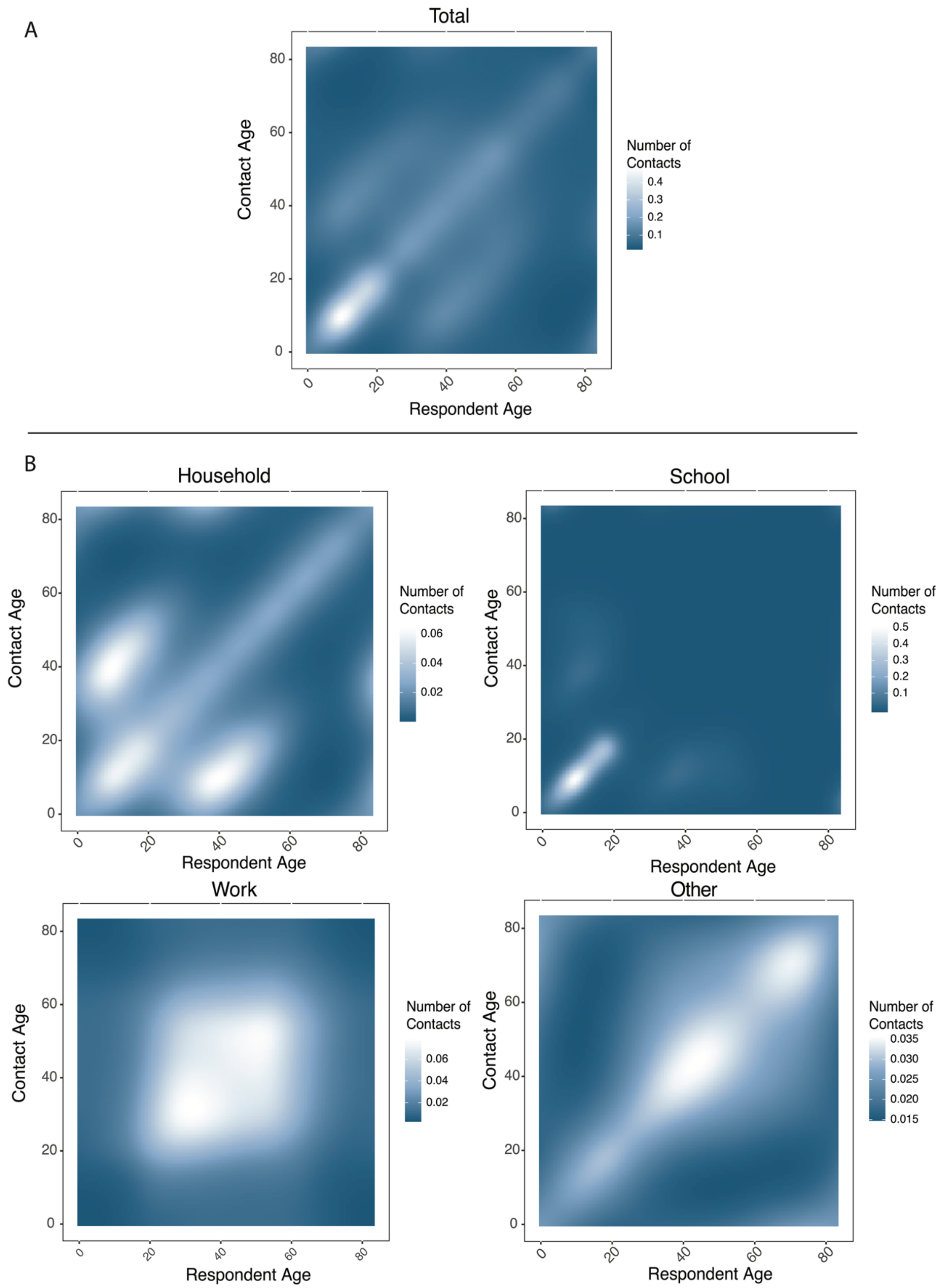


**Figure S 7 A.** Age-stratified symmetrized contact matrices of the number of contacts occurring in the total sample, **B.** As A, but setting-specific. Each element of the matrix is the estimated average number of contacts resulting from bivariate smoothing using GAM model with tensor-product regression spline function of the respondent and contact age (in years).

##### *2.5.4 Age-Stratified Contact Matrices for Alternative Scenarios: In-Person and Remote*

Figure S 8A-B presents two alternative scenarios: i) all working/studying individuals are at work/school in person; ii) all working/studying individuals are either at work/school remotely or not at work/school (not in-person). By applying the contact patterns of the in-person respondents to all working/studying individuals, we observe higher assortativity and relative difference between age groups (Figure S 8A). By applying "not in-person" contact patterns, which combine the contact patterns of those who are fully remote and those who are not at school or at work, we observe lower relative differences between age groups (Figure S 8B). These matrices could potentially be utilized in scenario analysis modeling exercises.

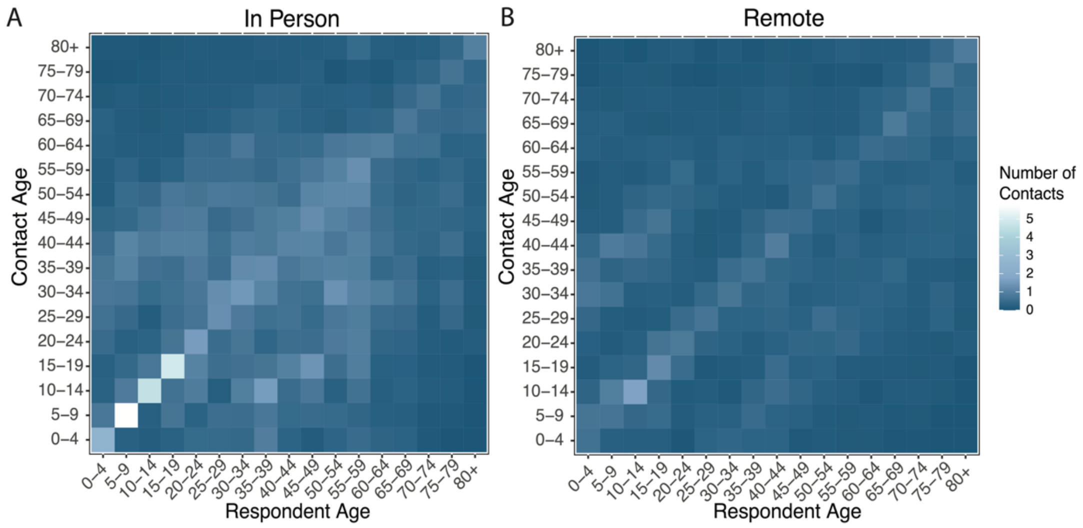


**Figure S 8.** Age-stratified contact matrices by working and/or studying in person vs. remote. Each element of the matrix is the estimated average number of contacts assuming that all working and/or studying participants are working and/or studying fully in person (A) or fully remote (B).

Figure S 9 presents the average number of total contacts by attendance modes and age group. We do not observe any trends in the differences between age groups of in-person individuals, while the average number of contacts for those not in person and those who are not students nor employed follow a wavelike pattern with the highest numbers observed for school-age individuals and those in their 40s.

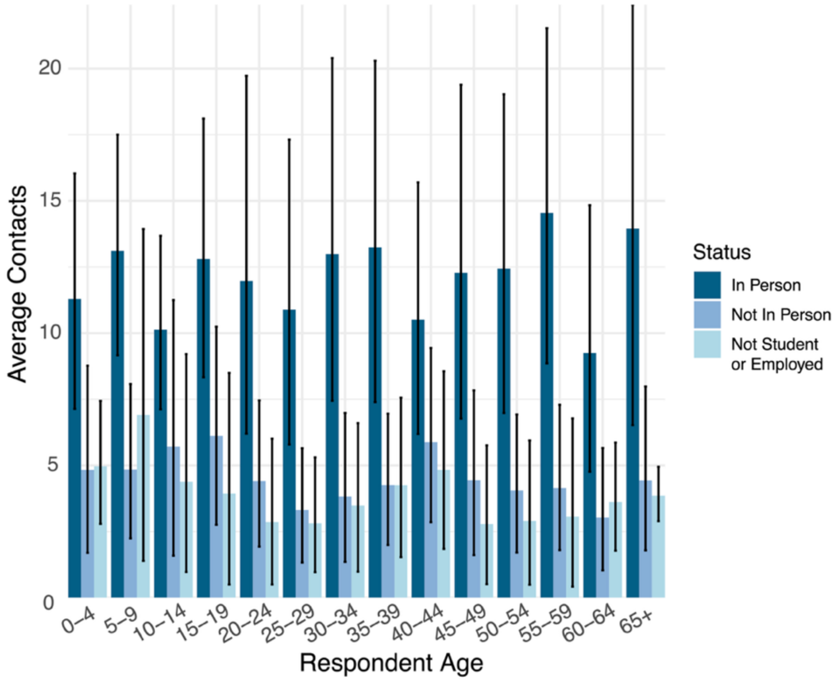


**Figure S 9.** Average number of contacts by age group and mode of participation in the main activity. Error bars show the uncertainty of the weighted mean obtained based on 500 bootstrap samples for each age group**.**

##### *2.5.5 Sensitivity Analysis: Unweighted Sample Analysis*

In addition to age-stratified contact matrices estimated from the weighted data, we also compute age-stratified contact matrices based on unweighted data (without applying respondent weights). The types of contact matrices show strong positive correlations with Pearson's r varying between 0.990 and 0.999 (Figure S 10A-B). In fact, the weighted and unweighted age-stratified contact matrices for the total sample and by setting are almost identical (Figure S 11-Figure S 12A-D), both for the total and for the setting-specific matrices.

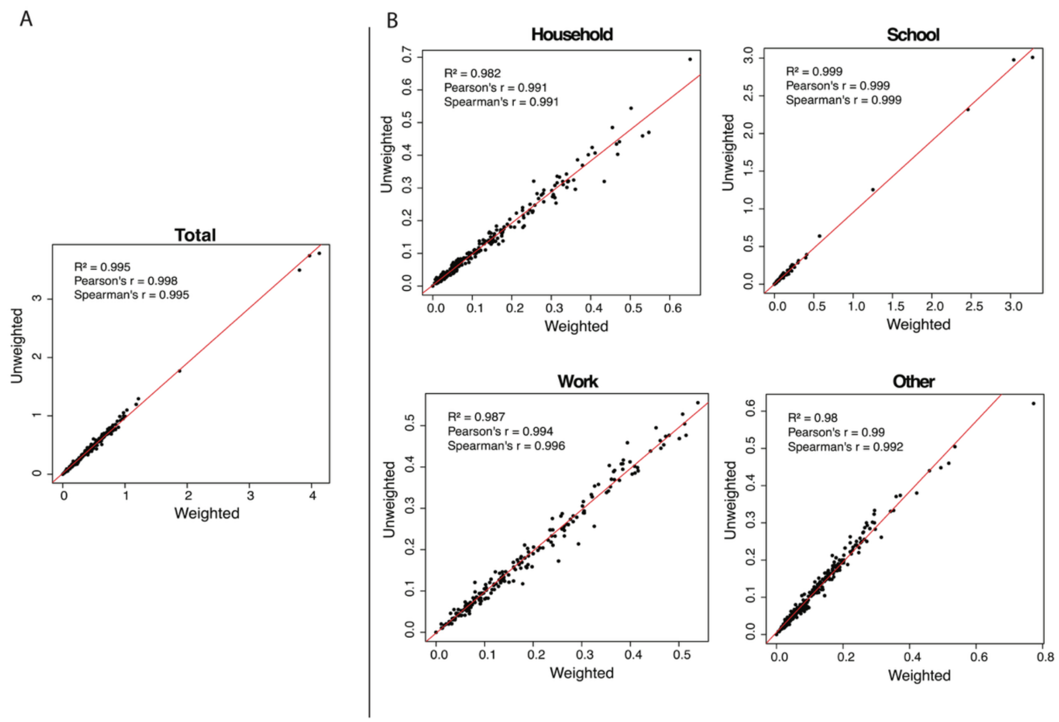


**Figure S 10. A.** Comparison of our estimated contact matrices for the weighted data with the unweighted data for the total sample. **B.** As A, but setting-specific.

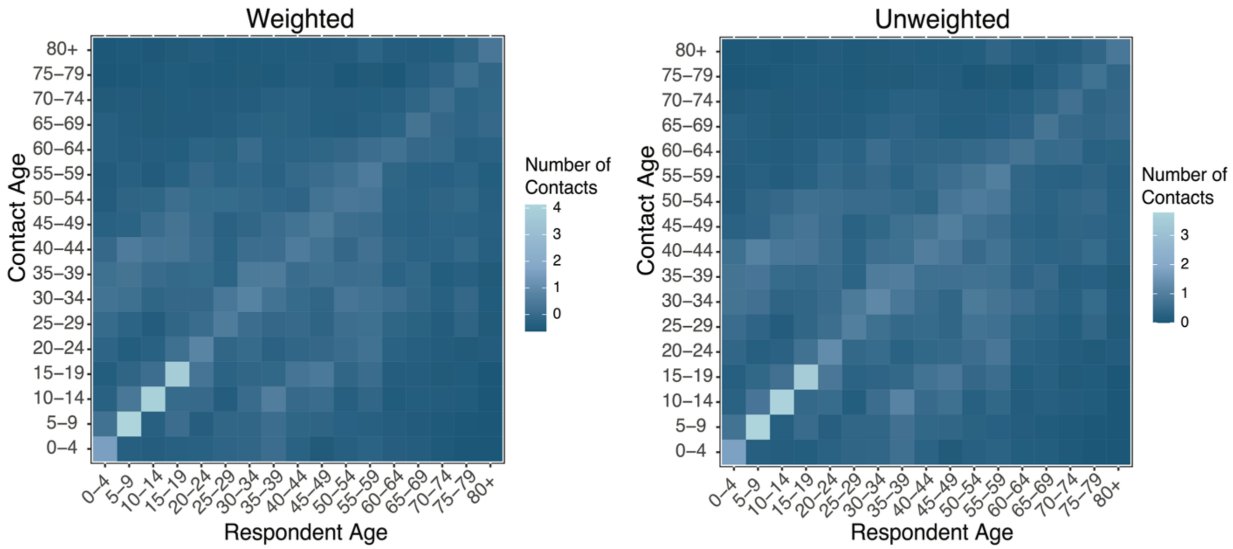


**Figure S 11.** Age-stratified contact matrices for the unweighted and weighted total sample.

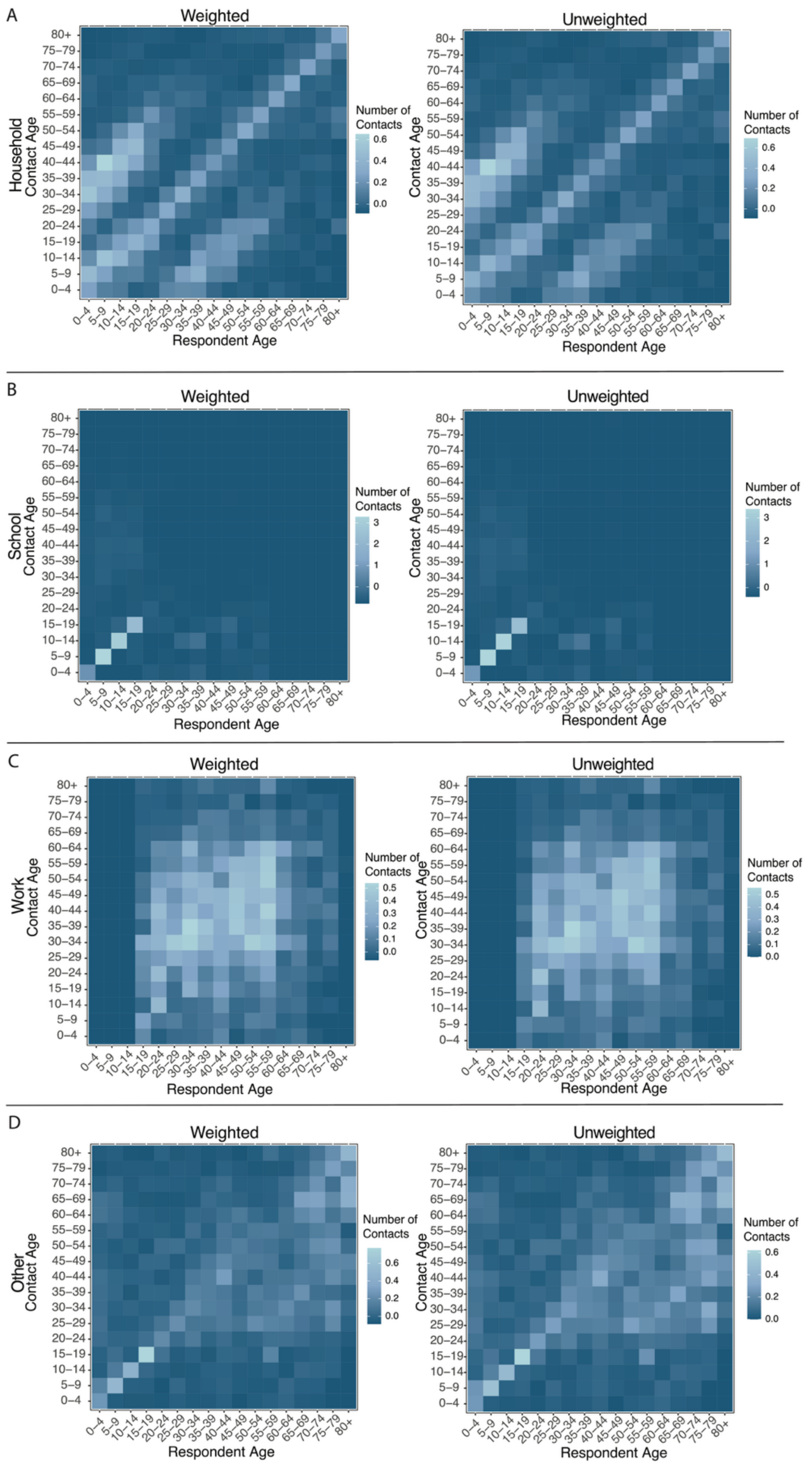


**Figure S 12. A** Setting-specific age-stratified contact matrices using number of household contacts from the weighted and unweighted data. **B** As A, but for school contacts. **C** As A, but for work contacts. **D** As A, but for "Other" contacts.

##### *2.5.6 Sensitivity Analysis: Raw Sample Analysis*

We also compare our results for the data after multiple imputations and inferring of multiple contacts and before (raw data). Age-stratified contact matrices estimated from the raw data showed strong positive correlations with contact matrices estimated using the imputed data (Figure S 13A-B). As expected, while showing the same strong, positive relationship school (Pearson's r = 0.982) and work (Pearson's r = 0.932) contact matrices differ slightly more than matrices for other settings (Pearson's r: 0.992). This is due to a large part of the imputations being used for incomplete data from multiple work contacts, including student-teacher interactions which were moved into school contacts. As for raw and imputed matrices, these matrices are almost identical for both total and setting-specific (Figure S 14-Figure S 15A-D).

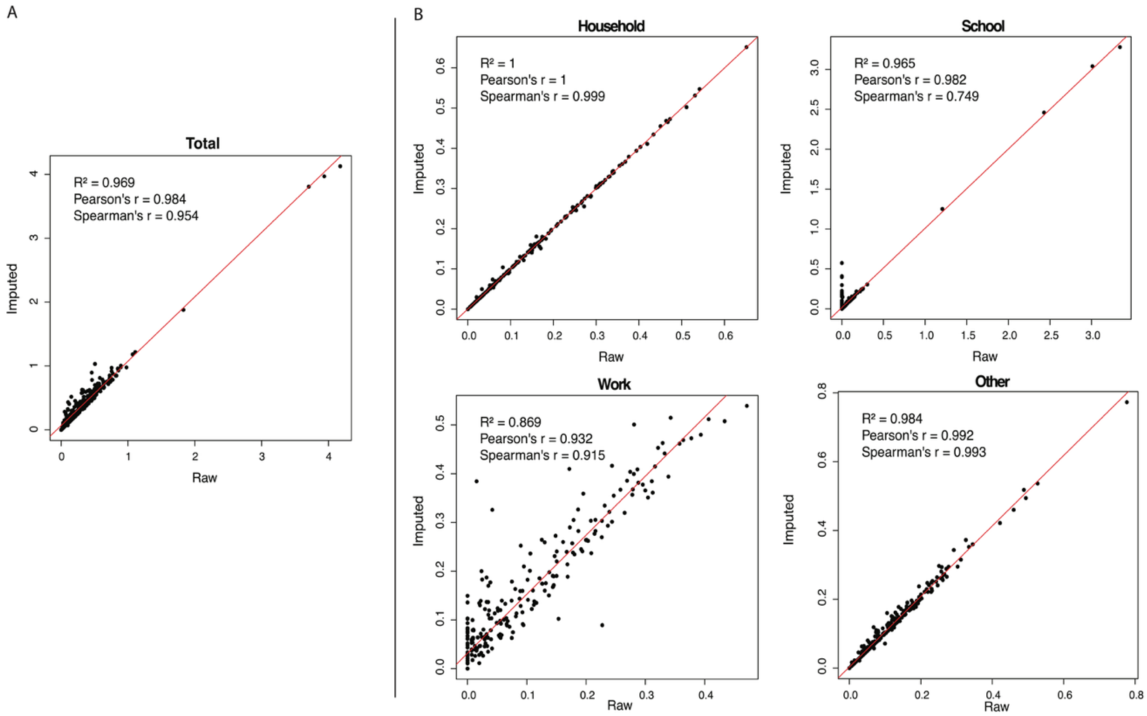


**Fi**

**Figure S 13. A.** Comparison of our estimated contact matrices for the raw data with the imputed data for the total sample. **B.** As A, but setting specific.

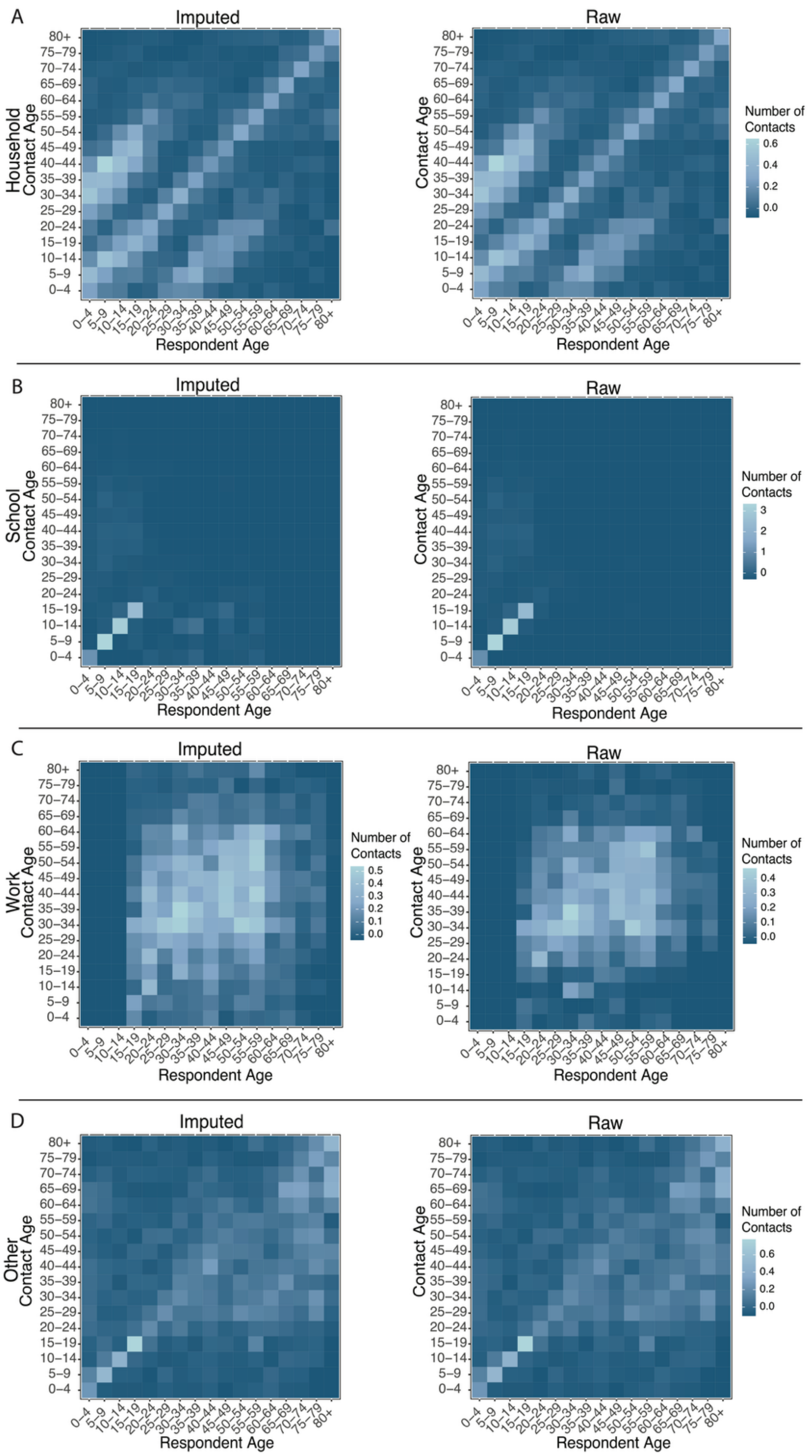


**Figure S 15. A** Setting-specific age-stratified contact matrices for household contacts based on the imputed and raw data. **B** As A, but for school contacts. **C** As A, but for work contacts. **D** As A, but for "Other" contacts.

#### 2.6 Contact Matrices by Race/Ethnicity

##### *2.6.1 Contacts by setting*

Similarly to age-stratified contact matrices, we estimated contact matrices stratified by race/ethnicity (Figure S 16) Overall at the population level, NH White respondents reported the largest number of contacts in the workplace (mean: 2.7) and in the community (mean: 2.2), while Hispanic/Latino respondents had the largest number of contacts in the household (mean: 2.3, Fig. 3A). NH Other and Hispanic/Latino respondents reported the largest number of contacts in school (mean: 1.7 and 1.2, respectively), while NH Black and NH White respondents reported the lowest number of contacts at school (mean: 0.8) with small differences from NH Asian (mean: 0.9). It is important to note that these results represent the average number of contacts of the entire population of each racial/ethnic group, including those who do not work or study.

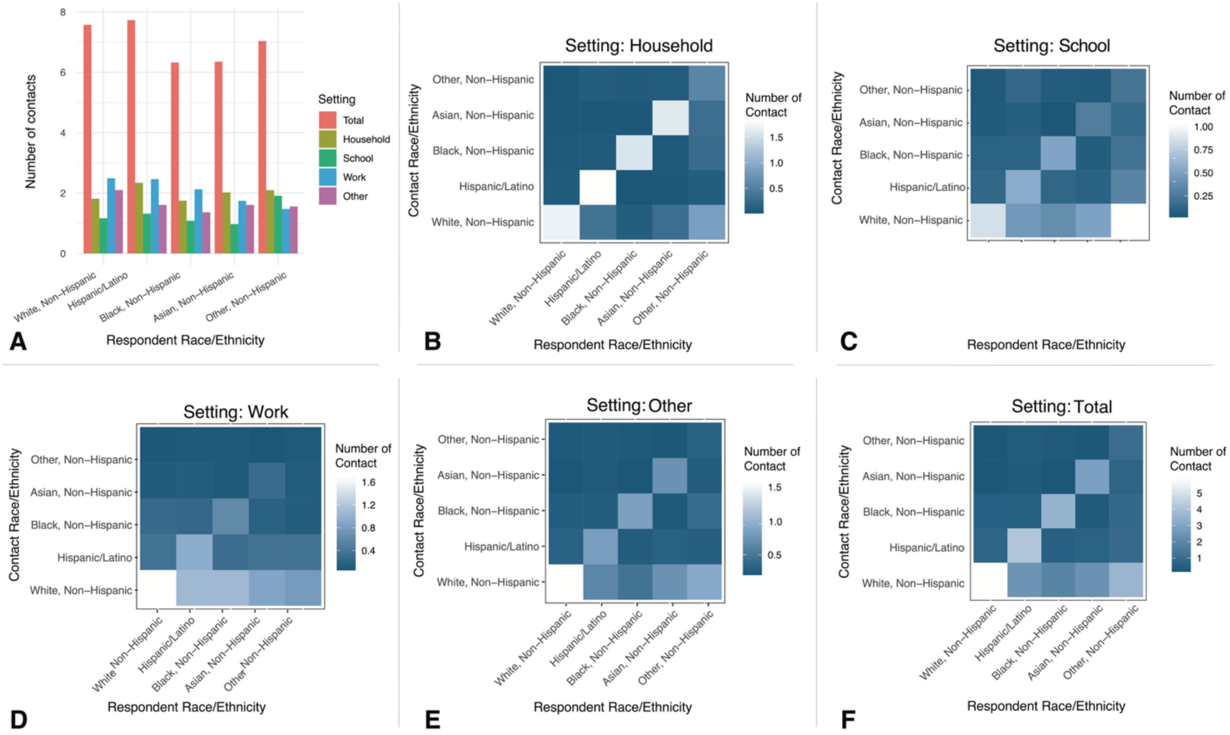


**Figure S 16 A** Number of contacts by race/ethnicity of the respondent in total and disaggregated by social setting. **B** Household contact matrix stratified by race/ethnicity. Each element of the matrix represents the estimated average number of contacts that a respondent of a given race/ethnicity has with individuals of any race/ethnicity. **C** As B, but for contacts at school. **D** As B but for contacts at work. **E** As B but for contacts in "other" (community) settings. **F** As B but for the total contacts.

##### *2.6.2 Conditional Contact Matrices*

Estimated conditional contact matrices stratified by race/ethnicity for each setting are shown in Figure S 17. There is high assortativity of (non-zero) contacts across race/ethnicity groups for all social settings. Among household contacts, assortativity is the highest (Figure S 19), with less but still notable assortativity of school, work and other contacts. Bar plot in Figure S 18 summarizes these contact matrices to show the total number of contacts for a respondent of given racial/ethnic group in a specific setting conditional on having at least one contact in that setting.

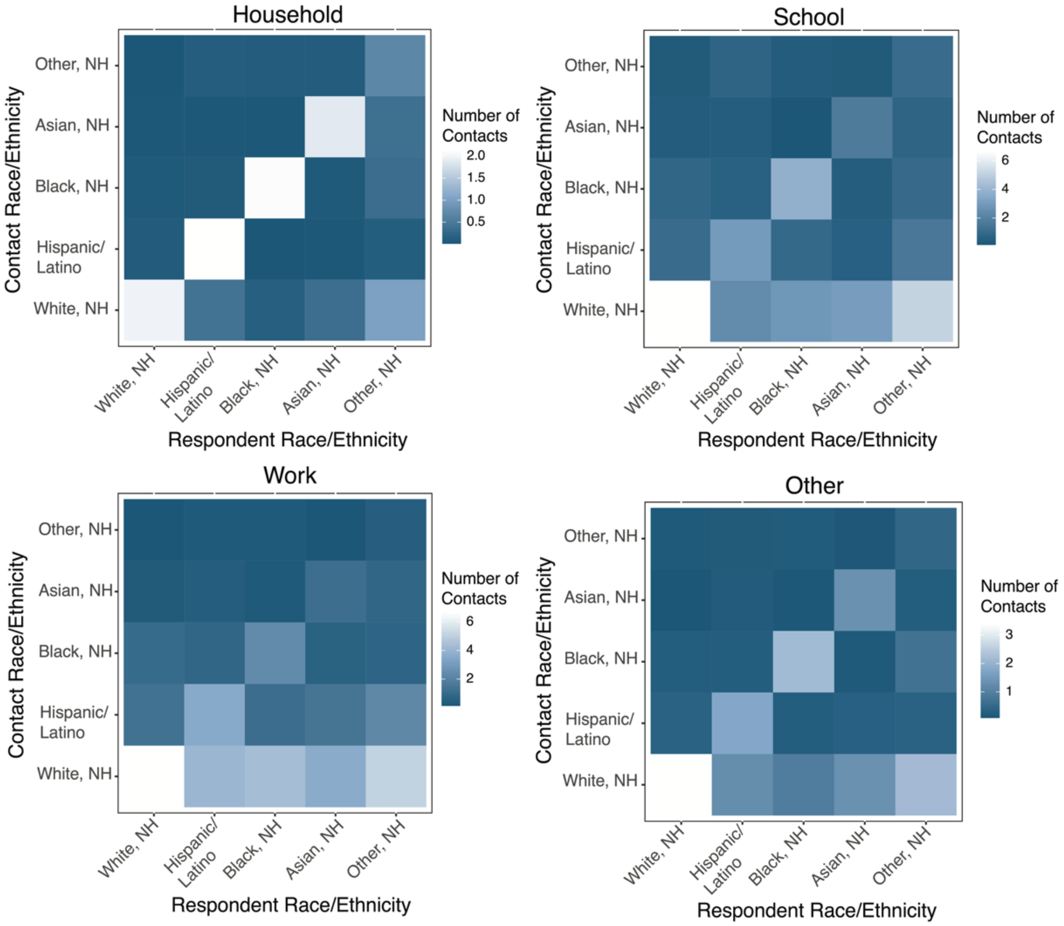


**Figure S 17.** Race-stratified contact matrices by setting ($\boldsymbol{\Theta}_{ij}^{k}$) conditional on the respondent having a contact in that setting.

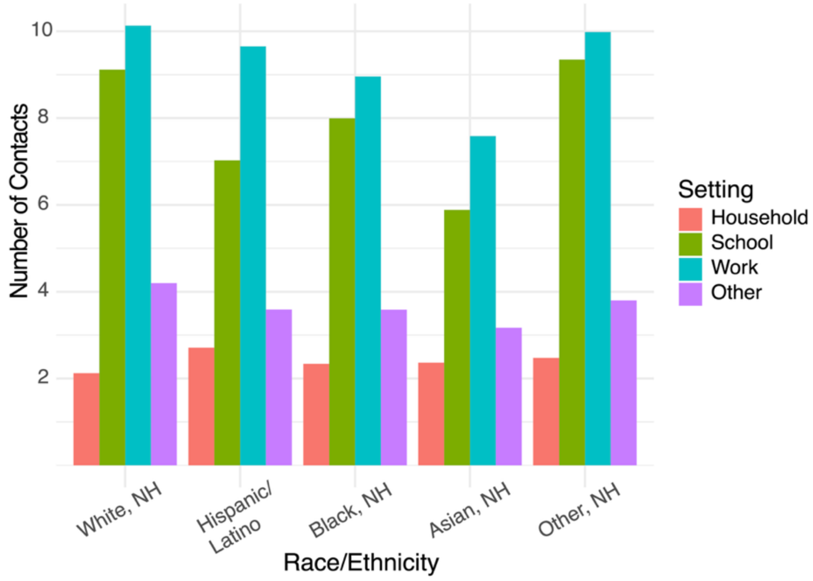


**Figure S 18.** Summary of race-stratified average contacts by setting conditional that the respondent has a contact in the setting.

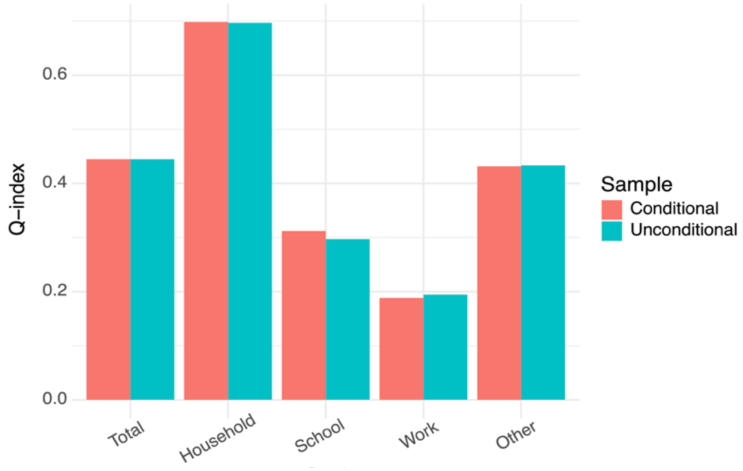


**Figure S 19.** Assortativity by race and by setting.

#### 2.7 Contact Matrices by Sex

Estimated sex-stratified matrices are presented in Figure S 20 and estimated sex-stratified conditional matrices are presented in Figure S 21. With respect to the contacts between male and female individuals, we observed disassortative household mixing that include a significant share of the contacts between spouses of the opposite sex, and highly assortative mixing at school and

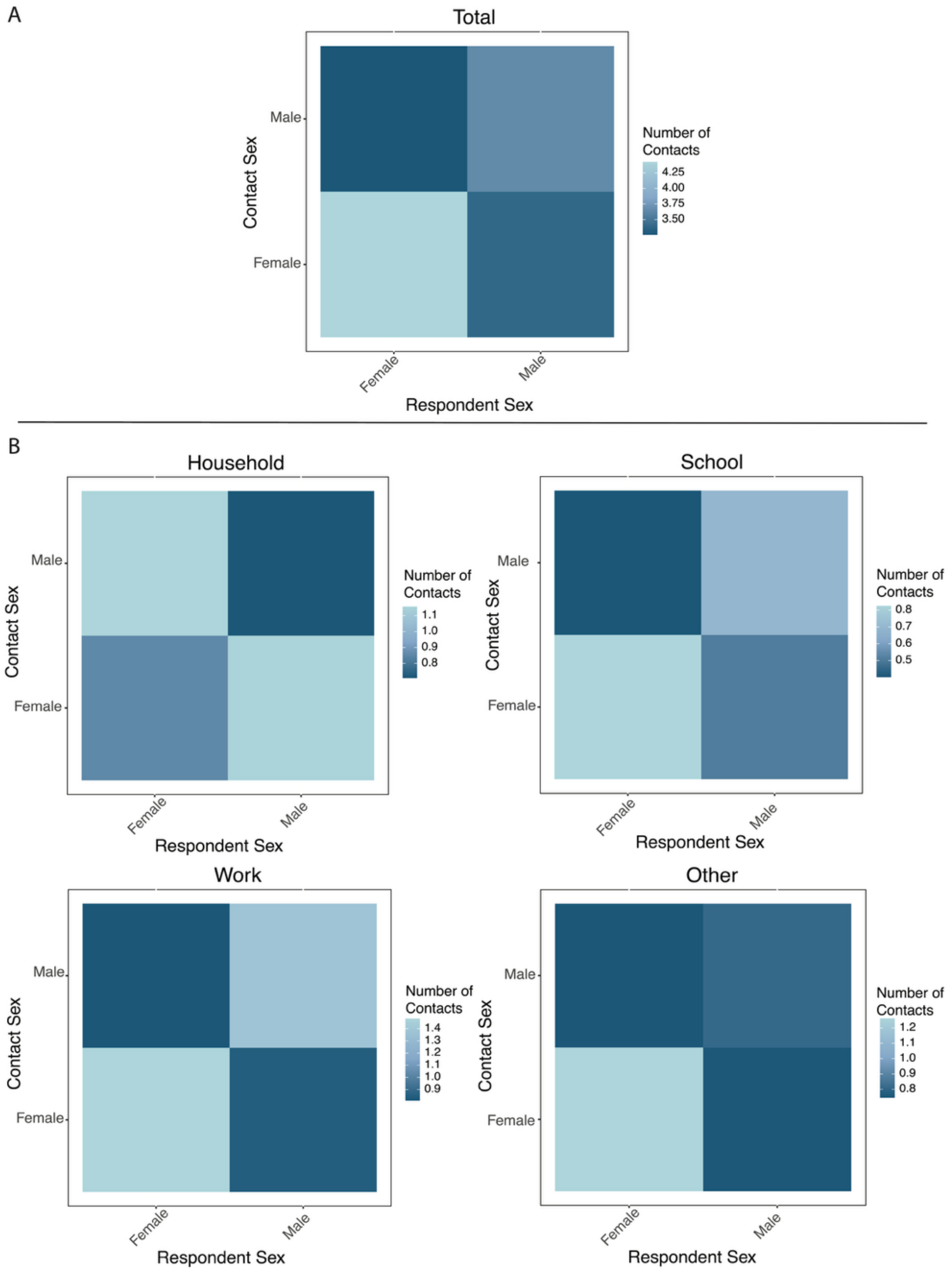


**Figure S 20. A.** Sex-stratified matrices of the number of contacts. **B.** As A, but in the household, school, workplace, or other settings.

work. The assortativity is highest in school (Figure S 22) and always higher for female respondents in non-household settings. The contact matrices by age by sex are presented in (Figure S 23 - Figure S 27). We observe notable differences of contact patterns by age between same-sex and opposite-sex interactions as well as interactions involving females (Figure S 23), mostly as a result of the combination of setting-specific differences (Figure S 24 - Figure S 27). For example, males have more contacts at work after 70 years of age, higher assortativity by age is observed in contacts between males and between females in school, while other contacts between males are much more assortative than across sexes and between females.

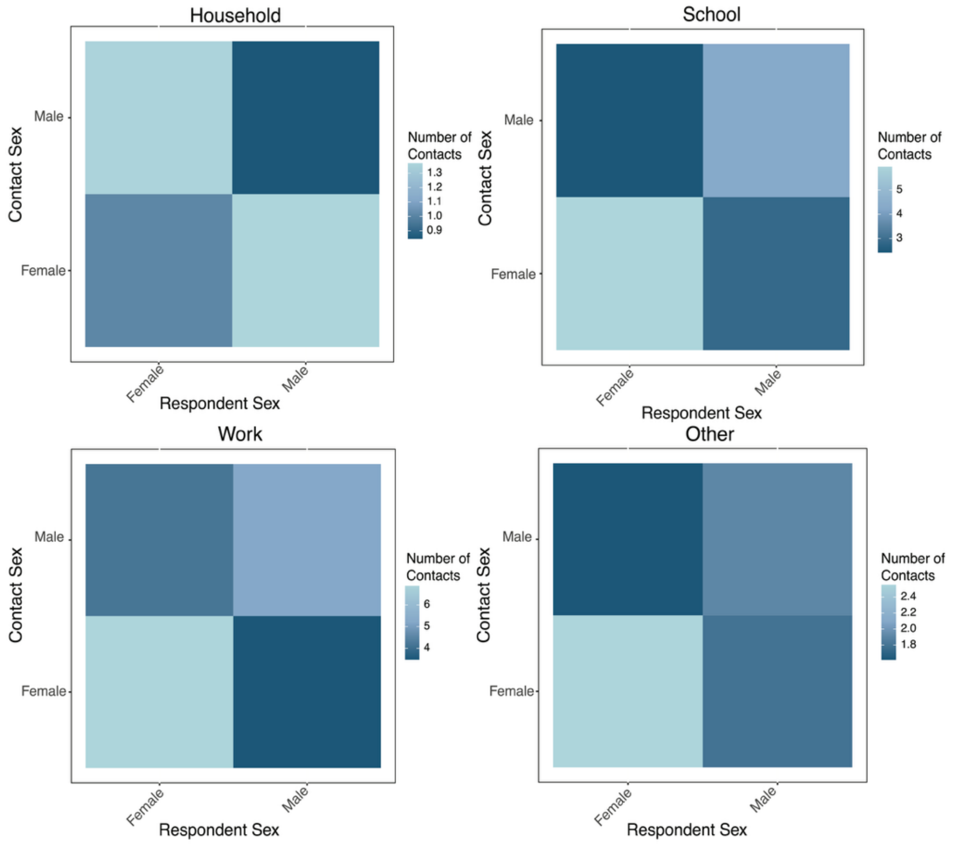


**Figure S 21.** Sex-stratified matrices of the number of contacts by setting ($\boldsymbol{\Theta}_{ij}^{k}$) conditional on the respondent having a contact in the specific setting.

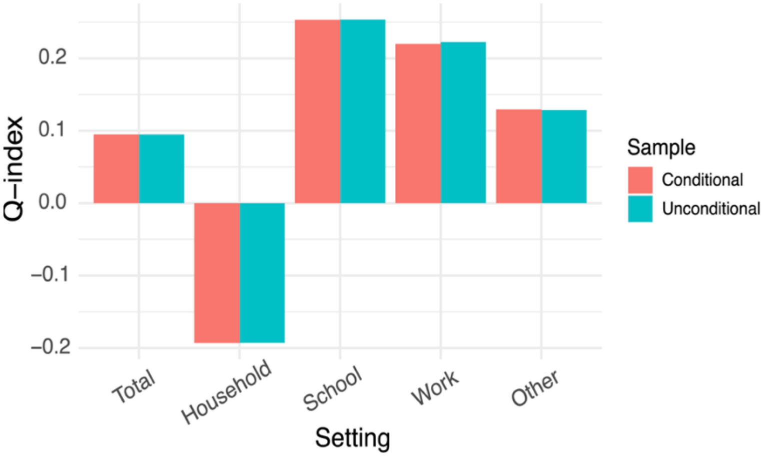


**Figure S 22.** Assortativity by sex and by setting.

##### *2.7.1 Contact Matrices by Age by Sex*

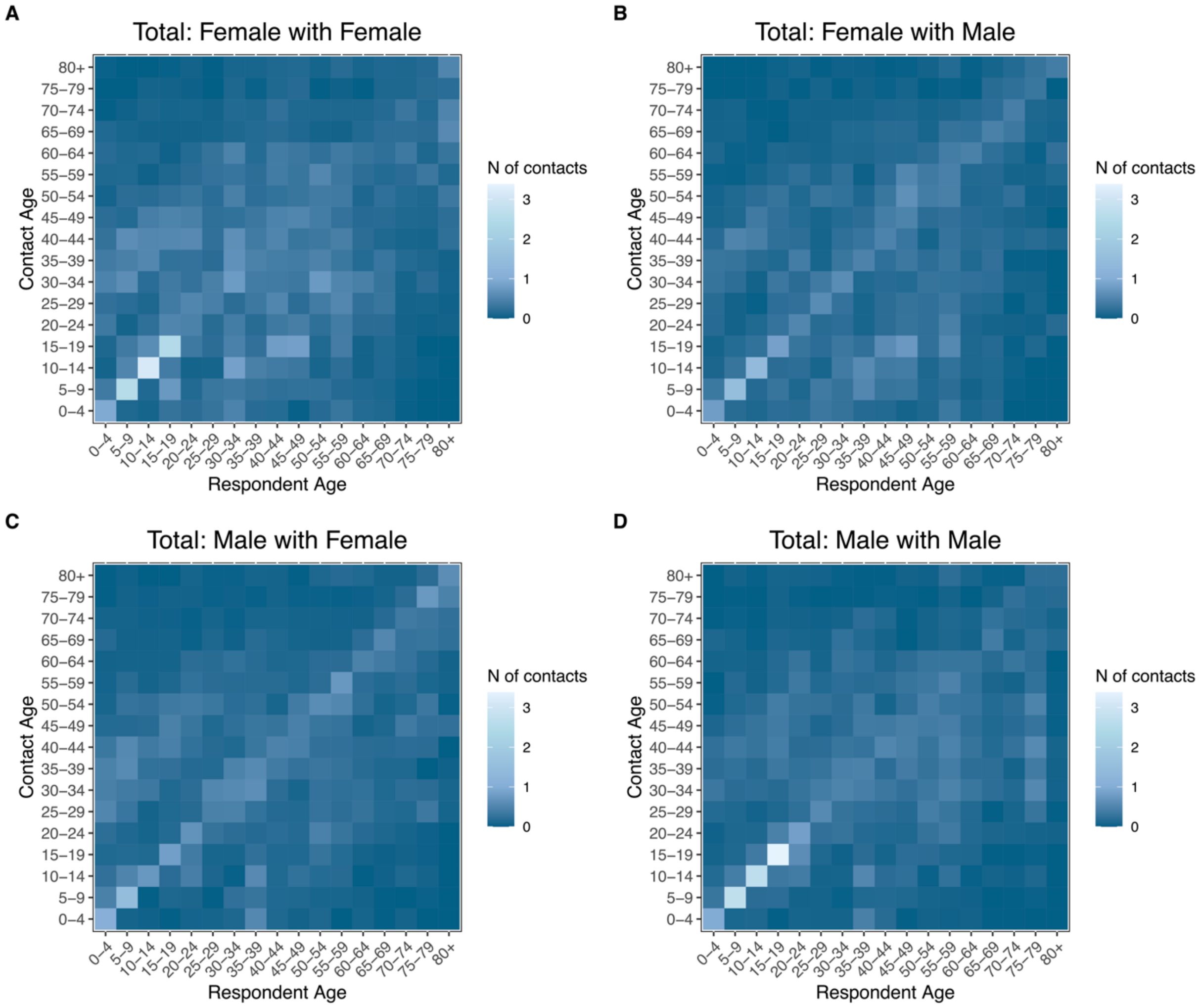


**Figure S 23.** Age-stratified contact matrices by sex for total contacts.

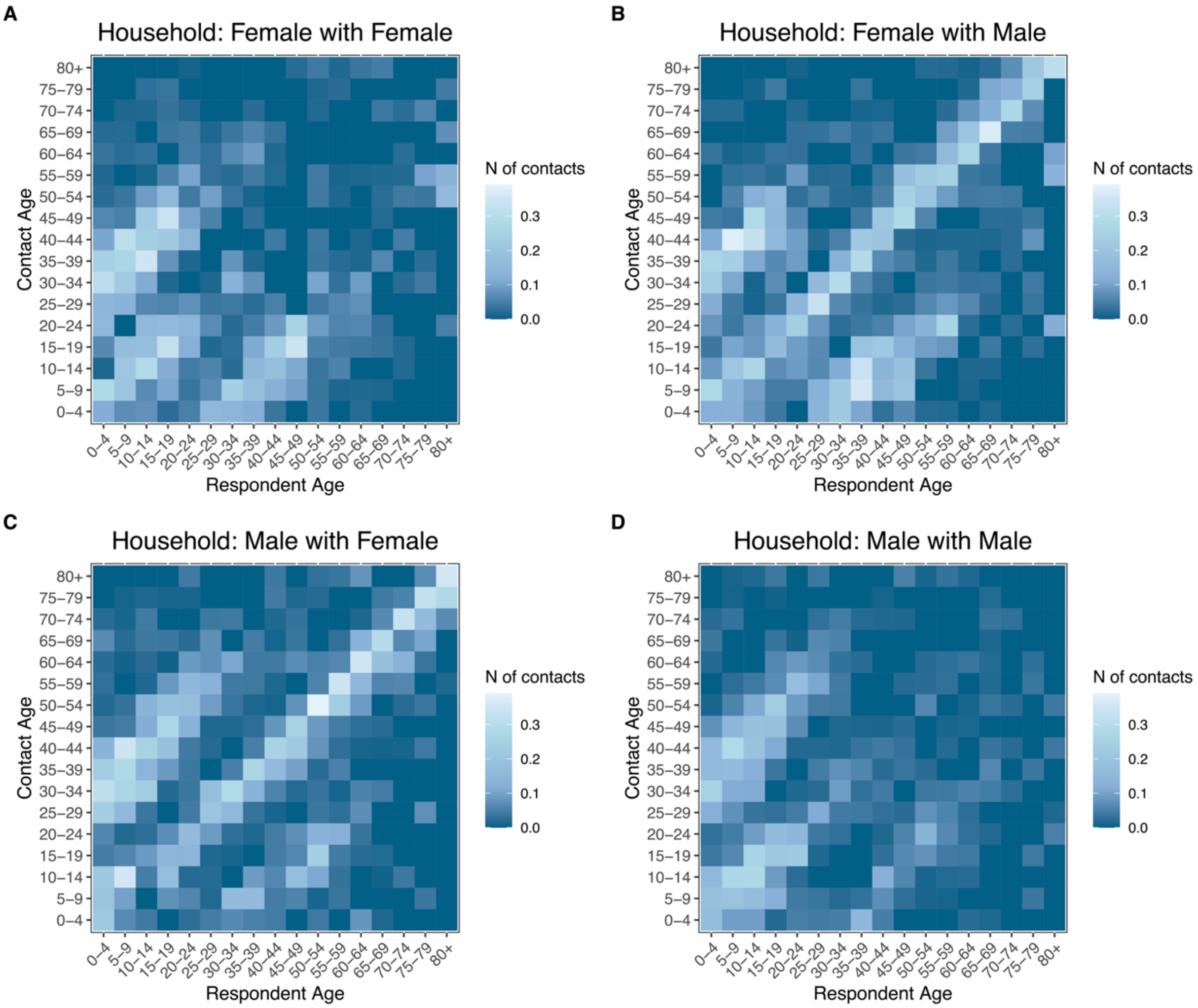


**Figure S 24.** Age-stratified contact matrices by sex for household contacts.

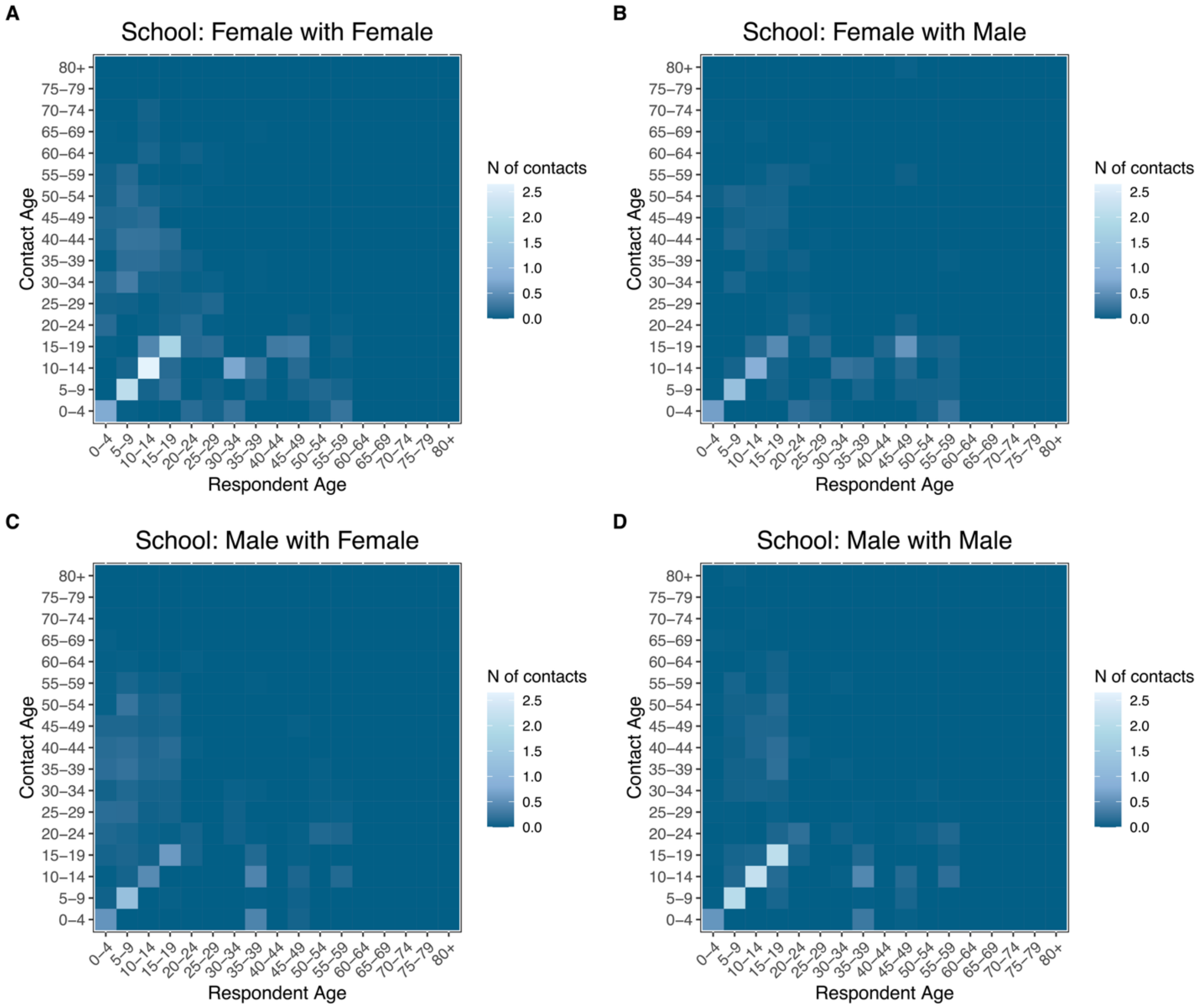


**Figure S 25.** Age-stratified contact matrices by sex for school contacts.

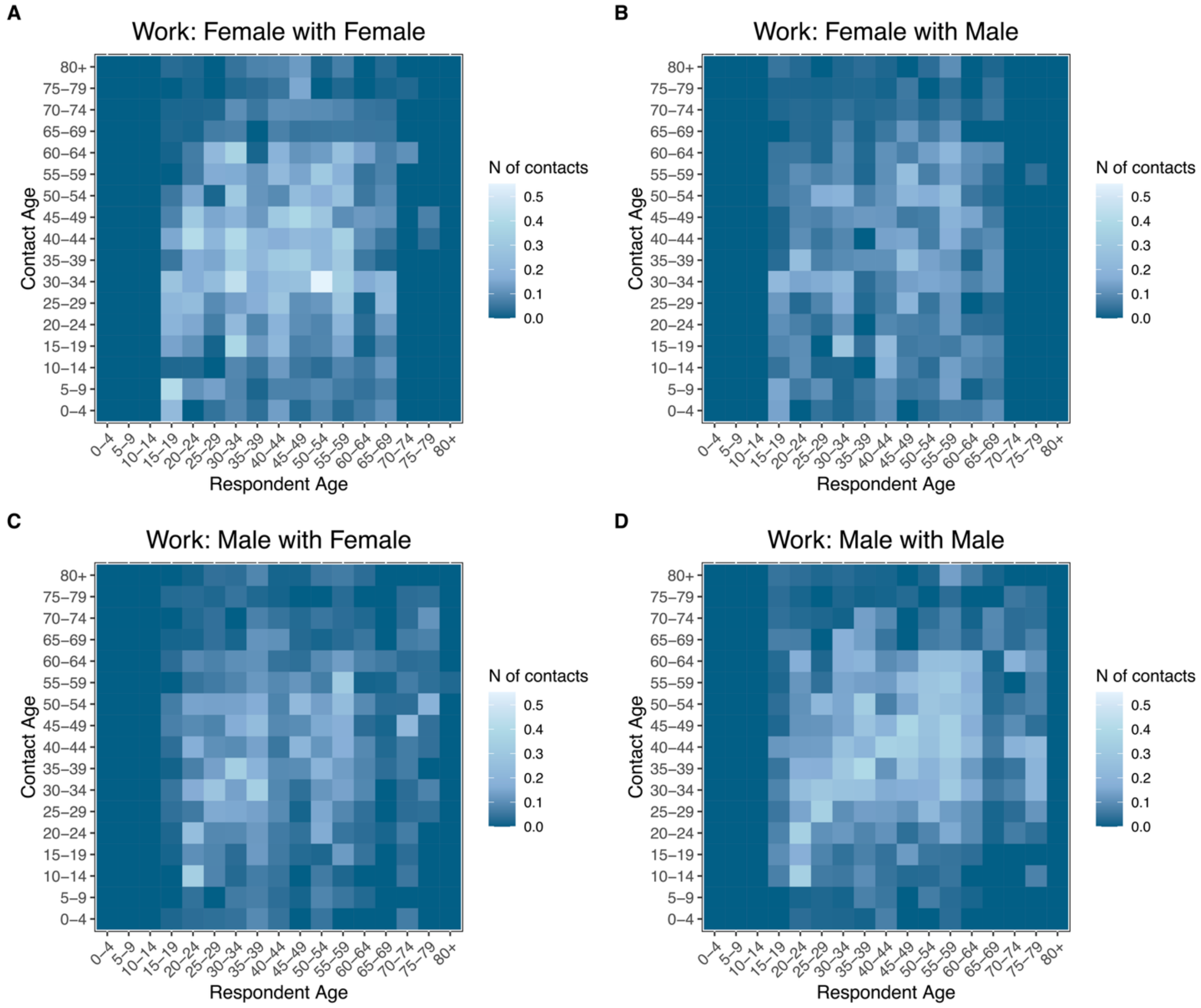


**Figure S 26.** Age-stratified contact matrices by sex for work contacts

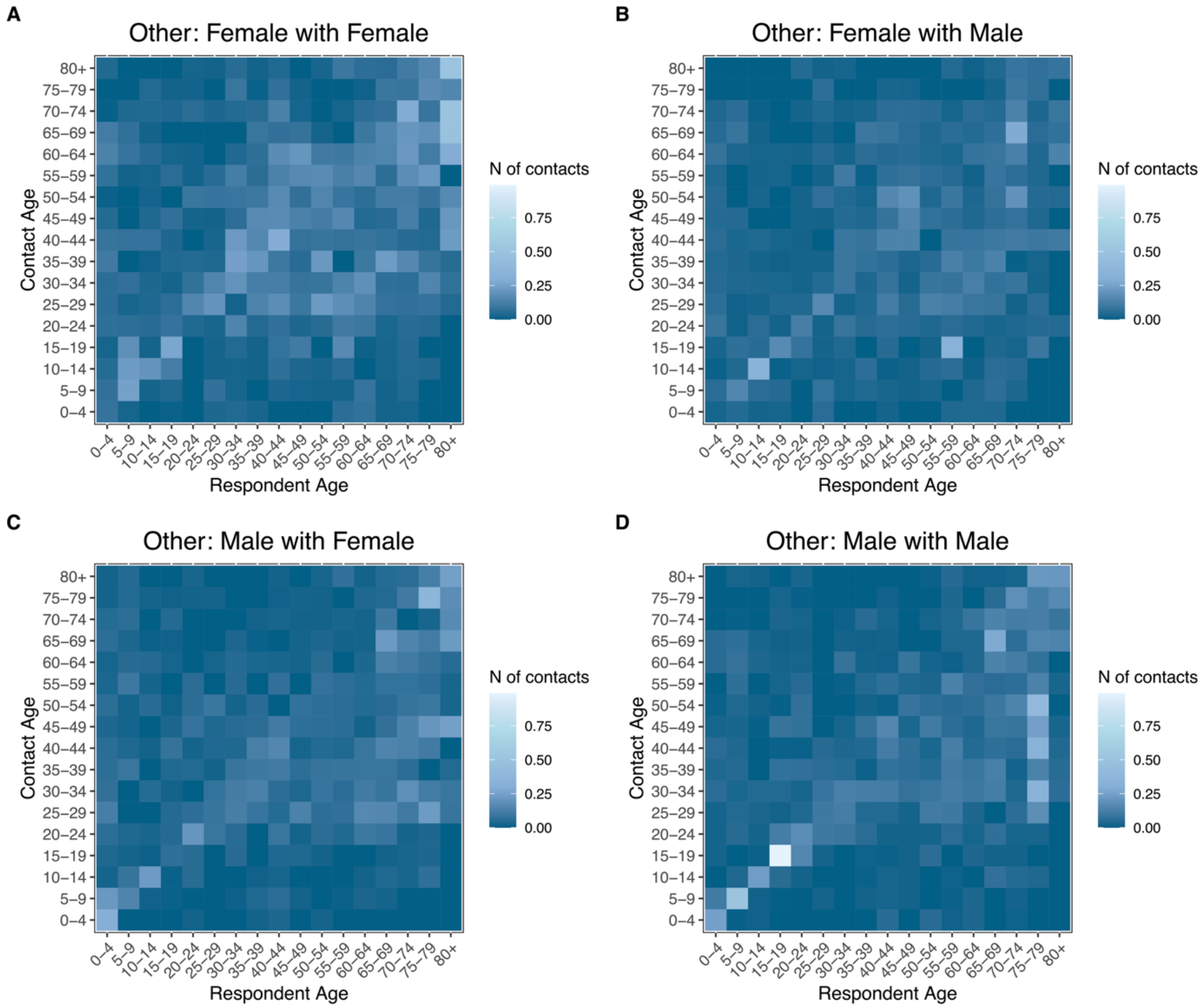


**Figure S 27** Age-stratified contact matrices by sex for other contacts.

#### 2.8 Comparison With Pre-Pandemic Age-Stratified Contact Matrices

##### *2.8.1 Pre- and Post-Pandemic*

Estimated contact matrices showed strong positive correlations with contact matrices inferred for the US in the literature during the pre-pandemic era[35,41] (Figure 3). The differences with Mistry et al.[35] are relatively random. In contrast, for Prem et al.[41], differences mostly arise from higher number of contacts in diagonal-adjacent age groups and lower number of contacts with those 60 years of age and above. The correlations between elements of age-stratified contact matrices for fully in-person scenario and the pre-pandemic period for the U.S. are shown below (Figure S 28). The magnitude of correlations is slightly lower but comparable than between the main analysis matrix and the pre-pandemic estimates.

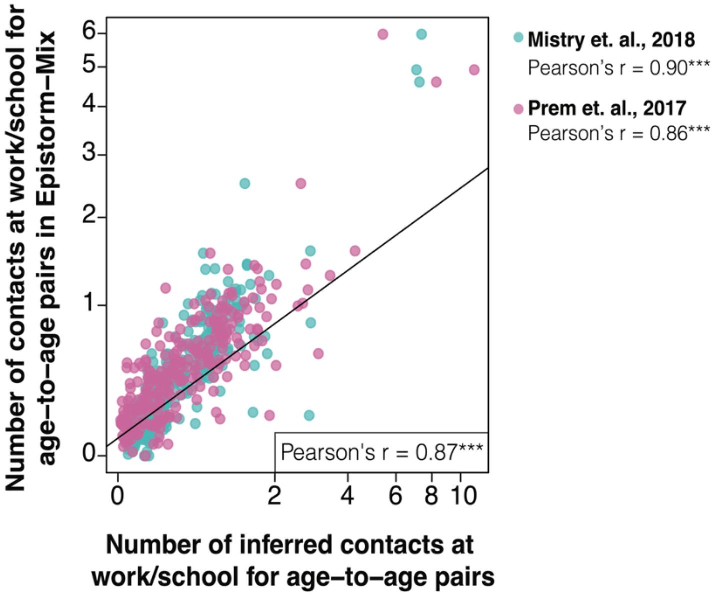


**Figure S 28.** Comparison of number of contacts for pre-pandemic estimates for fully in-person.

##### *2.8.2 Pre- and Post-Pandemic Frequency-Based Contact Matrices*

Figure 4 in the main text shows correlations between estimated frequency-based contact matrices by setting and pre-pandemic estimates of the synthetic frequency-based contact matrices from Mistry et al[35]. We observe high correlations for household, school, and work contacts. We observe no statistically significant correlation for other contacts, mostly because random mixing was assumed in the synthetic matrix approach and the per-capita probability of contact reflect the age structure, which is not the case of estimated contact matrices in this study.

#### 2.9 Comparisons with Other Countries

Figure S29 compares the US estimated age-based contact matrix with post-pandemic survey-based contact matrices from several other countries. Overall, the US estimates show a moderately strong and positive correlation (Pearson's $r$: 0.66 – 0.90, p-values < 0.001) with matrices from the UK, Belgium, the Netherlands, Switzerland, Italy, and two provinces in China (Suzhou and Hunan). While the statistical correlation was strongest with data from Switzerland and Suzhou, the ranges of the absolute number of contacts in the US estimates were more in line with the ranges reported in the UK, the Netherlands, Italy, and Belgium. The differences with the Netherlands arise from a higher number of off-diagonal interactions with minors and those 70+. For the UK, the differences were due to the US estimates being higher for contact among working-age adults and within school-age groups (mostly 10-50%), indicating a potential impact of higher involvement of UK workers in remote labor. In contrast, Switzerland's data showed higher contact rates than the US estimates across most age groups, especially in diagonal-adjacent age groups. The comparison with Italy revealed more distinct differences in social patterns. US contact rates are higher between young people (under 25) and adults aged 15-44, but lower between the under-25 group and older adults aged 40-70. This may reflect different family structures in Italy, such as lower fertility rates and a higher average age for parents. Finally, the differences were less pronounced for the two Chinese provinces. Compared to both Suzhou and Hunan, the US estimates were higher for the 10-19 age group. Estimates were also slightly lower for some age groups on the diagonal in Suzhou and for adults in their 70s in Hunan.

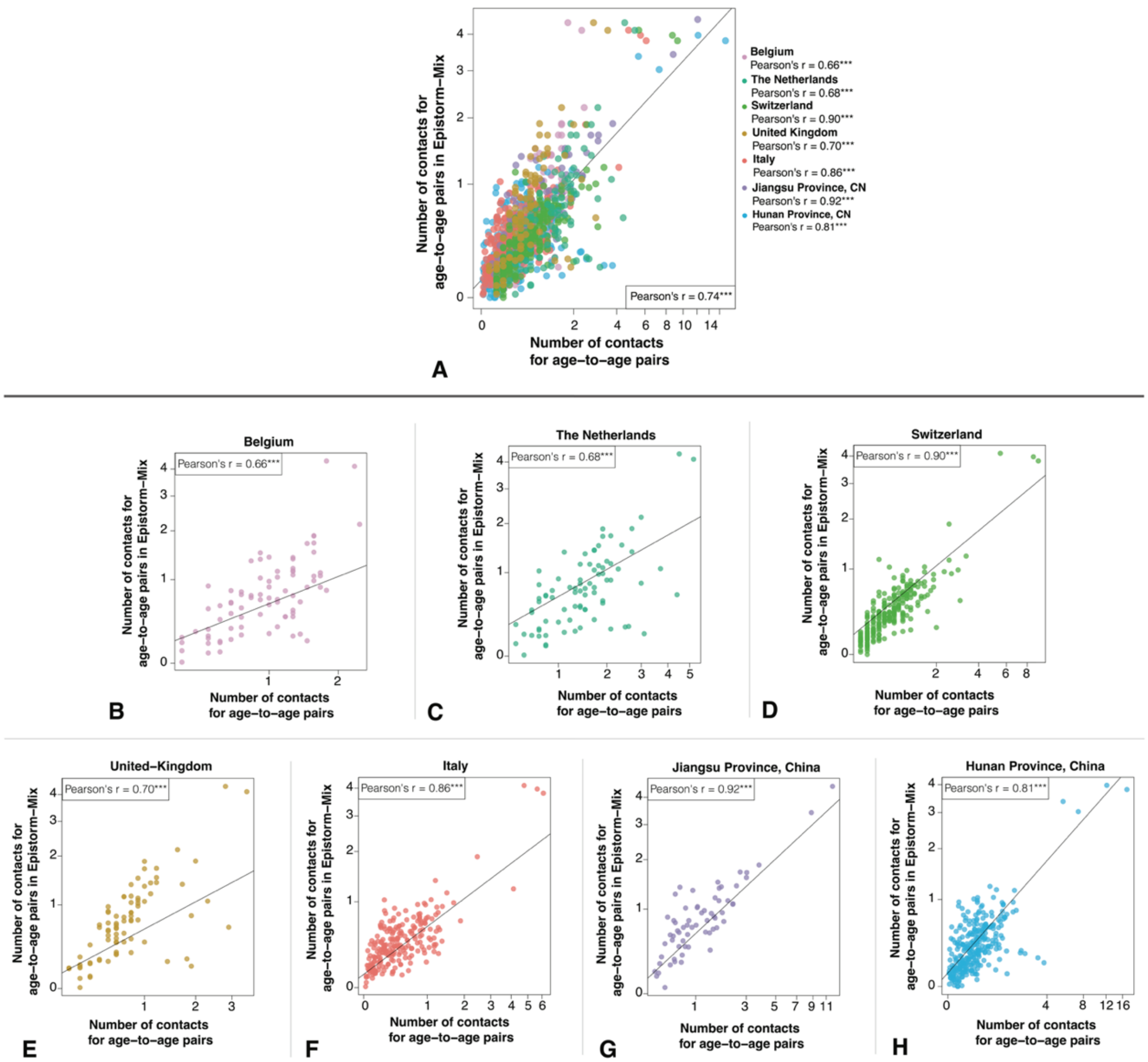


**Figure S 29.** Comparison of estimated US contact matrices with post-pandemic contact matrices for other countries.

When simulating the spread of an epidemic with country-specific contact matrices while assuming the same reproduction number, generation time, and initial conditions, we found broadly similar epidemic dynamics (peak timing and peak incidence) and infection attack rates by age (Fig. S30). The two main differences are: i) the lower incidence, peak incidence, and later peak in the Hunan province of China, where epidemic dynamics are mostly driven by school-age individuals with much higher number of contacts; and ii) the infection attack rate by age in Belgium, which is shifted from school-age individuals to working-age adults.

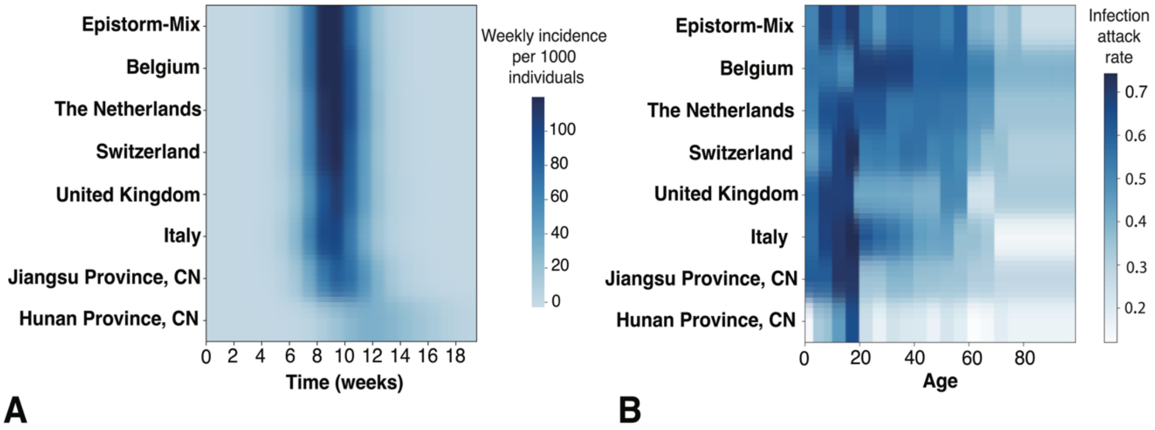


**Figure S 30. A** Comparison of weekly incidence (infections per 1000 individuals) based on SIR model simulations using different country-specific contact matrices. **B** Comparison of infection attack rate based on SIR model simulations using different country-specific contact matrices
